## Supplementary Material for "The impact of health inequity on regional variation of COVID-19 transmission in England"

5 Software and implementation 73

List of Figures 77

List of Tables 78

### 1 Data Gathering and Processing

In this section we provide an in-depth description of all data sources utilised and the processing applied before being integrated into the model.

#### 1.1 Geographic Data

We conduct our analysis at the Lower Tier Local Authority (LTLA) level. Local authorities are responsible for a wide range of provisions to their populations including health services, social services, housing, and environmental health. London is separated into multiple LTLAs via London boroughs. Populations of LTLAs vary, the smallest we consider is Rutland in the East Midlands, with a population of 40,476, the largest is the city of Birmingham with a population of 1,140,525.

These boundaries change from year-to-year. We use the 2021 boundary definitions which most closely resemble the boundaries by which epidemiological data is provided. The shape files for these boundaries is taken from the Office for National Statistics (ONS) website [1]. There are 309 English LTLAs within this data set. While epidemiological data is provided at the LTLA level, some LTLAs with small populations are combined in the UK online COVID-19 data dashboard [2], in order to protect individuals' privacy. Hence, to align better with the epidemiological data we combine the LTLAs of "Cornwall" and "Isles of Scilly", and we combine the LTLAs of "City of London" and "Hackney". We also do not consider the "Isle of Wight" LTLA, as it is the only remaining English LTLA that doesn't share a land barrier with another LTLA - a unique complication to our modelling which was better removed that included.

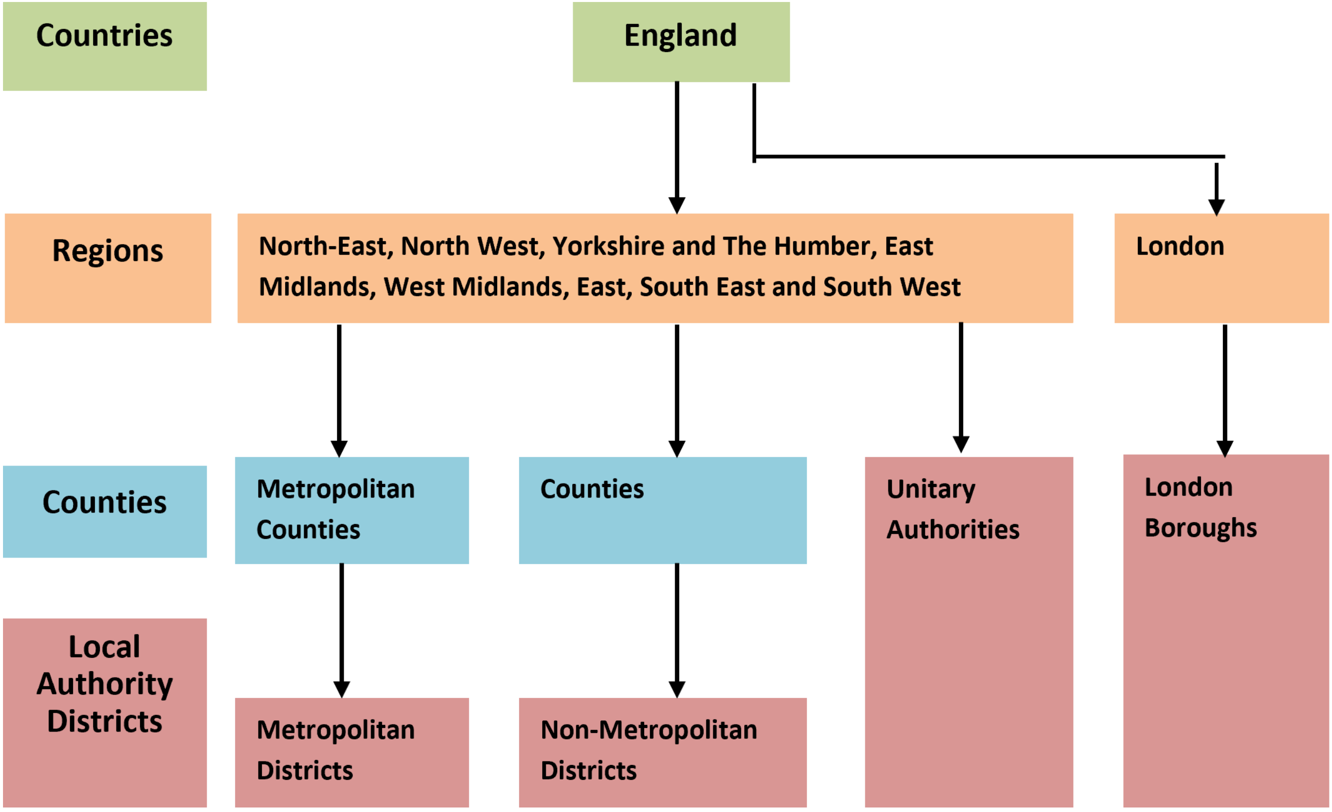

**Figure S1:** The administrative geography hierarchy in England. The country (green) can be separated into 9 NHS regions (orange). Some of these regions are then further separated into counties (blue), before ultimately separated into local authorities (red). Lower tier local authorities (LTLAs), the administrative level we consider, are the combined red boxes. Figure is taken from the mid-2019 ONS population report [3].

#### 1.2 COVID-19 Cases Data

Our model is fit to data of COVID-19 cases. A detailed line list of all COVID-19 cases is maintained by the UK Health Security Agency (UKHSA), collected by the Department of Health and Social Care. The location of each case is recorded, whether the test was via polymerase chain reaction (PCR) or via a self-performed lateral flow device (LFD), and what "pillar" the test was performed as part of. Testing in the UK is divided into four "pillars" of testing:

- Pillar 1: testing in Public Health England labs, NHS hospitals for those with a clinical need, and health and care workers.

- Pillar 2: swab testing for the wider population.
- Pillar 3: serology testing to show if people have antibodies from having had COVID-19.
- Pillar 4: blood and swab testing for national surveillance and research partners.

To best capture the number of cases in the general population for each LTLA, we extract only pillar 2 cases, and only those confirmed by PCR. We then aggregate all cases by LTLA, and aggregate by week that the test was conducted (weeks beginning Monday).

We also extract the number of confirmed first episodes (an instance of an individual becoming infected with COVID-19 for the first time), via PCR or LFD, pillar 1 or pillar 2, to integrate the estimated amount of acquired immunity present in the population of an LTLA. Number of first episodes can be directly extracted from the UK COVID-19 online dashboard [2]. Due to boundary changes occurring during the pandemic, we aggregate some of the LTLAs from this data set back into their original 2021 LTLAs. The LTLAs of South Bucks, Aylesbury Vale, Chiltern, and Wycombe are aggregated into the LTLA of Buckinghamshire. Northampton, South Northamptonshire, and Daventry are aggregated into the LTLA of West Northamptonshire. East Northamptonshire, Wellingborough, Corby, and Kettering are aggregated into the LTLA of North Northamptonshire. These first episodes are similarly then aggregated by week.

#### 1.3 Variants-of-Concern Data

For each LTLA, for each week, we record what proportion of the cases for that week were of the original "wild-type", the Alpha variant, the Delta variant, or the Omicron variant. The UK online dashboard reports the number of variants-of-concern (VOCs) identified via whole genome sequencing (WGS), however this is only available at the spatial level of the nine NHS regions. The UKHSA line list however records the result of spike gene target failure (SGTF) for PCR-confirmed cases. This simplified test relies on the fact that the Alpha (B.1.1.7) and Omicron (B.1.1.529, BA.1, BA.4 and BA.5) variants of concern (VOC) share specific mutations in their spike gene not observed for the Delta variant or original wild-type. McMillen et al. (2022) [4] report that at the peak of their corresponding wave, the positive predictive value of the SGTF was 98% for Alpha and 100% for Omicron. As such, we classify each case from the linelist into variants via the following criteria:

From the start of the pandemic to March 27th 2021, s-gene positive cases are tagged as "wild-type", s-gene negative cases are tagged as "Alpha variant".

From March 28th 2021 to September 11th 2021, s-gene positive cases are tagged as "Delta variant", s-gene negative cases are tagged as "Alpha variant".

From September 12th 2021 to January 1st 2022, s-gene positive cases are tagged as "Delta variant", s-gene negative cases are tagged as "Omicron variant".

From January 2nd 2022 onwards, we assume all cases are of the Omicron variant, as the BA.2 subtype is s-gene positive, and cases of Delta are seen to be negligible from this point on nationally.

From this classification, we record for each LTLA, weekly, what proportion of cases are made up of each variant.

#### 1.4 Population Data

This study considers the impact of LTLA-specific measures of socio-demographic and socio-economic data.

Data on the ethnic makeup of each LTLA's population is taken from a 2019 ONS report [5] estimating the 2016 population proportions by ethnicity based on 2011 census data. Ethnicity is divided into "White British", "All Other White", "Mixed / Multiple ethnic groups", "Asian / Asian British", "Black / African / Caribbean / Black British", and "Other ethnic group". We aggregate "White British" and "All Other White" into one group, and aggregate "Mixed / Multiple ethnic groups" and "Other ethnic group" into one group, leaving four ethnicity categories in total. This data does not vary across weeks.

The Index of Multiple Deprivation (IMD) is a metric given by the Ministry of Housing, Communities & Local Government (MHCLG), as a score to denote levels of deprivation across areas of the UK. A higher score indicates a higher level of deprivation. The index captures income deprivation, employment deprivation, education, skills and training deprivation, health deprivation and disability, crime, barriers to housing and services, and living environment deprivation. A score is provided for each LTLA in England. Full methodological details of its calculation can be found in the 2019 report the data was taken from [6]. We use these fixed estimates for all weeks.

The population, population density, and proportion of the population over the age of 65, is all taken from the mid-2019 ONS dataset [3] at the LTLA level. We use these fixed estimates for all weeks.

Data on the median annual income for each LTLA is taken from the 2021 and 2022 editions of the ONS Annual Survey of Hours and Earnings (ASHE) [7]. Earnings data is split by financial year; all weeks before April 6th 2021 use the 2020 earnings data, and all weeks after April 6th 2021 use the 2021 earnings data.

Data on time spent in locations is taken from the Google Community Mobility Reports [8]. For each LTLA, and for each day, the reports give the percent difference in time spent at a category of location compared to a pre-pandemic baseline (the median value from the 5-week period January 3rd – February 6th, 2020). Seven location categories are provided: Retail and recreation, grocery and pharmacy, parks, transit stations, workplaces, and residential. The residential category shows a change in duration — the other categories measure a change in total visitors. We use only the residential, workplace, and transit station categories in our analysis. For each week, we use the mean value across the seven days. As with first episodes data, we also average together the scores for the LTLAs making up the aggregate regions of Buckinghamshire, West Northamptonshire, and North Northamptonshire. In a small number of instances, where NAs are given for each day of that week for an LTLA, we replace this NA value with the average of the two weeks either side of this NA date. For example, if a residential percentage for Birmingham was -50 on week 10, NA on week 11, and -60 on week 12, we would set week 11 to -55.

Rutland, the smallest LTLA (population 40,476), has an additional 31 NA values that cannot be resolved as outlined above, due to its particularly small population. For these 31 weeks, we take the average value of all of Rutland's neighbours to use for its values for residential visit duration and transit station visits covariate values.

#### 1.5 COVID-19 Funding Data

During the pandemic extra funding was provided to administrative regions to help support efforts to reduce the spread of disease and support the region's communities, a full list of all funding allocations is given in the gov.uk funding report site [9]. We extract data on the three biggest funding provisions:

Unringfenced funding tranches, totalling £4,607 million in the 2020/21 financial year, and £1,550 million in the 2021/22 financial year. This funding is provided with limited restrictions for LTLAs to spend as they see fit.

The Contain Outbreak Management Fund (COMF) (including Test and Trace Support Grant), a funding provision for services including targeted testing of hard-to-reach groups, additional contact tracing, communication and marketing, and self-isolation support (full criteria can be found in the online guidance report [10]). £1,717 million was provided in 2020/21, and a further £400 million in 2021/22.

The Adult Social Care (ASC) infection control fund is funding specifically allocated to help support care homes, by supporting and paying staff who are self-isolating or additional hiring and facilitation needs (full guidance available online [11]). £1,146 million was provided in 2020/21, and a further £592 million in 2021/22.

In some instances, money was additionally assigned to Upper Tier Local Authorities (UTLAs) (blue areas in Figure S1), mostly for rural counties. In these instances, we divided these UTLA funds between the LTLAs they cover, proportional to their respective populations.

Money was allocated to LTLAs based on their respective populations and estimated health needs. This data varied by LTLA and by financial year. This data was divided by the LTLA population to include in the model as a "funding per person" value.

#### 1.6 Data visualisation

A total of 16 spatially (and in some instances temporally) heterogeneous variables, as detailed above, are considered;

- The proportion of the LTLA population that is Asian / British Asian.
- The proportion of the LTLA population that is Black / African / Caribbean.
- The proportion of the LTLA population that is mixed / multiple / other ethnicity.
- The average IMD score.
- The proportion of the LTLA population that is over the age of 65.
- The population density of the LTLA, in population per squared kilometer.
- The median annual income in the LTLA.
- The percent change in the number of people visiting a workplace.
- The percent change in the number of hours spent at places of residence.

- The percent change in the number of people visiting a transit station.
- The proportion of cases that are Alpha variant that week.
- The proportion of cases that are Delta variant that week.
- The proportion of cases that are Omicron variant that week.
- The amount of unringfenced COVID-19 funding allocated per person.
- The amount of COMF funding allocated per person.
- The amount of ASC infection control funding allocated per person.

The impact of the ethnicity variables are considered against the White British / Other White proportion baseline. The VOC proportions are considered against the wild-type baseline.

Figure 1 in the main manuscript demonstrates how IMD and the proportion of the population that is white British varies across England. Here we present the variation for all of the above variables considered. In each instance, blue represents the minimum value observed, red the maximum value observed, and yellow is set at the median value observed. The boroughs of London are inset next to the England boundaries for improved readability.

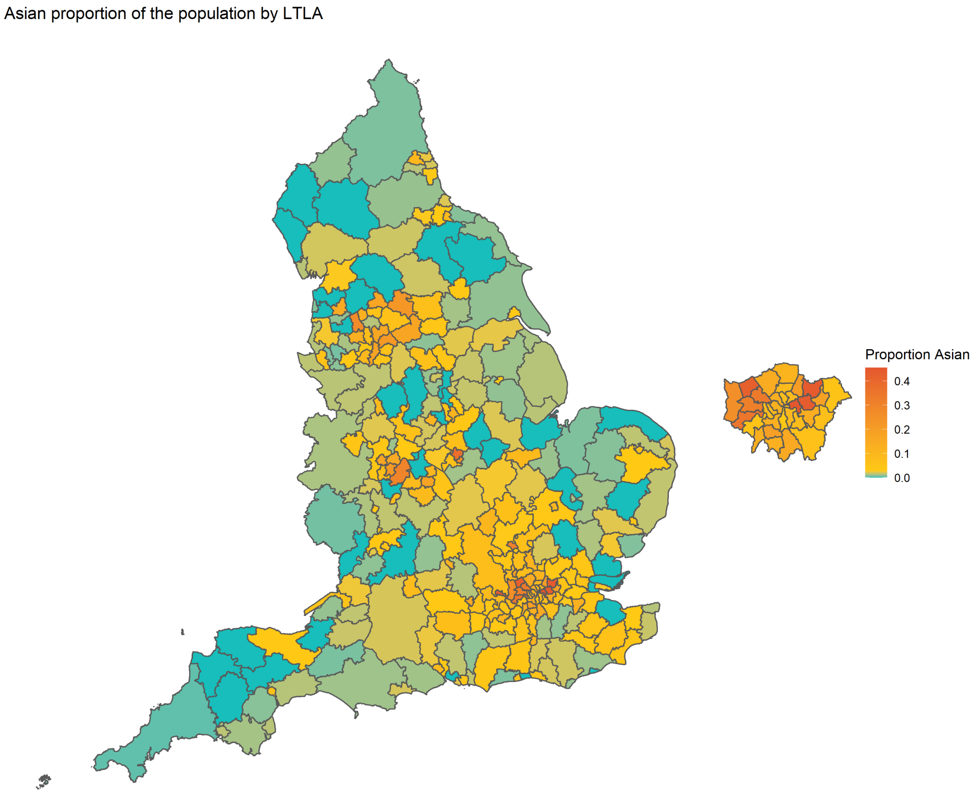

**Figure S2:** The proportion of the population of an LTLA that is Asian/Asian British, as given in the 2019 ONS report [5]. Boundary source: Office for National Statistics licensed under the Open Government Licence v.3.0 [12]. Contains OS data © Crown copyright and database right [2024].

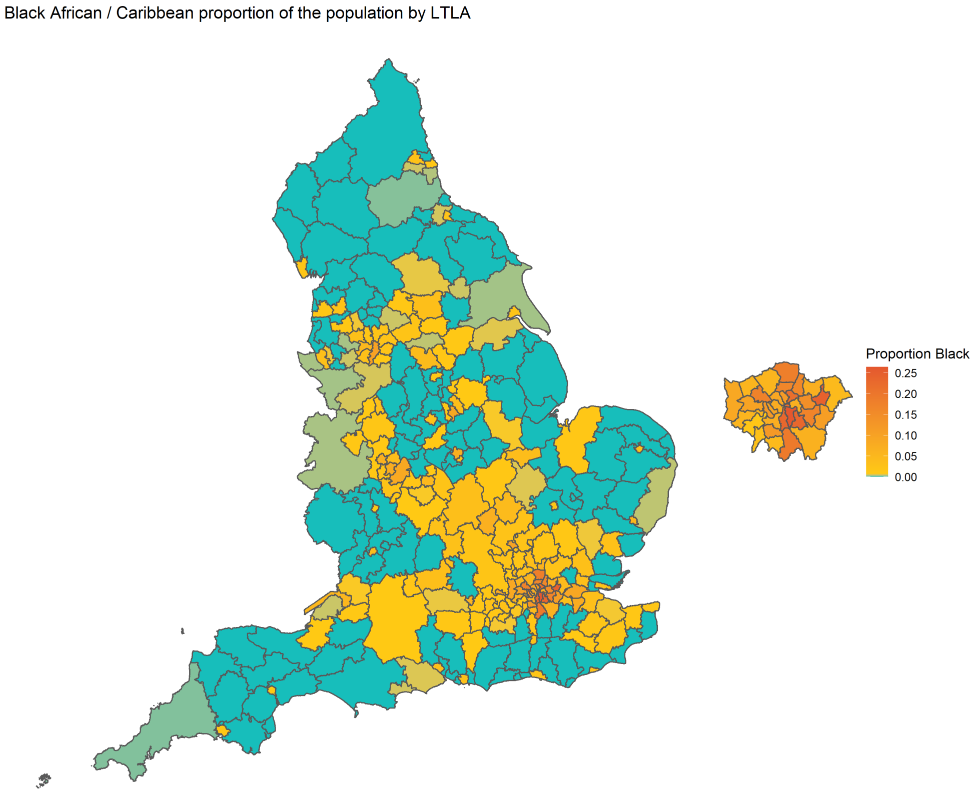

**Figure S3:** The proportion of the population of an LTLA that is Black British/African/Caribbean, as given in the 2019 ONS report [5]. Boundary source: Office for National Statistics licensed under the Open Government Licence v.3.0 [12]. Contains OS data © Crown copyright and database right [2024].

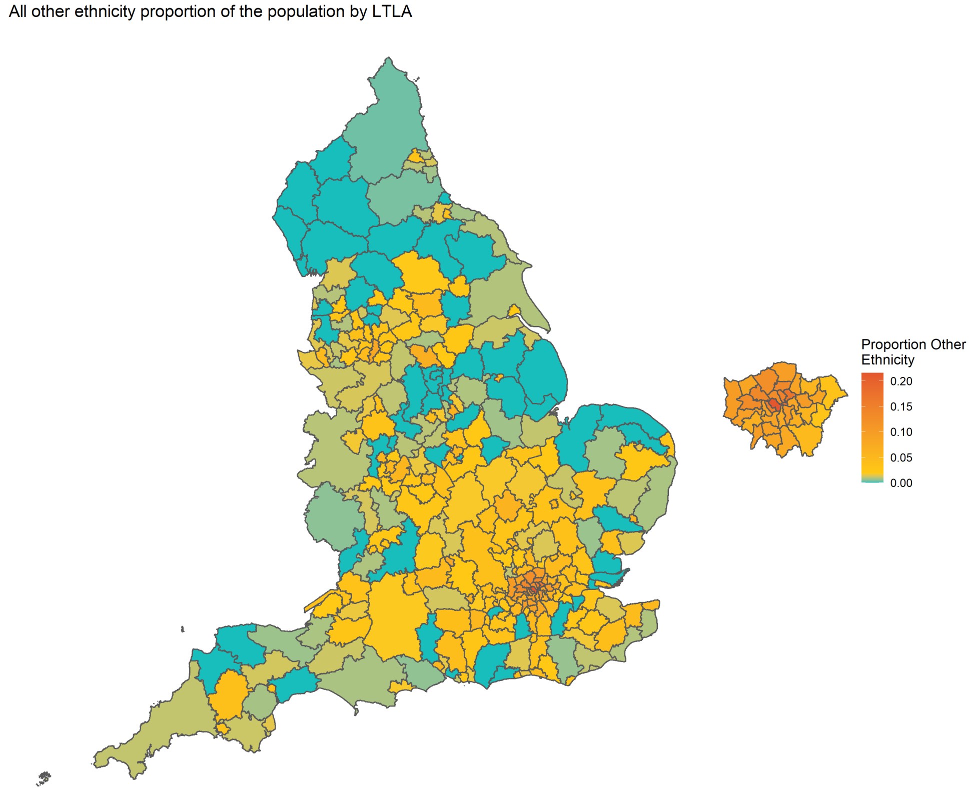

**Figure S4:** The proportion of the population of an LTLA that is "Mixed Multiple" or "Other" ethnicity, as given in the 2019 ONS report [5]. Boundary source: Office for National Statistics licensed under the Open Government Licence v.3.0 [12]. Contains OS data © Crown copyright and database right [2024].

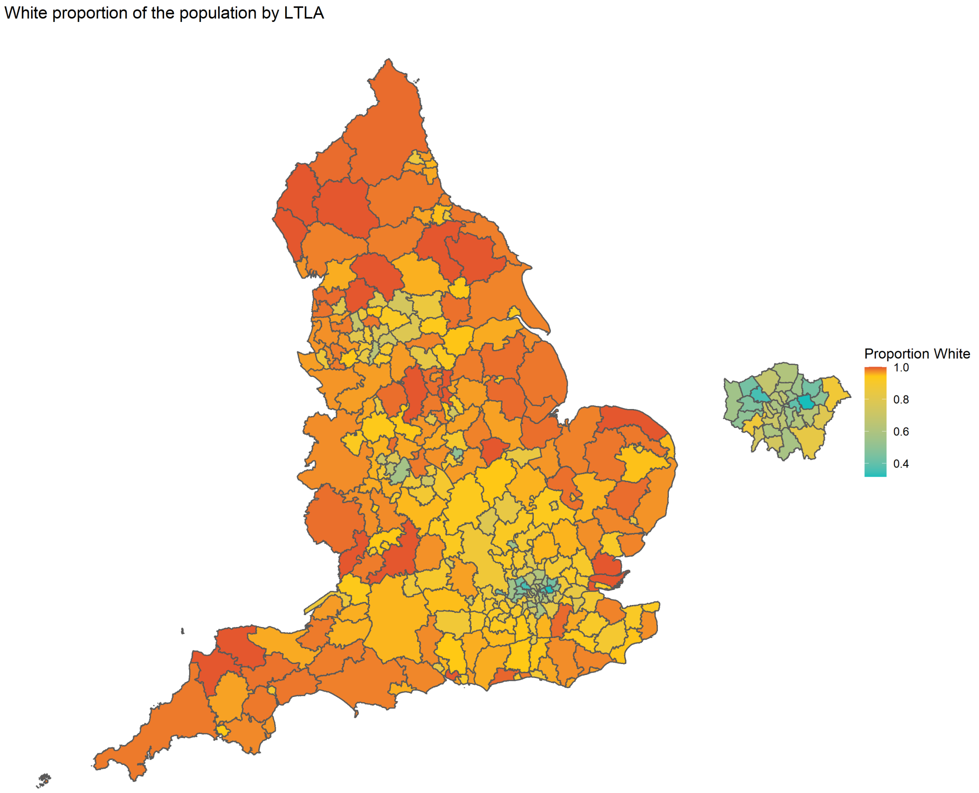

**Figure S5:** The proportion of the population of an LTLA that is White, as given in the 2019 ONS report [5]. Boundary source: Office for National Statistics licensed under the Open Government Licence v.3.0 [12]. Contains OS data © Crown copyright and database right [2024].

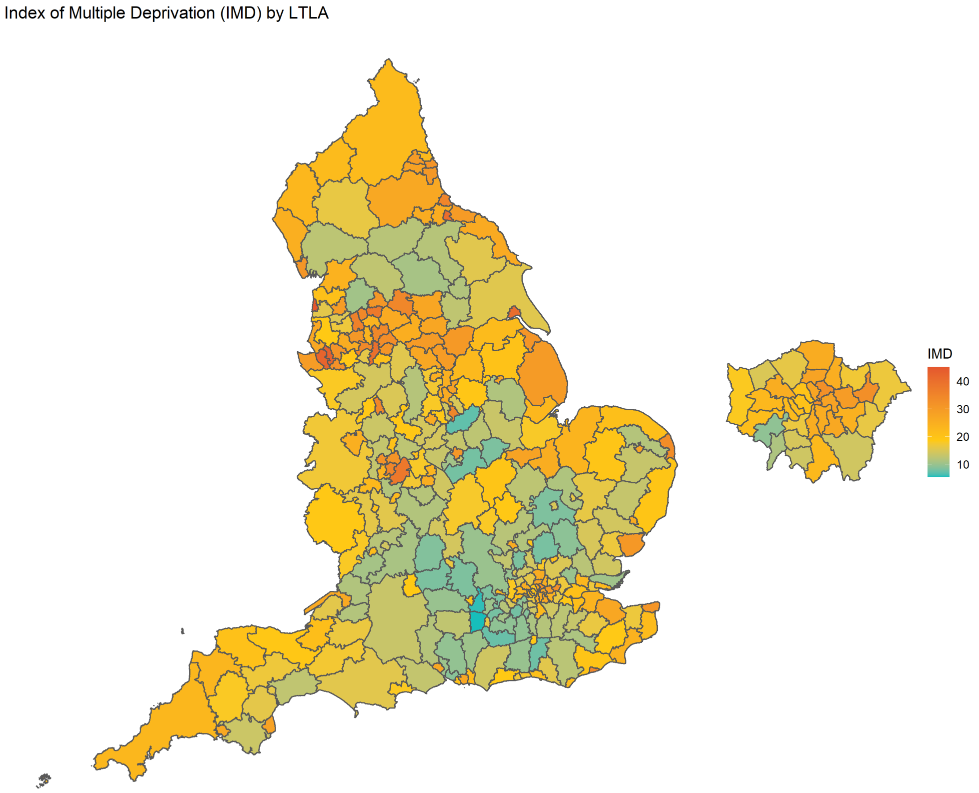

**Figure S6:** The average Index of Multiple Deprivation (IMD) score by LTLA, as given in [6]. Boundary source: Office for National Statistics licensed under the Open Government Licence v.3.0 [12]. Contains OS data © Crown copyright and database right [2024].

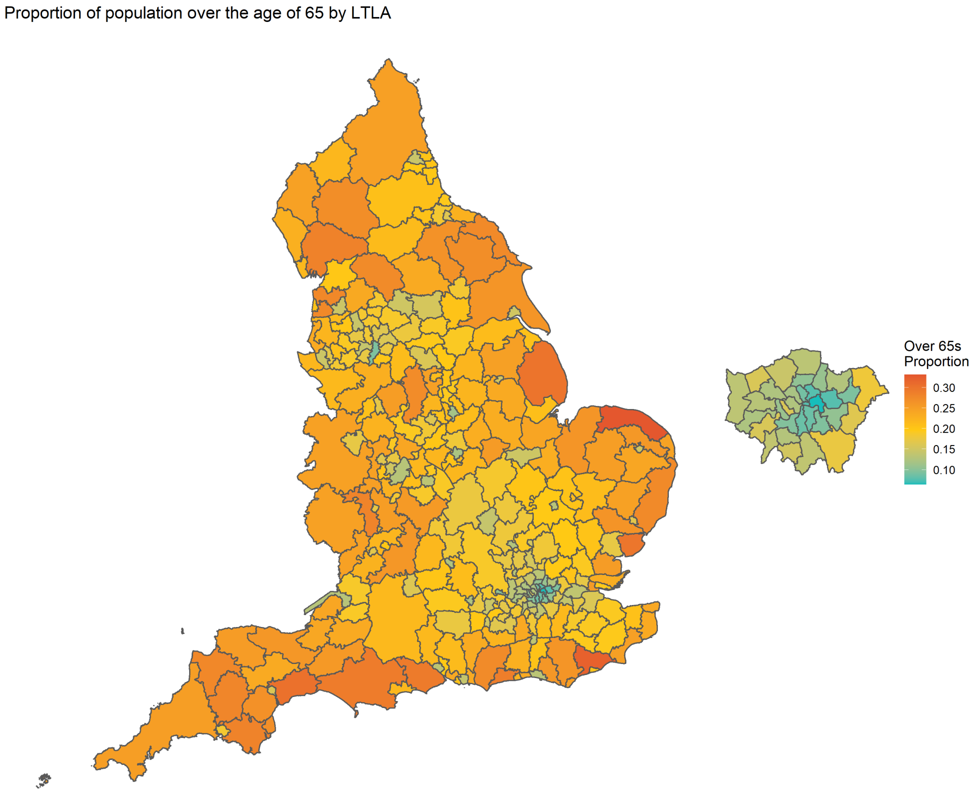

**Figure S7:** The proportion of an LTLA's population over the age of 65, as given in [3]. Boundary source: Office for National Statistics licensed under the Open Government Licence v.3.0 [12]. Contains OS data © Crown copyright and database right [2024].

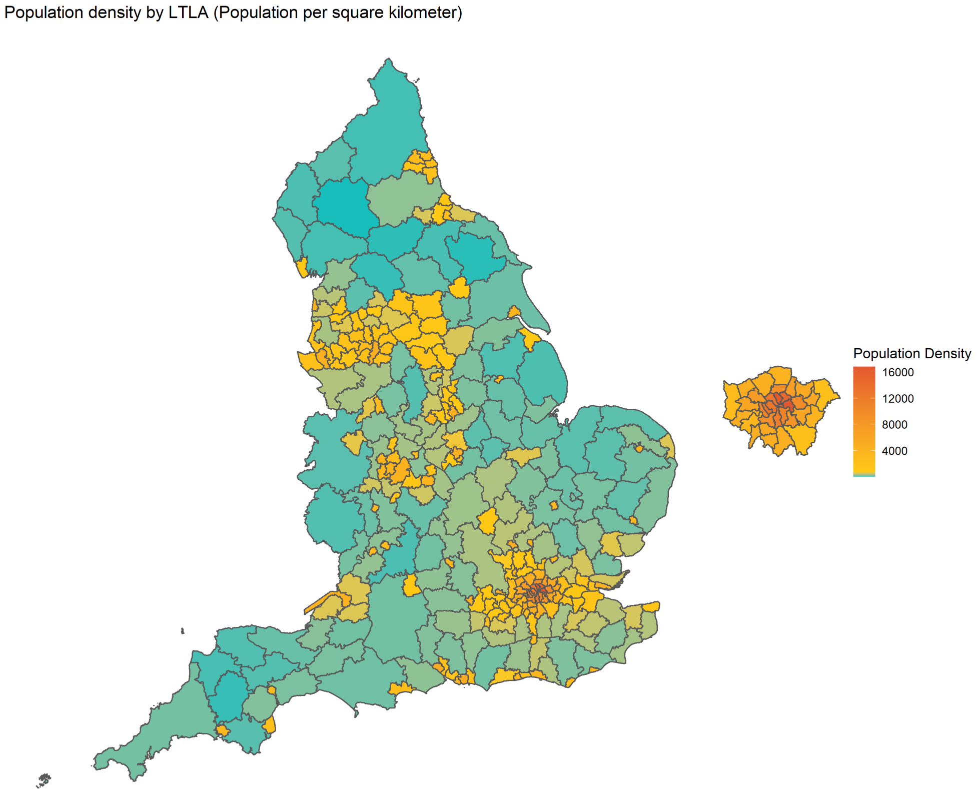

**Figure S8:** The population density of each LTLA, in people per km$^2$, as given in [3]. Boundary source: Office for National Statistics licensed under the Open Government Licence v.3.0 [12]. Contains OS data © Crown copyright and database right [2024].

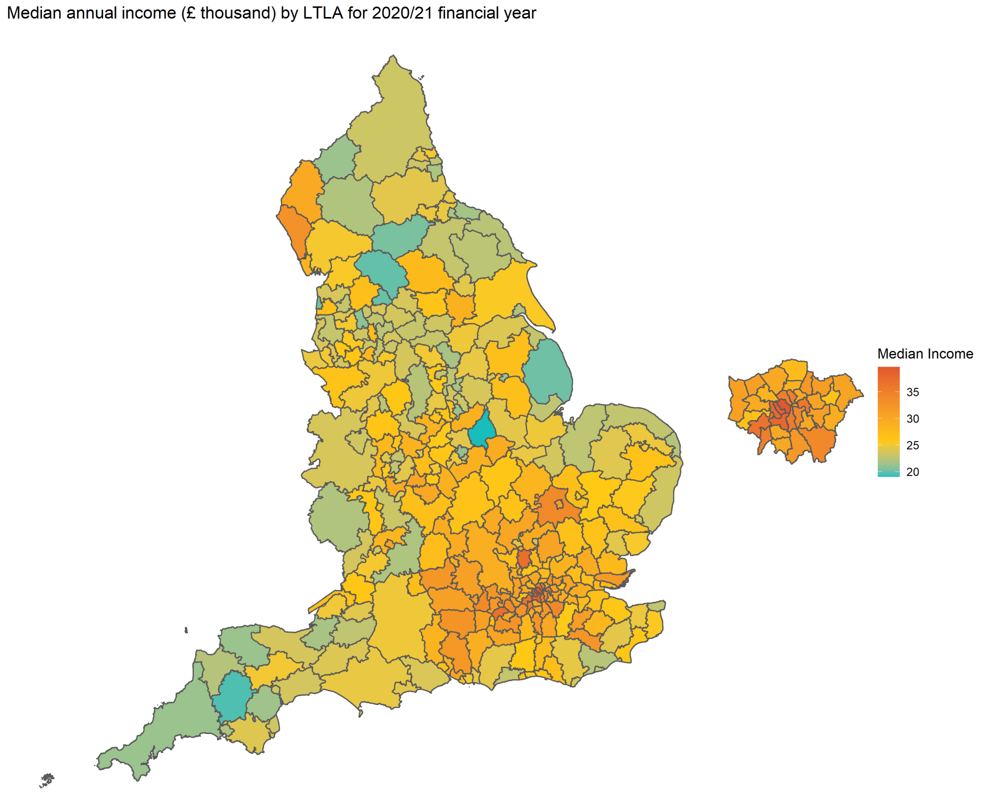

**Figure S9:** The median annual income of each LTLA, in the 2020/21 financial year, as given in [7]. Boundary source: Office for National Statistics licensed under the Open Government Licence v.3.0 [12]. Contains OS data © Crown copyright and database right [2024].

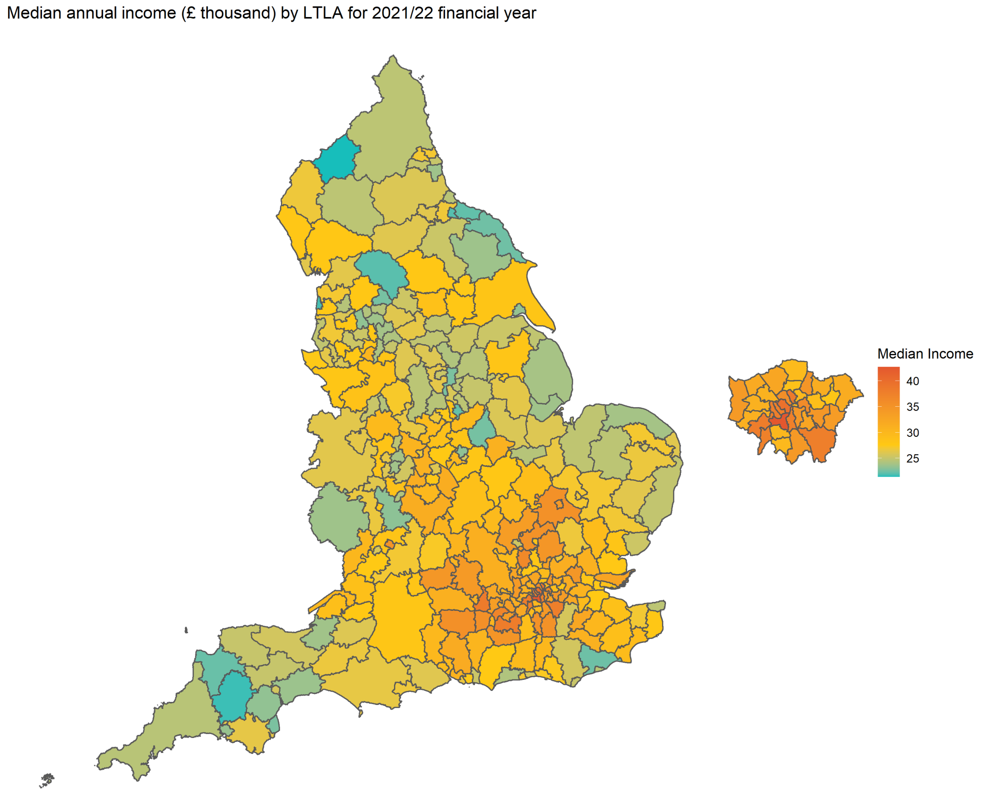

**Figure S10:** The median annual income of each LTLA, in the 2021/22 financial year, as given in [7]. Boundary source: Office for National Statistics licensed under the Open Government Licence v.3.0 [12]. Contains OS data © Crown copyright and database right [2024].

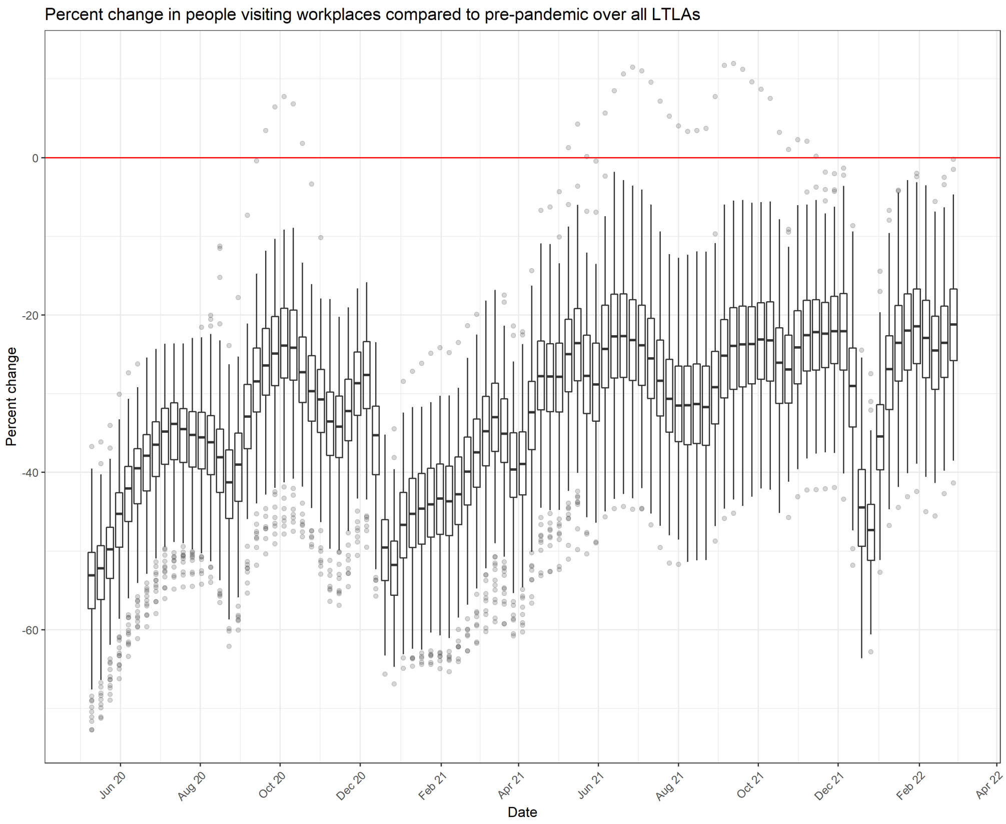

**Figure S11:** The variation between LTLAs in the percentage difference of people visiting workplaces compared to pre-pandemic. A boxplot is given for each week showing the spread of values across the 306 LTLAs considered. Data taken from Google community mobility reports [8].

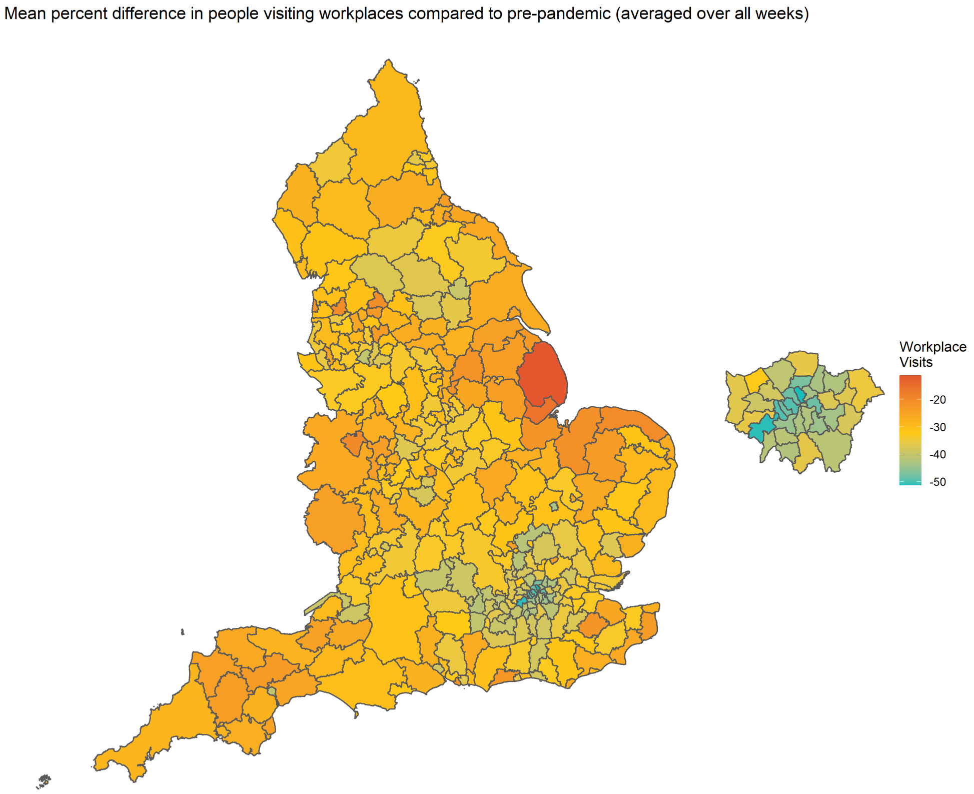

**Figure S12:** The mean percentage difference in the number of people visiting workplaces compared to pre-pandemic by LTLA, averaged over all weeks considered. Data taken from Google community mobility reports [8]. Boundary source: Office for National Statistics licensed under the Open Government Licence v.3.0 [12]. Contains OS data © Crown copyright and database right [2024].

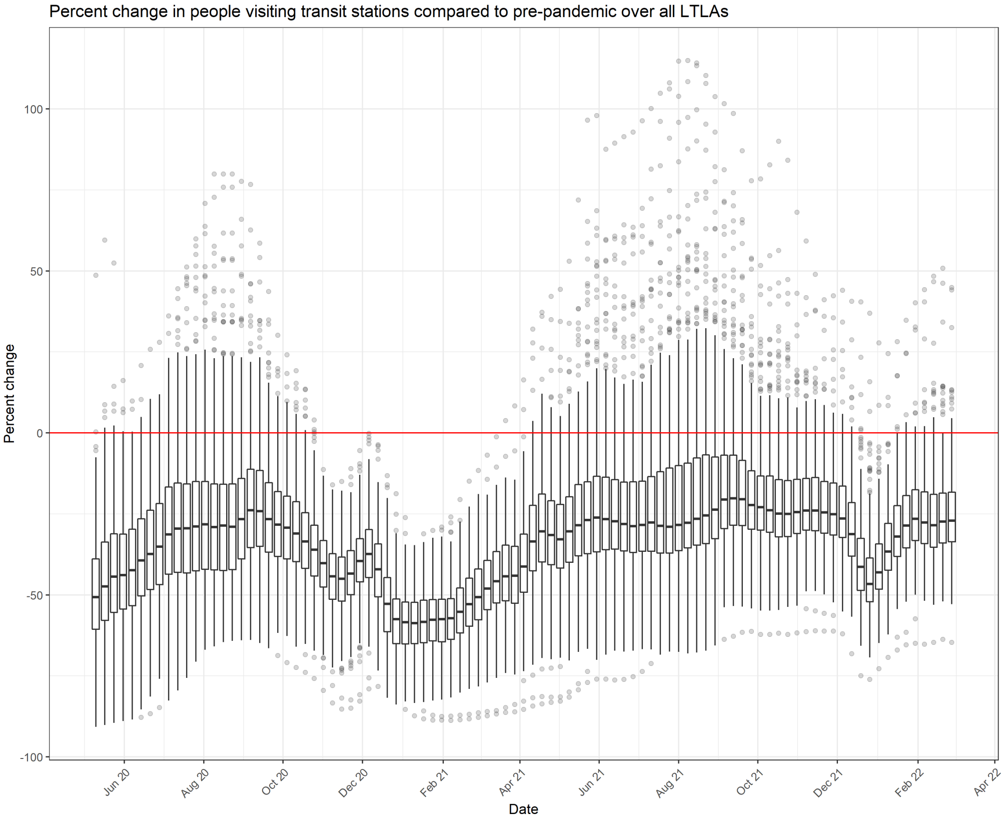

**Figure S13:** The variation between LTLAs in the percentage difference of people visiting transit stations compared to pre-pandemic. A boxplot is given for each week showing the spread of values across the 306 LTLAs considered. Data taken from Google community mobility reports [8].

Mean percent difference in people visiting transit stations compared to pre-pandemic (averaged over all weeks)

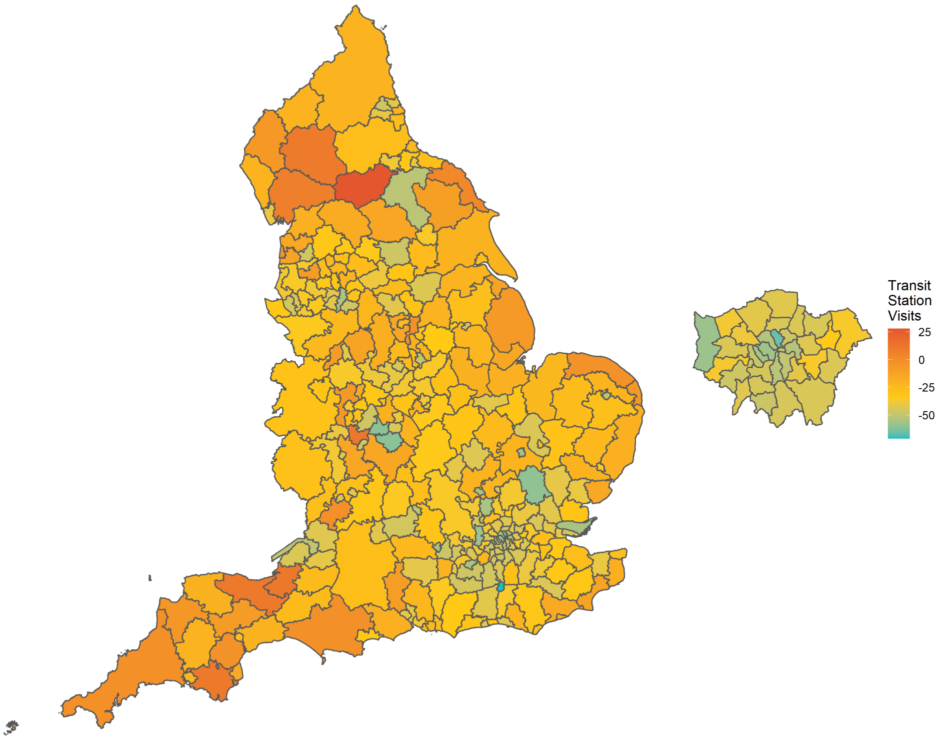

**Figure S14:** The mean percentage difference in the number of people visiting transit stations compared to pre-pandemic by LTLA, averaged over all weeks considered. Data taken from Google community mobility reports [8]. Boundary source: Office for National Statistics licensed under the Open Government Licence v.3.0 [12]. Contains OS data © Crown copyright and database right [2024].

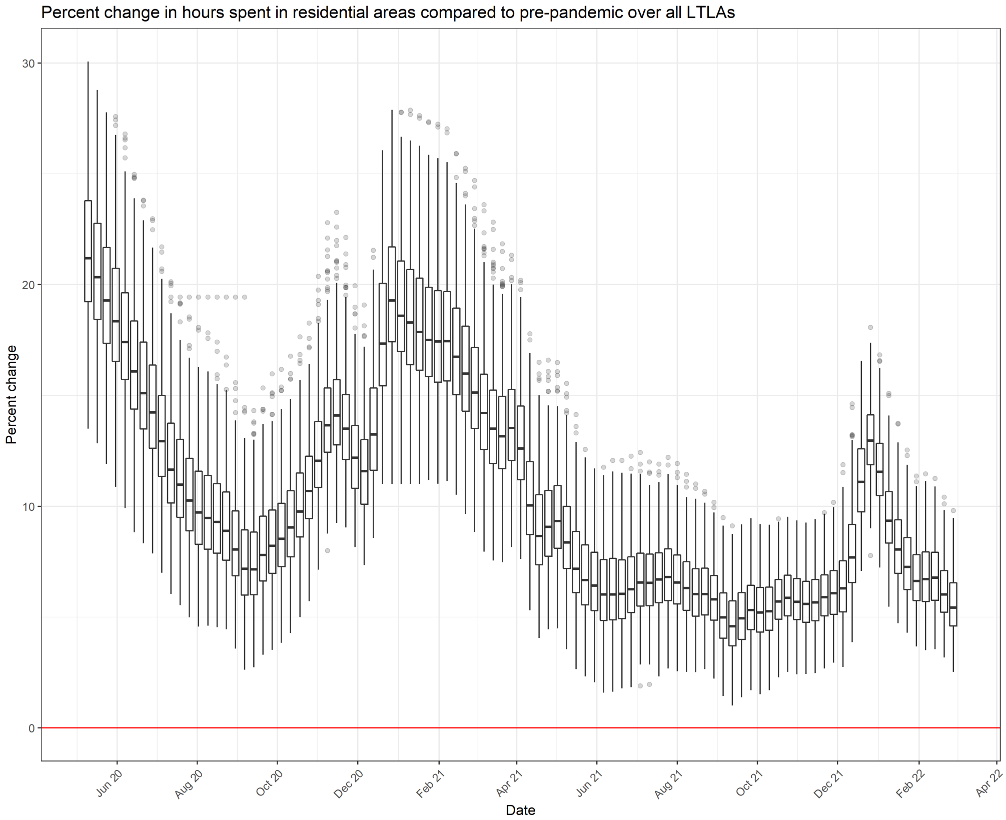

**Figure S15:** The variation between LTLAs in the percentage difference of hours spent in residential areas compared to pre-pandemic. A boxplot is given for each week showing the spread of values across the 306 LTLAs considered. Data taken from Google community mobility reports [8].

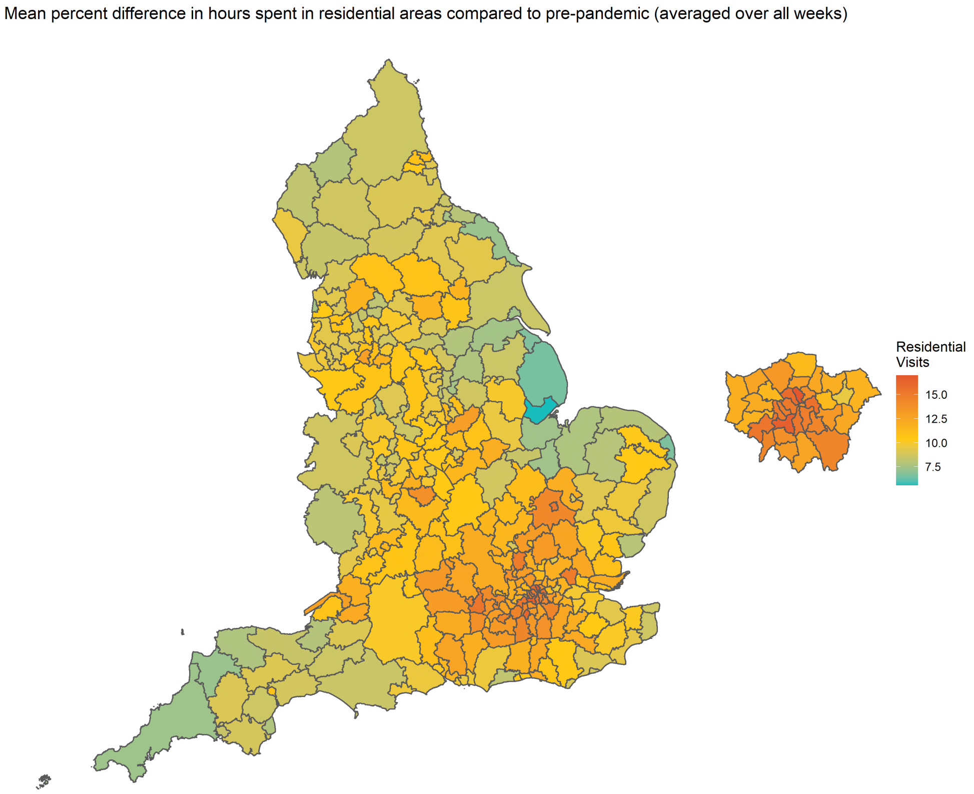

**Figure S16:** The mean percentage difference in the hours people spent in residential areas compared to pre-pandemic by LTLA, averaged over all weeks considered. Data taken from Google community mobility reports [8]. Boundary source: Office for National Statistics licensed under the Open Government Licence v.3.0 [12]. Contains OS data © Crown copyright and database right [2024].

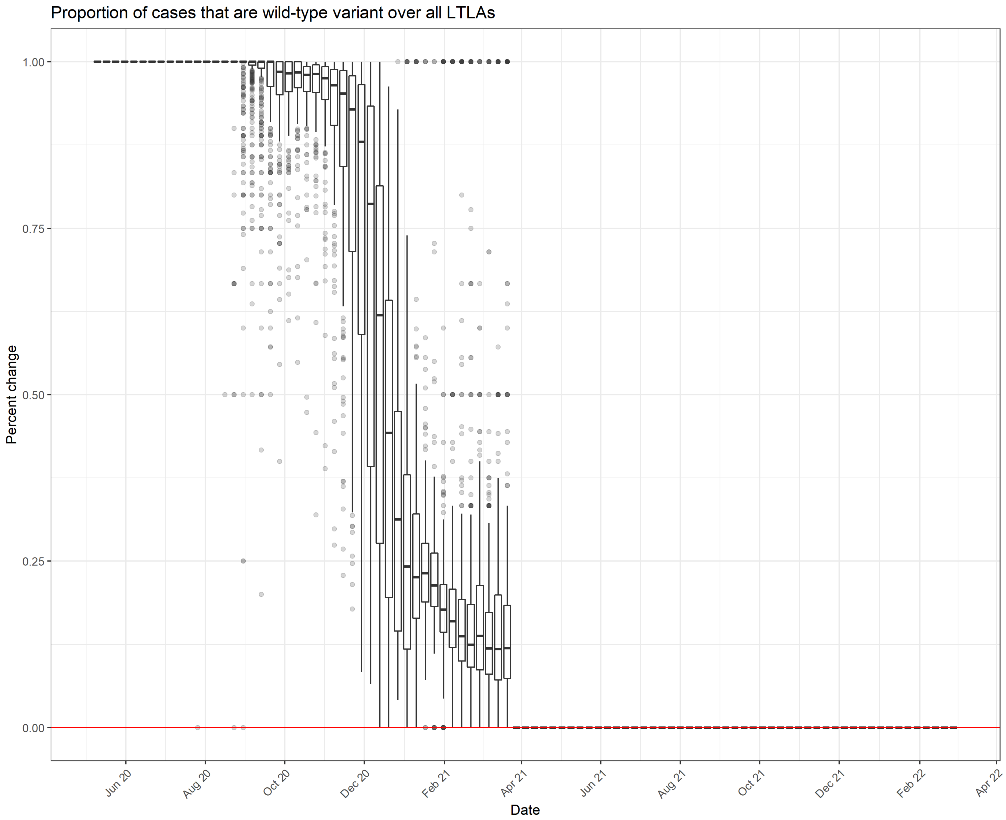

**Figure S17:** The variation between LTLAs in the proportion of cases that are wild-type. A boxplot is given for each week showing the spread of values across the 306 LTLAs considered. Variant type is defined by S-gene target failure (SGTF), as reported through the UKHSA line list.

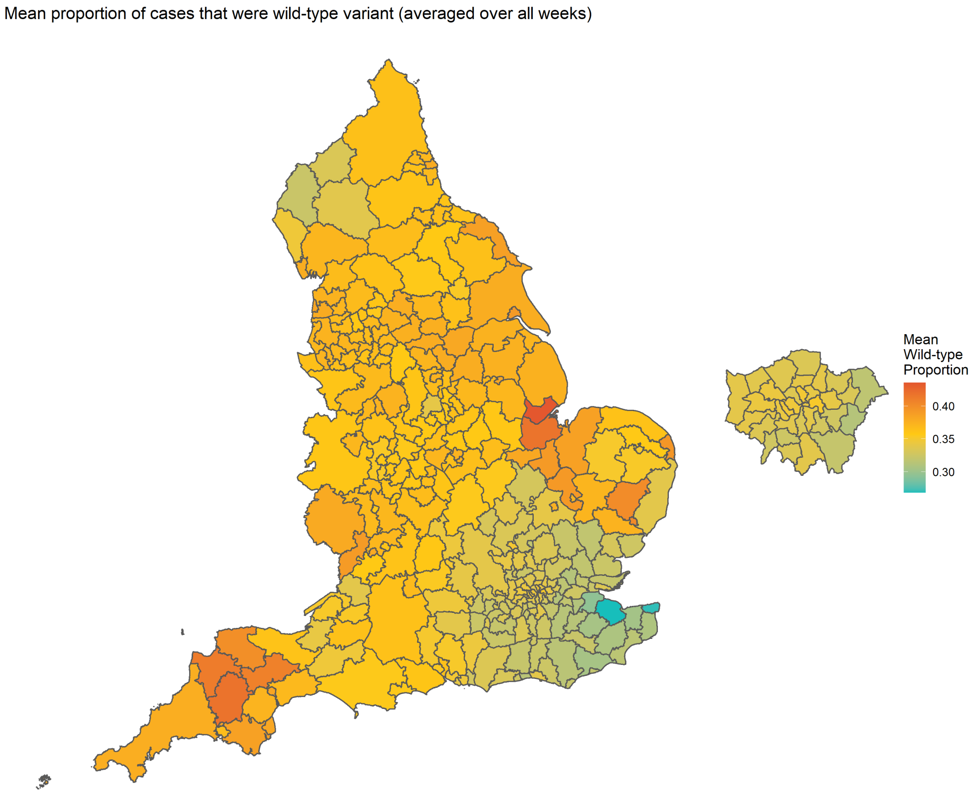

**Figure S18:** The mean proportion of cases that are defined as wild-type by LTLA, averaged over all weeks considered. Variant type is defined by S-gene target failure (SGTF), as reported through the UKHSA line list. Boundary source: Office for National Statistics licensed under the Open Government Licence v.3.0 [12]. Contains OS data © Crown copyright and database right [2024].

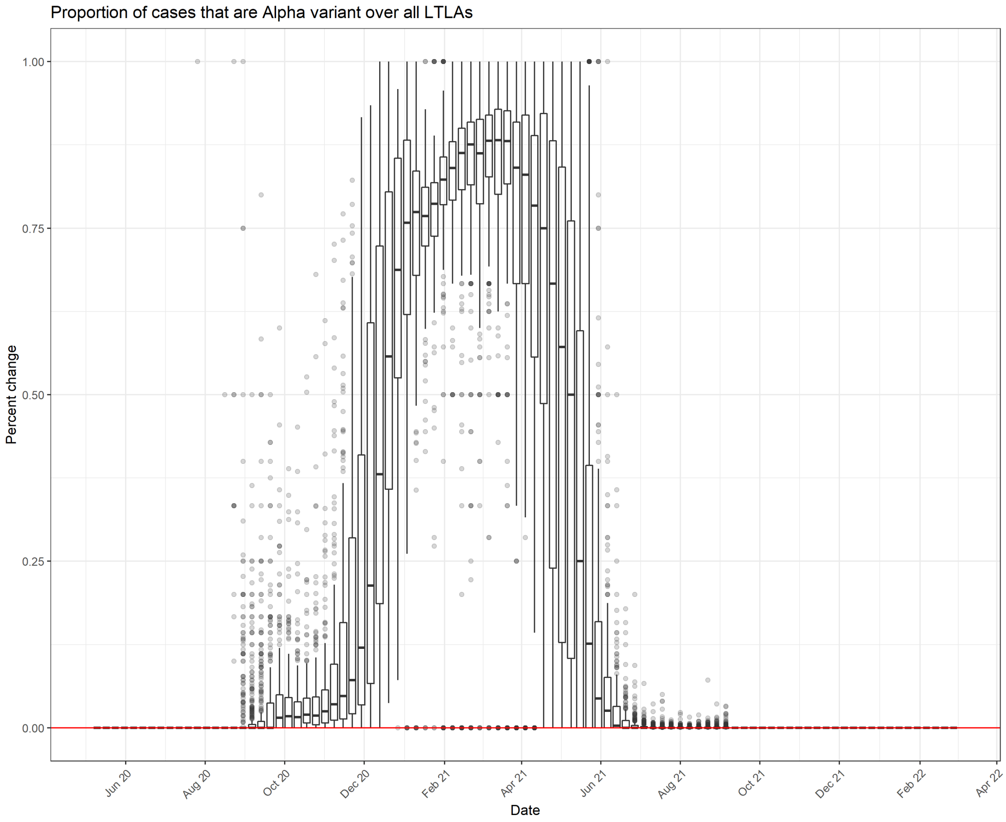

**Figure S19:** The variation between LTLAs in the proportion of cases that are Alpha variant. A boxplot is given for each week showing the spread of values across the 306 LTLAs considered. Variant type is defined by S-gene target failure (SGTF), as reported through the UKHSA line list.

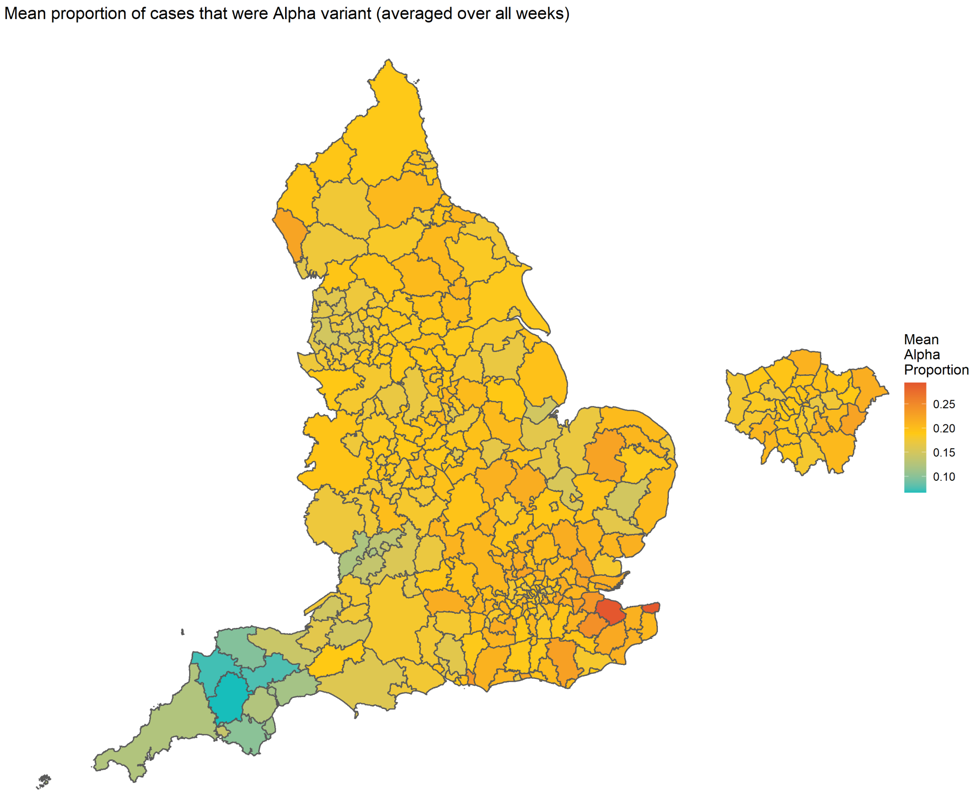

**Figure S20:** The mean proportion of cases that are defined as Alpha variant by LTLA, averaged over all weeks considered. Variant type is defined by S-gene target failure (SGTF), as reported through the UKHSA line list. Boundary source: Office for National Statistics licensed under the Open Government Licence v.3.0 [12]. Contains OS data © Crown copyright and database right [2024].

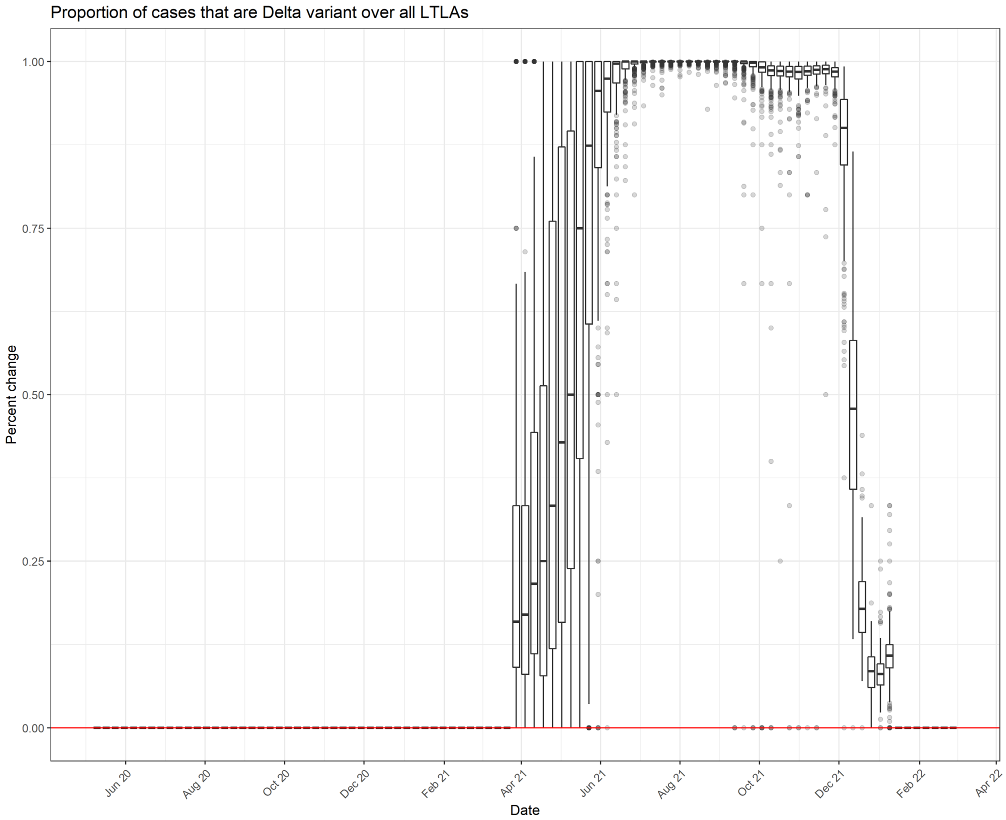

**Figure S21:** The variation between LTLAs in the proportion of cases that are Delta variant. A boxplot is given for each week showing the spread of values across the 306 LTLAs considered. Variant type is defined by S-gene target failure (SGTF), as reported through the UKHSA line list.

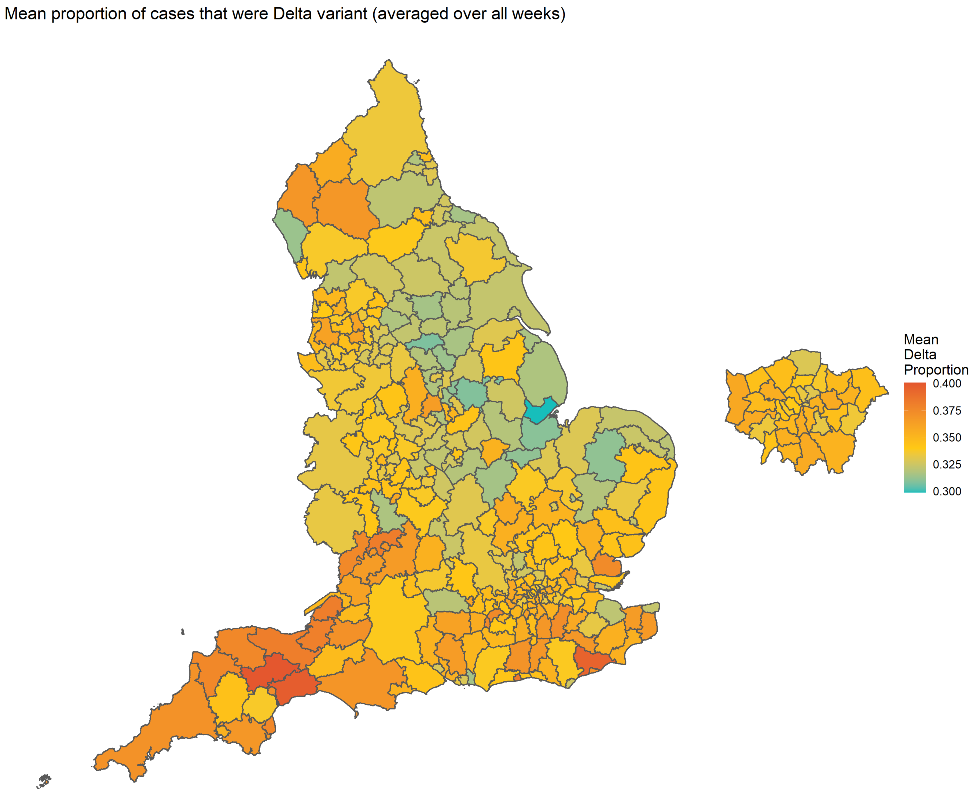

**Figure S22:** The mean proportion of cases that are defined as Delta variant by LTLA, averaged over all weeks considered. Variant type is defined by S-gene target failure (SGTF), as reported through the UKHSA line list. Boundary source: Office for National Statistics licensed under the Open Government Licence v.3.0 [12]. Contains OS data © Crown copyright and database right [2024].

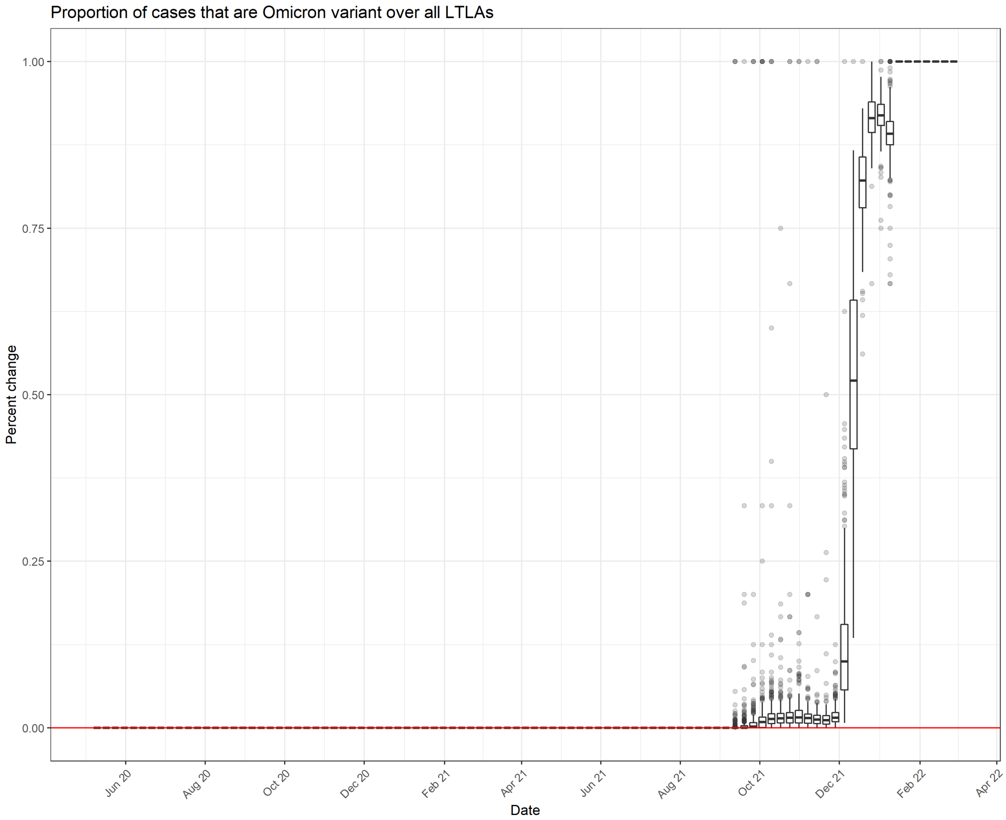

**Figure S23:** The variation between LTLAs in the proportion of cases that are Omicron variant. A boxplot is given for each week showing the spread of values across the 306 LTLAs considered. Variant type is defined by S-gene target failure (SGTF), as reported through the UKHSA line list.

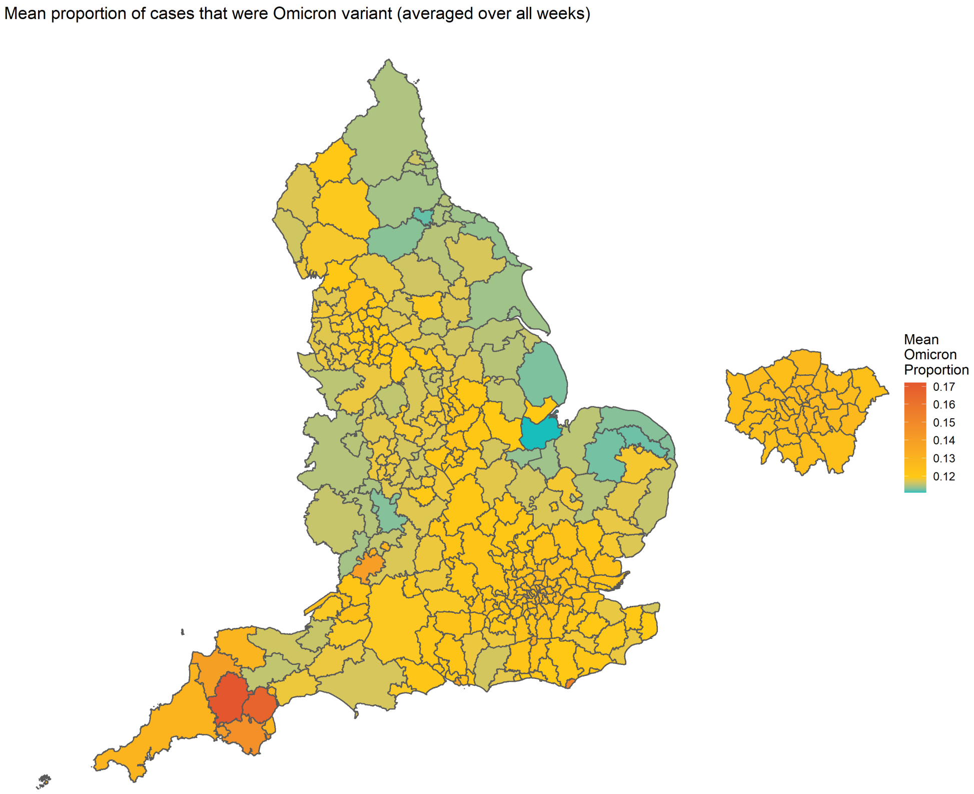

**Figure S24:** The mean proportion of cases that are defined as Omicron variant by LTLA, averaged over all weeks considered. Variant type is defined by S-gene target failure (SGTF), as reported through the UKHSA line list. Boundary source: Office for National Statistics licensed under the Open Government Licence v.3.0 [12]. Contains OS data © Crown copyright and database right [2024].

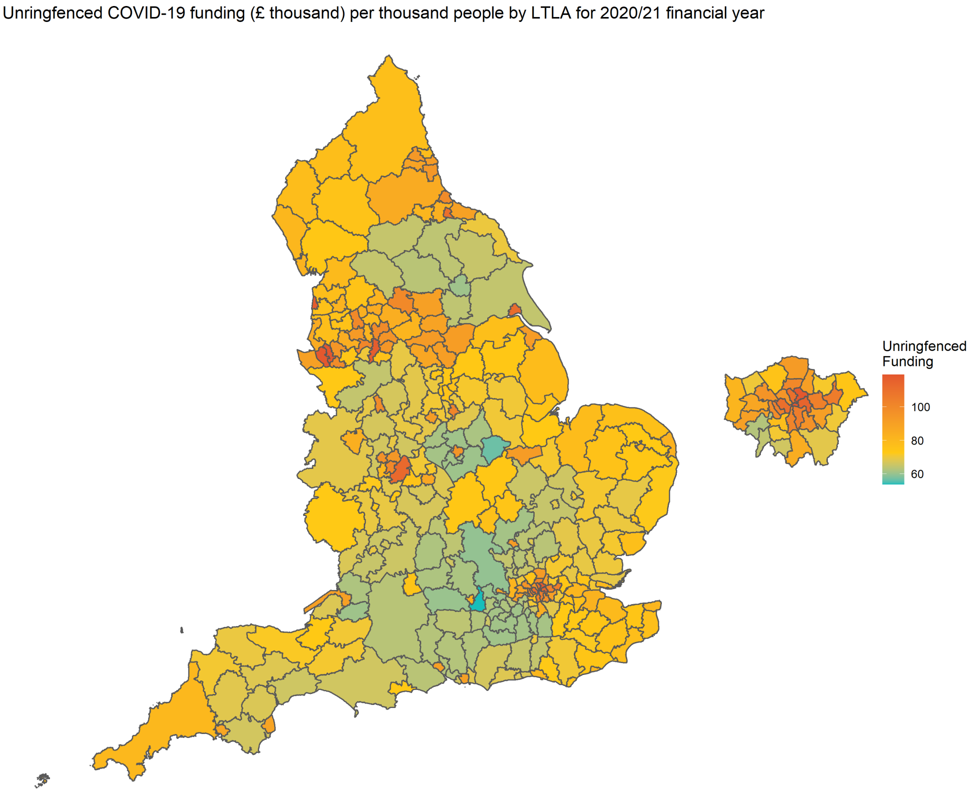

**Figure S25:** The amount of unringfenced funding provided per thousand people in the 2020/21 financial year by LTLA. Data reported at [9]. Boundary source: Office for National Statistics licensed under the Open Government Licence v.3.0 [12]. Contains OS data © Crown copyright and database right [2024].

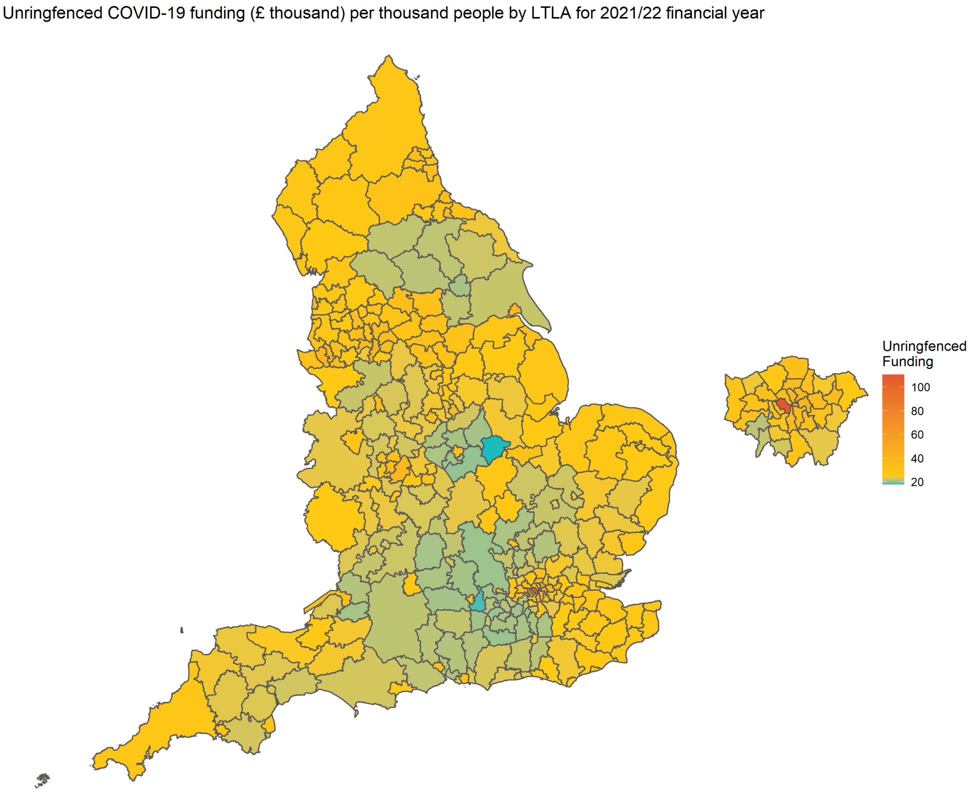

**Figure S26:** The amount of unringfenced funding provided per thousand people in the 2021/22 financial year by LTLA. Data reported at [9]. Boundary source: Office for National Statistics licensed under the Open Government Licence v.3.0 [12]. Contains OS data © Crown copyright and database right [2024].

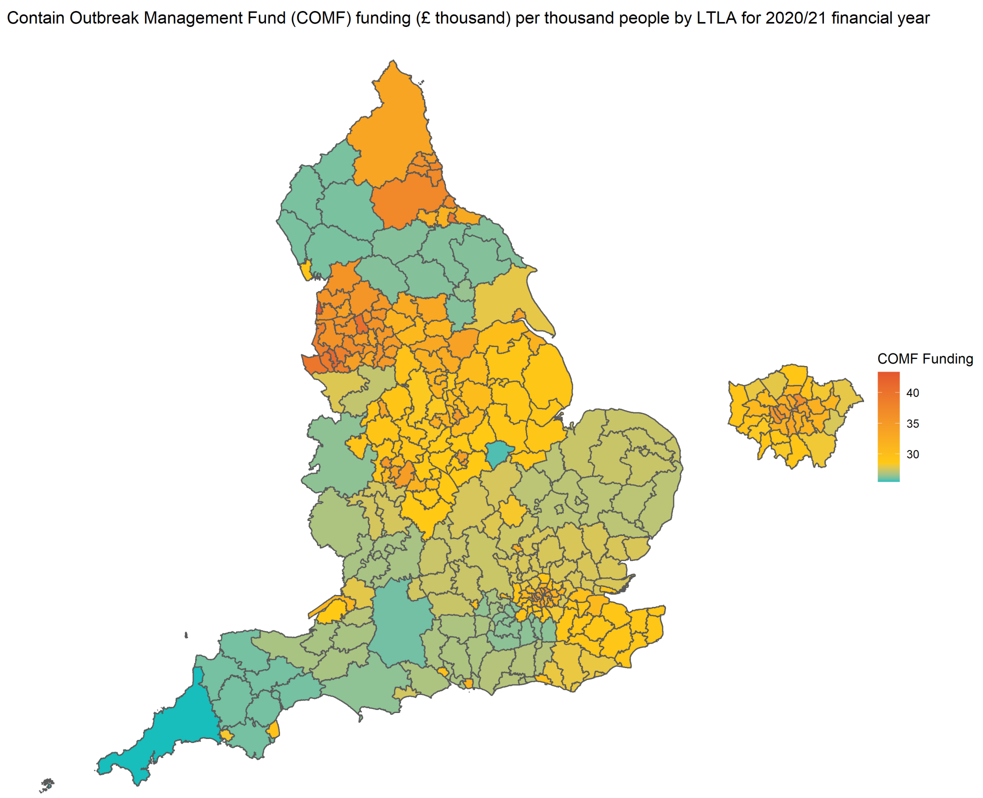

**Figure S27:** The amount of Contain Outbreak Management Fund (COMF) funding provided per thousand people in the 2020/21 financial year by LTLA. Data reported at [9]. Boundary source: Office for National Statistics licensed under the Open Government Licence v.3.0 [12]. Contains OS data © Crown copyright and database right [2024].

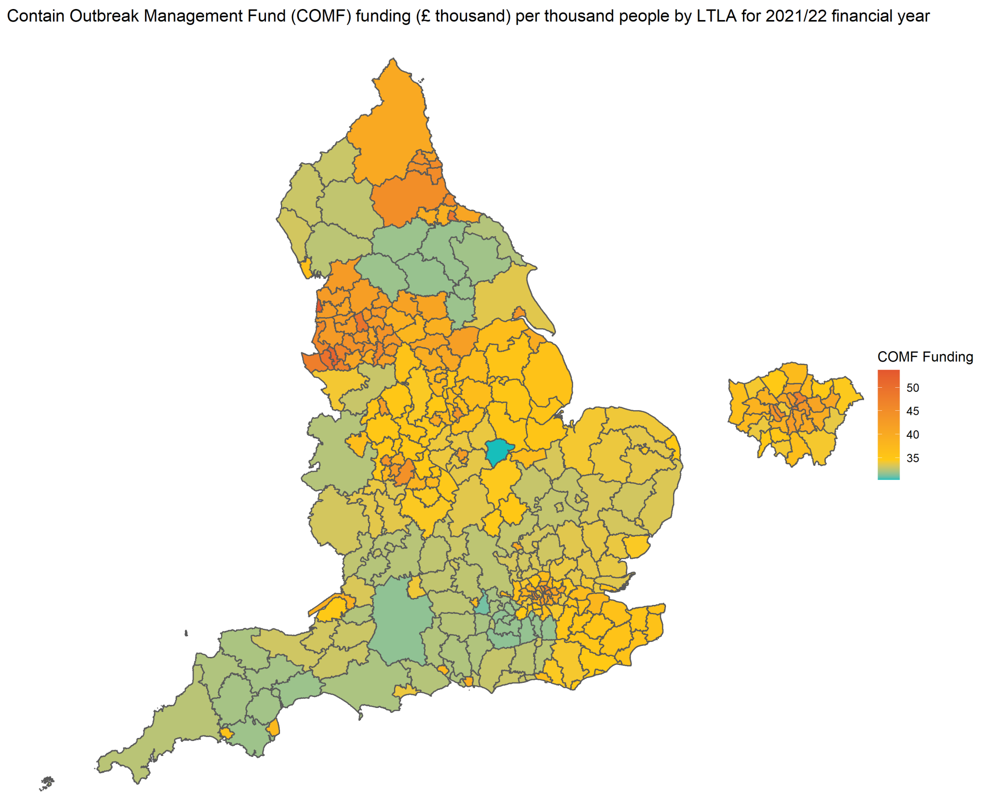

**Figure S28:** The amount of Contain Outbreak Management Fund (COMF) funding provided per thousand people in the 2021/22 financial year by LTLA. Data reported at [9]. Boundary source: Office for National Statistics licensed under the Open Government Licence v.3.0 [12]. Contains OS data © Crown copyright and database right [2024].

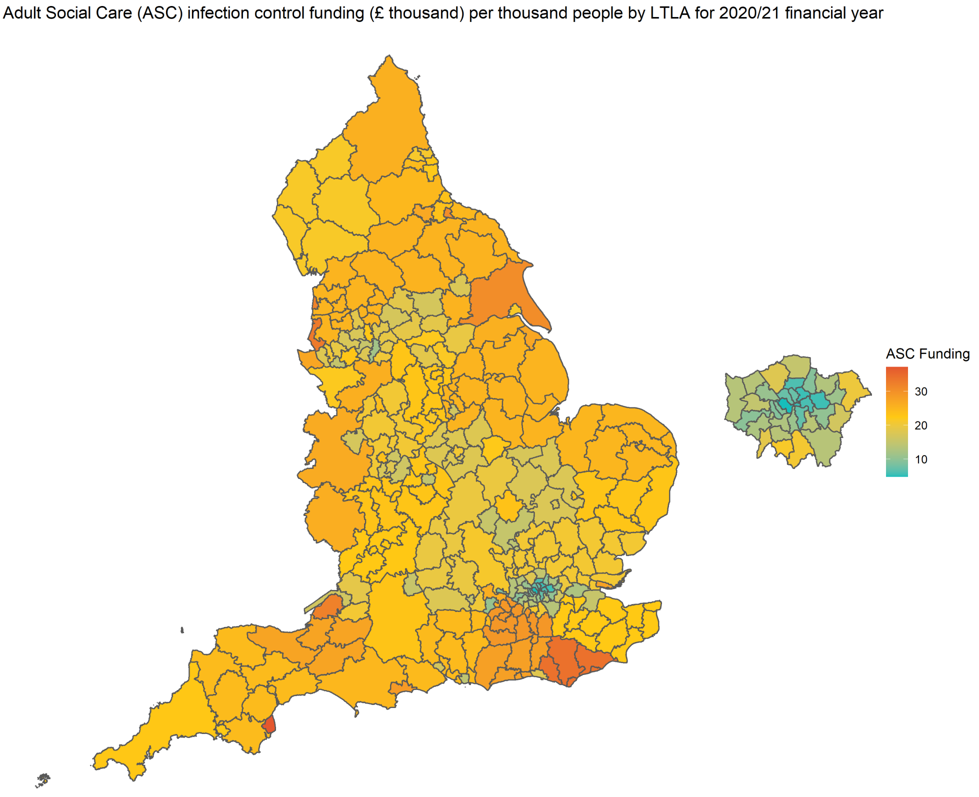

**Figure S29:** The amount of Adult Social Care (ASC) infection control funding provided per thousand people in the 2020/21 financial year by LTLA. Data reported at [9]. Boundary source: Office for National Statistics licensed under the Open Government Licence v.3.0 [12]. Contains OS data © Crown copyright and database right [2024].

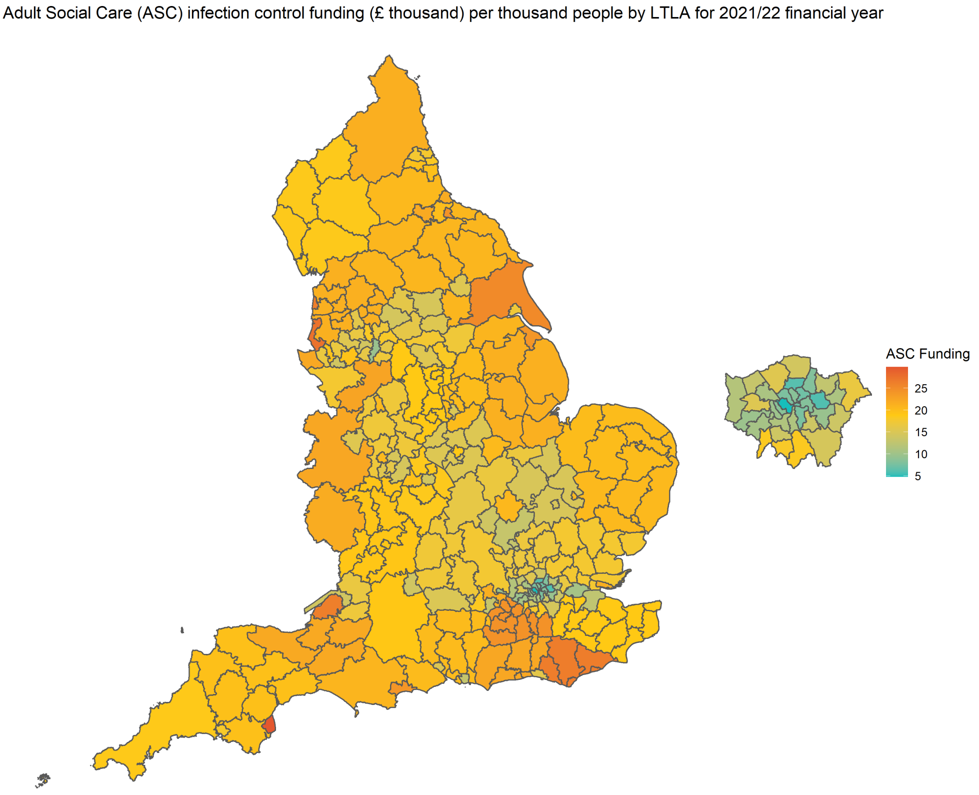

**Figure S30:** The amount of Adult Social Care (ASC) infection control funding provided per thousand people in the 2021/22 financial year by LTLA. Data reported at [9]. Boundary source: Office for National Statistics licensed under the Open Government Licence v.3.0 [12]. Contains OS data © Crown copyright and database right [2024].

#### 1.7 Variable autocorrelation

Naturally, many of the variables we've introduced above correlate with one another, in that the spatial patterns observed in the above heatmaps may look similar for some variables. Below in Figure S31 we present a paired correlation plot. We remove the VOC variables, and include only the proportion White of the ethnicity variables. As these ethnicity proportions sum to 1 they are naturally negatively correlated. The main diagonal of Figure S31 shows the density plot of variables, the upper diagonal shows the overall Pearson correlation coefficient (black) and the correlation coefficient for different financial years. The triple asterisks indicates that coefficients are statistically significant (p value $< 0.001$).

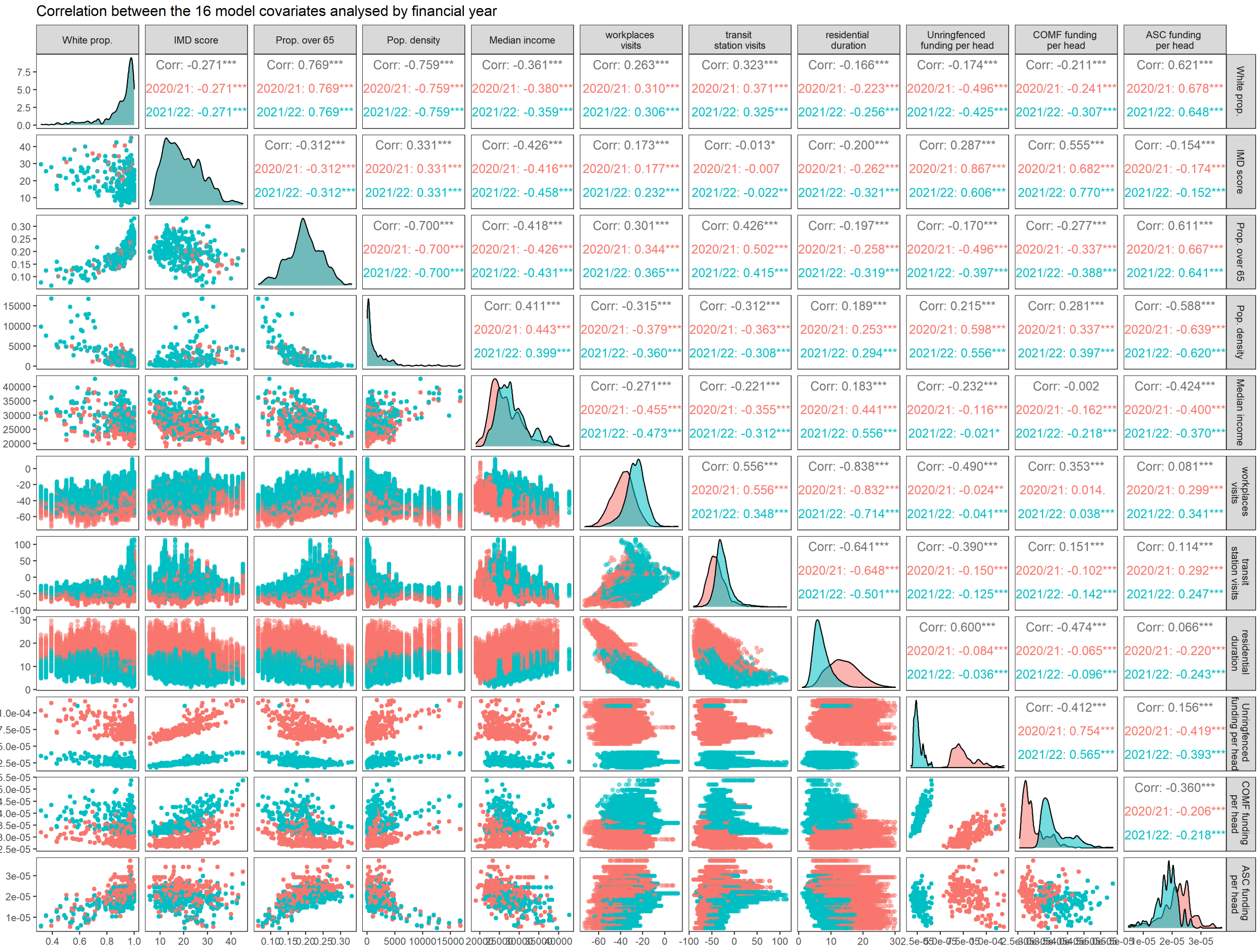

**Figure S31:** A paired correlation plot for model variables for separate financial years. Red is data before April 6th 2021, blue is data after April 6th 2021. Upper diagonal gives Pearson correlation coefficients. Asterisks denote whether the value is statistically significant from 0 (*** $p < 0.001$, ** $p < 0.01$, * $p < 0.05$, . $p < 0.1$). Lower diagonal shows the paired data plots.

We also show the paired correlation plots for the ethnicity proportion variables below in Figure S32.

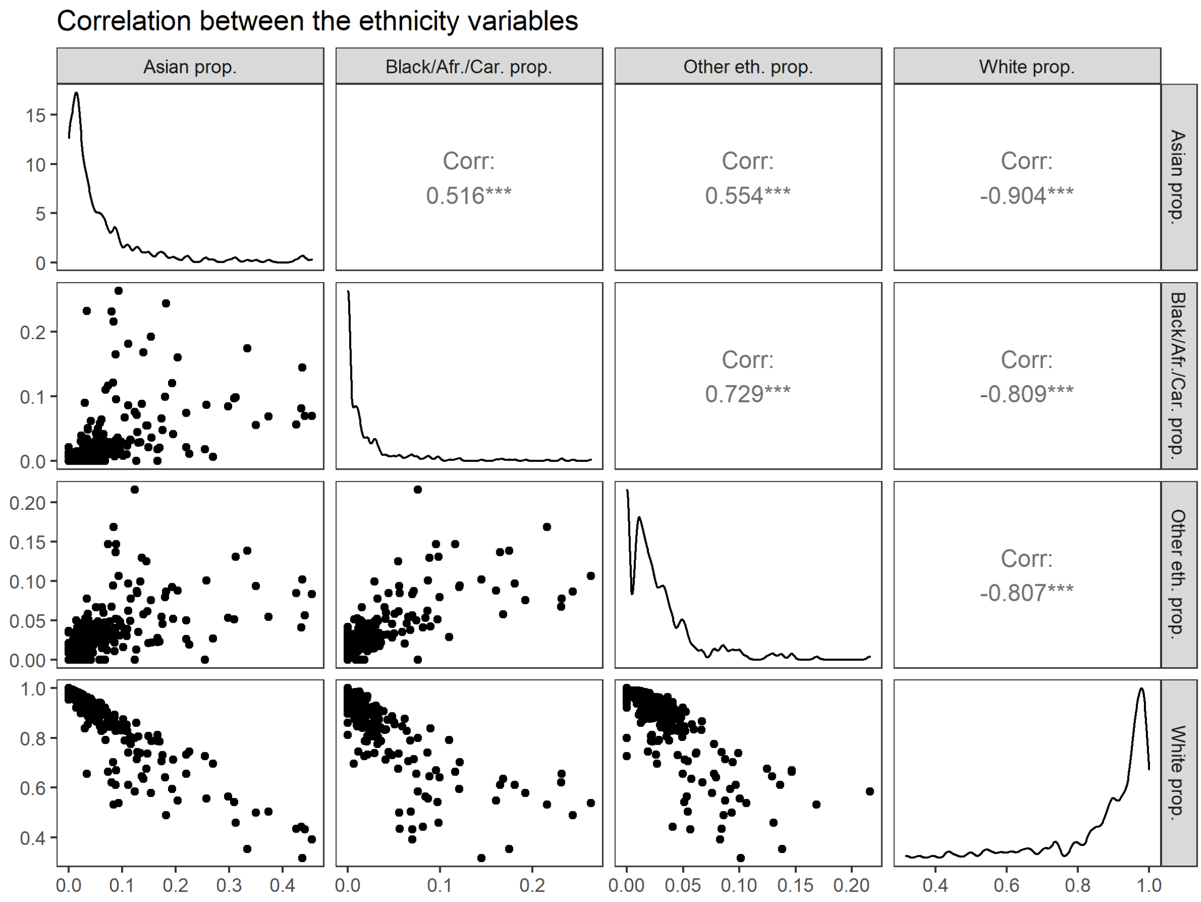

**Figure S32:** A paired correlation plot for proportion ethnicity model variables. Upper diagonal gives Pearson correlation coefficients. Asterisks denote whether the value is statistically significant from 0 (*** $p < 0.001$). Lower diagonal shows the paired data plots.

#### 1.8 Heterogeneity within LTLAs

LTLAs are based upon administrative boundaries, and as such can differ greatly in population size. Figure S33 below shows the distribution of populations across the 306 LTLAs considered. The most populous LTLA is Birmingham - 1,140,525 people, and the least populous is Rutland - 40,476 people. As would be expected, heterogeneity within LTLA can be expected at this scale. The IMD data used [6], provides IMD calculations for as fine as Lower layer Super Output Areas (LSOAs) scale, consisting of 32,844 regions in England. LSOAs broadly range from population sizes of 1,000 to 2,500. Many of the larger LTLAs considered can be seen to contain LSOAs of both high IMD and low IMD within their bounds - a key caveat to take into account. Many variables of interest however are not available at finer spatial level, and the COVID-19 case count data becomes predominantly zero-count data at finer spatial levels, pushing the bounds of modelling feasibilty. As such, the LTLA-scale is considered the most reasonable trade-off between demographic detail and covaraite availability.

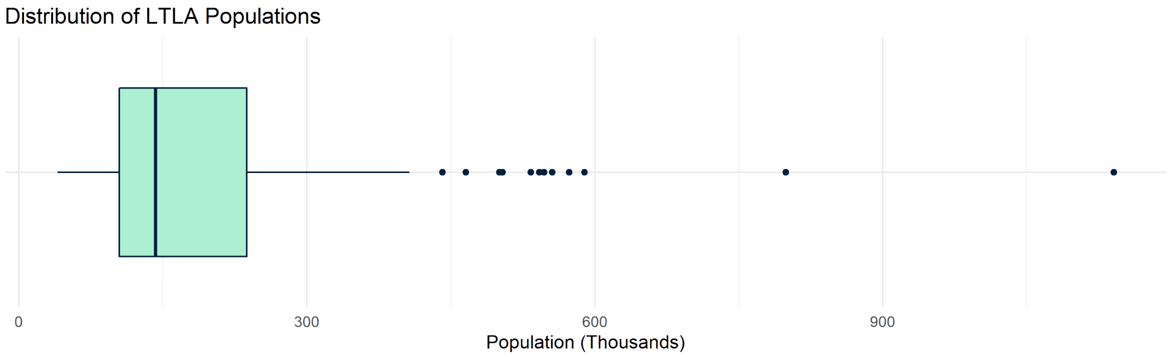

**Figure S33:** A boxplot showing the distribution of LTLA population sizes. Median population is 142,621 (IQR 104,869 – 237,616). Twelve outliers depicted, and their respective population sizes are: Birmingham (1,140,525), Leeds (798,786), Sheffield (589,214), Cornwall and the Isles of Scilly (573,299), Manchester (555,741), Buckinghamshire (547,060), Bradford (542,128), County Durham (533,149), Wiltshire (504,070), Liverpool (500,474), City of Bristol (465,866), and Kirklees (441,290).

### 2 Model Description and Fitting

In this section we explicitly explain the full mathematical model formulation, parameterisation, and fitting procedure.

#### 2.1 Model Definition

Our model is fit to COVID-19 case count data as introduced in Section 1.2. We define $Y_{i,t}$ as the aggregated number of pillar 2, PCR-confirmed COVID-19 cases in LTLA $i$, during week $t$ ($i \in \{1,2,...,306\}$, $t \in \{1,2,...,95\}$). We consider 306 LTLAs, as outlined in Section 1.1, and 95 weeks, running from the week beginning Monday May 10th 2020, to the week beginning Monday February 27th 2022.

Data is assumed to follow a negative binomial distribution. The negative binomial distribution can be expressed for multiple parameter forms, and we express the distribution in the form of a mean ($\mu$) and overdispersion ($\phi$) parameter. Overdispersion arises when the variance observed in data does not linearly scale with the value observed, as is commonly the case for ecological data [13]. The variance of the negative binomial distribution is defined by $\sigma^2 = \mu + \frac{\mu^2}{\phi}$, thus, as $\phi$ gets very large, the distribution converges to a Poisson distribution.

We define our data, $Y_{i,t}$, to be explained by

$$Y_{i,t} \sim \text{NegBinom}(\mu_{i,t}, \phi). \tag{1}$$

Heuristically, we say our mean ($\mu_{i,t}$) number of observed cases takes the form

$$\mu_{i,t} = ((\text{number of cases the previous week}) * (\text{Number of onward infections caused by each case})) \tag{2}$$

The number of onward infections caused by each case is referred to as the effective (time-varying) reproduction number ($R_t$). Our sensitivity analyses presented in Section 4 consider alternate ways of defining (number of cases the previous week) and/or (Number of onward infections caused by each case).

Which cases are likely to be responsible for the observed cases $Y_{i,t}$? We assume the previous week's number of cases in the same LTLA, $Y_{i,t-1}$. We also assume that cases from other LTLAs the previous week may contribute to further infections in LTLA $i$. For our main model analysis, we assume that some proportion of all the cases in LTLAs that share a geographic border with LTLA $i$ will contribute to the subsequent number of observed cases. We also assume that the proportion of cases that "enter" LTLA $i$ varies by LTLA. Thus, we define;

$$\mu_{i,t} = \left( \left( Y_{i,t-1} + \zeta_i \sum_{j \in \Omega_i} Y_{j,t-1} \right) * (\text{Number of onward infections caused by each case}) \right)$$

where $\Omega_i$ is the set of LTLAs who share a geographic boundary with LTLA $i$, and $\zeta_i$ is the proportion of all cases in neighboring LTLAs which "enter" LTLA $i$ to cause onward infections. $\zeta_i \in [0,1]$, where $\zeta_i = 0$ indicates that no cases from outside LTLA $i$ cause further infections inside LTLA $i$, and $\zeta_i = 1$ would indicate that all cases from neighbouring LTLAs cause onward infections inside LTLA $i$.

What parameters or variables do we expect to determine the effective reproduction number at time $t$? This study investigates what impact the LTLA population data, the COVID-19 variants present in an LTLA, and the funding provided to an LTLA (as outlined in Sections 1.3 to 1.5) all have on the effective reproduction number. We also assume there is some background change to the reproduction number over time, affecting all LTLAs, reflecting the impact of population behaviour, vaccine uptake, seasonal variation etc. Thus, we define:

$$\mu_{i,t} = \left( \left( Y_{i,t-1} + \zeta_i \sum_{j \in \Omega_i} Y_{j,t-1} \right) \exp(x_{i,t-1}\beta + z_{t-1} + \theta_i) \right).$$

Here, $x_{i,t-1}$ is a row vector of length 16 containing the values for the sixteen variables listed in Section 1.6 for LTLA $i$ at week $t-1$. $\beta$ is a column vector of length 16 of the coefficients for each variable in $x_{i,t-1}$, denoting the strength of their respective impacts. Therefore $x_{i,t-1}\beta$, following vector multiplication rules, is a single value capturing the impact of the sixteen variables.

We apply feature scaling, meaning each of the sixteen data categories (except for the variant proportion data, which is already between 0 and 1) is standardised by subtracting the mean of the data category and dividing by the standard deviation of the data category. This ensures that the values of $\beta$ can be directly compared, and is not scaled respective to the original units of the data.

$\theta_i$ is an LTLA-specific error term, capturing any background variation in the reproduction number across LTLAs for all weeks that is not already captured by $x_{i,t-1}$.

$z_{t-1}$ is the $(t-1)^{\text{th}}$ step in a Gaussian random walk process, capturing the background time-varying changes to the reproduction number seen across all LTLAs. By definition, we model a random variable $X_t \sim \mathcal{N}(0,Q)$, where $Q$ is a model parameter, and $z_t = \sum_{j=1}^{t} X_j$.

The summation of these terms is then set within an exponential function, to ensure the expression is bounded below by 0.

Lastly, we assume that the population of an LTLA will build up acquired immunity over time. Thus, we assume that only the proportion of the LTLA's population that has not yet been infected is susceptible to infection. We adapt the formulation like so:

$$\mu_{i,t} = \left(\lambda S_{i,t-1}\left(Y_{i,t-1} + \zeta_i \sum_{j\in\Omega_i} Y_{j,t-1}\right) \exp(x_{i,t-1}\beta + z_{t-1} + \theta_i)\right), \tag{3}$$

where $S_{i,t}$ is the proportion of the population of LTLA $i$ that has not yet been infected. We define

$$S_{i,t} = 1 - \frac{\text{Total number of first episodes in LTLA } i \text{ by week } t}{\text{Population of LTLA } i},$$

where the number of first episodes is as defined in Section 1.2. $\lambda$ is a scaling parameter between 0 and 1 to account for the fact that there will be under-reporting of first-episodes.

#### 2.2 Vaccination

Data on vaccination uptake by LTLA is available on the UK data dashboard. However, the variable was not included within the model for several reasons.

Firstly, our model is not able to capture the mechanistic impact of waning vaccine effectiveness [14, 15, 16, 17], vaccine escape features by variant, or differences in effectiveness by vaccine type (AstraZeneca, Pfizer, and Moderna) [18]. Including vaccine uptake would not capture the waning of effectiveness over time or via variant.

Secondly, the vaccine rollout in the UK was regularly adapted in response to the emergence of variants [19] and changes to non-pharmaceutical interventions (NPI) like stay-at-home orders. As such, vaccination correlates heavily and counter-effectively with multiple variables we already include, namely VOC proportion and Google mobility metrics.

Instead, the impact of vaccination is captured within the random walk term $z_t$, alongside changes in behaviour across time, such as in response to public holidays. The gradual downwards trajectory of $z_t$ shown in Figure 4C of the main manuscript demonstrates how vaccination gradually reduces the risk of infection over time.

### 3 Model Parameterisation and Fitting

The model is fit via a Bayesian evidence synthesis approach, defining prior distributions for our model parameters, and calculating posterior distributions given the likelihood defined in Equations (1) and (3), and data $Y_{i,t}$.

#### 3.1 Prior distributions

Prior distributions are described in Table S1. Non-informative prior distributions are assumed throughout.

**Table S1:** Inferred model parameter notations and prior distributions.

| Parameter | Description | Prior distribution | Rationale |
|---|---|---|---|
| $\phi$ | Overdispersion parameter. | $\Gamma(2,1)$ | Slightly weighted towards a large amount of overdispersion. |
| $\lambda$ | The under-reporting factor for number of first episodes. | Beta$(1,1)$ | Uninformative. |
| $\zeta_i$ | Proportion of cases in neighbouring LTLAs that cause further infections inside LTLA $i$. | Beta$(1,1)$ | Uninformative. |
| $\beta[j]$ | $j \in \{1,2,...,16\}$ Population / Variant / Funding variable coefficients. | $\mathcal{N}(0,1)$ | Uninformative. |
| $X_t$ | Random walk process. | $\mathcal{N}(0,Q)$ | Standard random walk. |
| $Q$ | Random walk standard deviation hyperparameter. | $\Gamma(1,1)$ | Uninformative. |
| $\theta_i$ | LTLA-specific error term. | $\mathcal{N}(0,1)$ | Uninformative. |

#### 3.2 Model Fitting

The model is fit using the No-U-Turn sampling variant of Hamiltonian Monte Carlo [20], via the rstan package [21] in R. 16 chains were run, with a maximum tree depth of 15, for 2000 iterations each, with 1000 iterations discarded as burn-in.

Our main model fit was produced through approximately 1,000 CPU hours via a 32-core Xeon (dual 16-core 2.6 GHz).

#### 3.3 Posterior Distributions

Table S2 below provides the mean and 95% credible interval of the posterior distributions for each of the model parameters.

**Table S2:** Mean and 95% credible intervals of posterior distributions of model parameters..

| Parameter | Posterior distribution mean (95% CrI) |
|---|---|
| $\phi$ | 19.00 (18.56 - 19.44) |
| $\lambda$ | 0.821 (0.564 - 0.992) |
| $Q$ | 0.254 (0.222 - 0.295) |
| $\beta[1]$ - Asian proportion of the population coefficient | 0.019 (-0.133 - 0.171) |
| $\beta[2]$ - Black/African/Caribbean proportion of the population coefficient | -0.169 (-0.358 - 0.021) |
| $\beta[3]$ - Other ethnicity proportion of the population coefficient | 0.054 (-0.162 - 0.272) |
| $\beta[4]$ - IMD average score coefficient | 0.115 (-0.065 - 0.296) |
| $\beta[5]$ - Proportion of the population over the age of 65 coefficient | -0.108 (-0.296 - 0.078) |
| $\beta[6]$ - Population per km$^2$ coefficient | -0.017 (-0.245 - 0.204) |
| $\beta[7]$ - Median annual income coefficient | -0.002 (-0.018 - 0.014) |
| $\beta[8]$ - Percent change in visits to workplaces coefficient | 0.029 (0.015 - 0.043) |
| $\beta[9]$ - Percent change in hours spent in residential areas coefficient | -0.301 (-0.326 - -0.276) |
| $\beta[10]$ - Percent change in visits to transit stations coefficient | 0.004 (-0.004 - 0.011) |
| $\beta[11]$ - Proportion of cases that are Alpha variant coefficient | 0.237 (0.203 - 0.271) |

Continued on next page

| **Table S2 – continued from previous page** | |
|---|---|
| Parameter | Posterior distribution mean (95% CrI) |
| $\beta[12]$ - Proportion of cases that are Delta variant coefficient | 0.439 (0.381 - 0.496) |
| $\beta[13]$ - Proportion of cases that are Omicron variant coefficient | 0.579 (0.484 - 0.675) |
| $\beta[14]$ - Unringfenced COVID-19 funding provided coefficient | -0.030 (-0.162 - 0.101) |
| $\beta[15]$ - COMF funding provided coefficient | -0.084 (-0.270 - 0.096) |
| $\beta[16]$ - ASC infection control funding coefficient | -0.100 (-0.121 - -0.078) |
| $\zeta_1$ - West Northamptonshire | 0.104 (0.034 - 0.202) |
| $\zeta_2$ - Barnsley | 0.0402 (0.0113 - 0.0818) |
| $\zeta_3$ - Birmingham | 0.111 (0.00795 - 0.275) |
| $\zeta_4$ - Bolton | 0.0194 (0.000782 - 0.0534) |
| $\zeta_5$ - Bradford | 0.0422 (0.00326 - 0.1) |
| $\zeta_6$ - Bury | 0.0357 (0.0148 - 0.0658) |
| $\zeta_7$ - Calderdale | 0.0255 (0.00805 - 0.049) |
| $\zeta_8$ - Wolverhampton | 0.312 (0.1 - 0.741) |
| $\zeta_9$ - Coventry | 0.385 (0.155 - 0.749) |
| $\zeta_{10}$ - Doncaster | 0.022 (0.00149 - 0.0609) |
| $\zeta_{11}$ - Dudley | 0.0138 (0.00199 - 0.0302) |
| $\zeta_{12}$ - Kirklees | 0.0949 (0.0311 - 0.187) |
| $\zeta_{13}$ - Knowsley | 0.306 (0.11 - 0.748) |
| $\zeta_{14}$ - Leeds | 0.0736 (0.027 - 0.133) |
| $\zeta_{15}$ - Liverpool | 0.192 (0.0732 - 0.362) |
| $\zeta_{16}$ - Manchester | 0.0443 (0.00303 - 0.112) |
| $\zeta_{17}$ - Salford | 0.0862 (0.0353 - 0.171) |
| $\zeta_{18}$ - Sandwell | 0.0912 (0.026 - 0.195) |
| $\zeta_{19}$ - Sefton | 0.131 (0.0478 - 0.251) |
| $\zeta_{20}$ - Sheffield | 0.187 (0.0278 - 0.416) |
| $\zeta_{21}$ - Solihull | 0.0891 (0.0411 - 0.169) |
| $\zeta_{22}$ - South Tyneside | 0.0804 (0.03 - 0.159) |
| $\zeta_{23}$ - Stockport | 0.135 (0.0582 - 0.258) |
| $\zeta_{24}$ - St. Helens | 0.147 (0.063 - 0.291) |
| $\zeta_{25}$ - Sunderland | 0.348 (0.158 - 0.681) |
| $\zeta_{26}$ - Tameside | 0.0498 (0.00901 - 0.108) |
| $\zeta_{27}$ - Trafford | 0.11 (0.0497 - 0.202) |
| $\zeta_{28}$ - Wakefield | 0.136 (0.0458 - 0.301) |
| $\zeta_{29}$ - Walsall | 0.0473 (0.0122 - 0.103) |
| $\zeta_{30}$ - Wigan | 0.0872 (0.0377 - 0.157) |
| $\zeta_{31}$ - Wirral | 0.0614 (0.0163 - 0.126) |
| $\zeta_{32}$ - Bath and North East Somerset | 0.0484 (0.0205 - 0.0916) |
| $\zeta_{33}$ - Bedford | 0.0332 (0.00587 - 0.0691) |
| $\zeta_{34}$ - Blackburn with Darwen | 0.0499 (0.0173 - 0.0905) |
| $\zeta_{35}$ - Blackpool | 0.717 (0.377 - 0.983) |
| $\zeta_{36}$ - Bournemouth, Christchurch and Poole | 0.17 (0.0656 - 0.315) |
| $\zeta_{37}$ - Bracknell Forest | 0.404 (0.195 - 0.785) |
| $\zeta_{38}$ - Central Bedfordshire | 0.191 (0.0577 - 0.493) |
| $\zeta_{39}$ - Cheshire East | 0.0532 (0.0276 - 0.088) |
| $\zeta_{40}$ - Cheshire West and Chester | 0.0984 (0.042 - 0.183) |
| $\zeta_{41}$ - Bristol, City of | 0.137 (0.0161 - 0.312) |
| $\zeta_{42}$ - Derby | 0.241 (0.0615 - 0.494) |
| $\zeta_{43}$ - Kingston upon Hull, City of | 0.718 (0.427 - 0.975) |
| $\zeta_{44}$ - Leicester | 0.0505 (0.00223 - 0.144) |
| $\zeta_{45}$ - Nottingham | 0.0369 (0.00111 - 0.115) |
| $\zeta_{46}$ - Peterborough | 0.159 (0.0593 - 0.302) |
| $\zeta_{47}$ - Stoke-on-Trent | 0.53 (0.285 - 0.849) |
| $\zeta_{48}$ - Oldham | 0.0761 (0.0354 - 0.132) |
| $\zeta_{49}$ - Rochdale | 0.0504 (0.00609 - 0.117) |
| $\zeta_{50}$ - Darlington | 0.242 (0.107 - 0.542) |
| $\zeta_{51}$ - Luton | 0.0627 (0.00591 - 0.144) |
| $\zeta_{52}$ - Medway | 0.257 (0.104 - 0.499) |
| $\zeta_{53}$ - Middlesbrough | 0.173 (0.0563 - 0.335) |
| $\zeta_{54}$ - Milton Keynes | 0.0311 (0.0111 - 0.0587) |
| $\zeta_{55}$ - North East Lincolnshire | 0.0802 (0.0401 - 0.132) |
| $\zeta_{56}$ - North Lincolnshire | 0.0571 (0.0211 - 0.108) |
| $\zeta_{57}$ - Halton | 0.123 (0.0618 - 0.237) |
| $\zeta_{58}$ - Shropshire | 0.0672 (0.0304 - 0.122) |
| $\zeta_{59}$ - Slough | 0.0568 (0.023 - 0.101) |
| $\zeta_{60}$ - Southend-on-Sea | 0.362 (0.0897 - 0.731) |
| $\zeta_{61}$ - South Gloucestershire | 0.677 (0.333 - 0.981) |
| $\zeta_{62}$ - Stockton-on-Tees | 0.131 (0.0541 - 0.263) |
| $\zeta_{63}$ - Swindon | 0.121 (0.0572 - 0.199) |
| $\zeta_{64}$ - Telford and Wrekin | 0.312 (0.157 - 0.545) |
| $\zeta_{65}$ - Brighton and Hove | 0.261 (0.094 - 0.475) |
| $\zeta_{66}$ - Thurrock | 0.129 (0.0587 - 0.253) |

Continued on next page

| Table S2 – continued from previous page | |
|---|---|
| Parameter | Posterior distribution mean (95% CrI) |
| $\zeta_{67}$ - Torbay | 0.262 (0.118 - 0.468) |
| $\zeta_{68}$ - Warrington | 0.0698 (0.0358 - 0.124) |
| $\zeta_{69}$ - Redcar and Cleveland | 0.167 (0.0881 - 0.297) |
| $\zeta_{70}$ - Rutland | 0.119 (0.0524 - 0.27) |
| $\zeta_{71}$ - Wiltshire | 0.199 (0.0774 - 0.406) |
| $\zeta_{72}$ - Windsor and Maidenhead | 0.0885 (0.0397 - 0.17) |
| $\zeta_{73}$ - North Somerset | 0.284 (0.148 - 0.52) |
| $\zeta_{74}$ - Plymouth | 0.11 (0.0432 - 0.198) |
| $\zeta_{75}$ - Cornwall and Isles of Scilly | 0.299 (0.142 - 0.498) |
| $\zeta_{76}$ - North Northamptonshire | 0.0211 (0.000811 - 0.0599) |
| $\zeta_{77}$ - Dorset | 0.0741 (0.0197 - 0.158) |
| $\zeta_{78}$ - York | 0.0311 (0.00383 - 0.0691) |
| $\zeta_{79}$ - East Riding of Yorkshire | 0.349 (0.121 - 0.806) |
| $\zeta_{80}$ - Southampton | 0.261 (0.0965 - 0.47) |
| $\zeta_{81}$ - North Tyneside | 0.0926 (0.0274 - 0.188) |
| $\zeta_{82}$ - Newcastle upon Tyne | 0.12 (0.0422 - 0.218) |
| $\zeta_{83}$ - Gateshead | 0.0351 (0.00747 - 0.0783) |
| $\zeta_{84}$ - County Durham | 0.542 (0.244 - 0.925) |
| $\zeta_{85}$ - Northumberland | 0.12 (0.0606 - 0.209) |
| $\zeta_{86}$ - Herefordshire, County of | 0.273 (0.146 - 0.465) |
| $\zeta_{87}$ - Portsmouth | 0.079 (0.00508 - 0.198) |
| $\zeta_{88}$ - Buckinghamshire | 0.194 (0.0613 - 0.463) |
| $\zeta_{89}$ - Wokingham | 0.38 (0.14 - 0.867) |
| $\zeta_{90}$ - West Berkshire | 0.099 (0.0475 - 0.179) |
| $\zeta_{91}$ - Reading | 0.239 (0.0976 - 0.449) |
| $\zeta_{92}$ - Hartlepool | 0.118 (0.0613 - 0.206) |
| $\zeta_{93}$ - Rotherham | 0.123 (0.0193 - 0.327) |
| $\zeta_{94}$ - Fenland | 0.145 (0.0642 - 0.292) |
| $\zeta_{95}$ - South Cambridgeshire | 0.106 (0.0468 - 0.215) |
| $\zeta_{96}$ - East Cambridgeshire | 0.102 (0.0426 - 0.207) |
| $\zeta_{97}$ - Huntingdonshire | 0.0336 (0.0174 - 0.0562) |
| $\zeta_{98}$ - Cambridge | 0.274 (0.112 - 0.488) |
| $\zeta_{99}$ - High Peak | 0.00714 (0.00284 - 0.0129) |
| $\zeta_{100}$ - Erewash | 0.117 (0.0465 - 0.235) |
| $\zeta_{101}$ - North East Derbyshire | 0.0224 (0.00903 - 0.0407) |
| $\zeta_{102}$ - Amber Valley | 0.0863 (0.0438 - 0.158) |
| $\zeta_{103}$ - Chesterfield | 0.535 (0.211 - 0.944) |
| $\zeta_{104}$ - Brentwood | 0.104 (0.0442 - 0.224) |
| $\zeta_{105}$ - Epping Forest | 0.0587 (0.0177 - 0.143) |
| $\zeta_{106}$ - Uttlesford | 0.34 (0.122 - 0.817) |
| $\zeta_{107}$ - Chelmsford | 0.211 (0.0817 - 0.443) |
| $\zeta_{108}$ - Harlow | 0.278 (0.14 - 0.5) |
| $\zeta_{109}$ - Cotswold | 0.0494 (0.0207 - 0.109) |
| $\zeta_{110}$ - Tewkesbury | 0.396 (0.182 - 0.828) |
| $\zeta_{111}$ - Gloucester | 0.526 (0.288 - 0.85) |
| $\zeta_{112}$ - Cheltenham | 0.772 (0.444 - 0.99) |
| $\zeta_{113}$ - New Forest | 0.104 (0.0394 - 0.232) |
| $\zeta_{114}$ - Hart | 0.186 (0.0785 - 0.402) |
| $\zeta_{115}$ - Gosport | 0.101 (0.0505 - 0.173) |
| $\zeta_{116}$ - Rushmoor | 0.132 (0.0616 - 0.233) |
| $\zeta_{117}$ - Three Rivers | 0.262 (0.0646 - 0.786) |
| $\zeta_{118}$ - Broxbourne | 0.611 (0.253 - 0.975) |
| $\zeta_{119}$ - Dacorum | 0.256 (0.104 - 0.6) |
| $\zeta_{120}$ - East Hertfordshire | 0.185 (0.1 - 0.324) |
| $\zeta_{121}$ - St Albans | 0.0756 (0.0339 - 0.136) |
| $\zeta_{122}$ - Welwyn Hatfield | 0.17 (0.0798 - 0.321) |
| $\zeta_{123}$ - North Hertfordshire | 0.0476 (0.0211 - 0.0889) |
| $\zeta_{124}$ - Watford | 0.175 (0.0884 - 0.299) |
| $\zeta_{125}$ - Stevenage | 0.15 (0.0789 - 0.244) |
| $\zeta_{126}$ - Tunbridge Wells | 0.214 (0.114 - 0.38) |
| $\zeta_{127}$ - Sevenoaks | 0.081 (0.0364 - 0.157) |
| $\zeta_{128}$ - Tonbridge and Malling | 0.17 (0.073 - 0.349) |
| $\zeta_{129}$ - Ashford | 0.13 (0.0669 - 0.226) |
| $\zeta_{130}$ - Maidstone | 0.366 (0.167 - 0.738) |
| $\zeta_{131}$ - Dartford | 0.313 (0.112 - 0.72) |
| $\zeta_{132}$ - Gravesham | 0.143 (0.0546 - 0.301) |
| $\zeta_{133}$ - Chorley | 0.0141 (0.00459 - 0.0264) |
| $\zeta_{134}$ - Rossendale | 0.0361 (0.0192 - 0.0612) |
| $\zeta_{135}$ - Preston | 0.0598 (0.00534 - 0.139) |
| $\zeta_{136}$ - Pendle | 0.0587 (0.0329 - 0.0941) |
| $\zeta_{137}$ - Burnley | 0.0759 (0.0259 - 0.147) |
| $\zeta_{138}$ - Hinckley and Bosworth | 0.168 (0.0775 - 0.326) |

Continued on next page

| Table S2 – continued from previous page | |
|---|---|
| Parameter | Posterior distribution mean (95% CrI) |
| $\zeta_{139}$ - Melton | 0.114 (0.0632 - 0.2) |
| $\zeta_{140}$ - Harborough | 0.0186 (0.00887 - 0.0322) |
| $\zeta_{141}$ - Blaby | 0.0323 (0.0144 - 0.0574) |
| $\zeta_{142}$ - Charnwood | 0.0291 (0.00992 - 0.0545) |
| $\zeta_{143}$ - Oadby and Wigston | 0.0768 (0.0384 - 0.138) |
| $\zeta_{144}$ - West Lindsey | 0.113 (0.0611 - 0.198) |
| $\zeta_{145}$ - South Kesteven | 0.0628 (0.0296 - 0.112) |
| $\zeta_{146}$ - South Holland | 0.0608 (0.0305 - 0.104) |
| $\zeta_{147}$ - Boston | 0.213 (0.116 - 0.358) |
| $\zeta_{148}$ - North Kesteven | 0.0635 (0.0319 - 0.111) |
| $\zeta_{149}$ - Lincoln | 0.11 (0.0187 - 0.233) |
| $\zeta_{150}$ - Breckland | 0.188 (0.0822 - 0.362) |
| $\zeta_{151}$ - Norwich | 0.483 (0.235 - 0.853) |
| $\zeta_{152}$ - Craven | 0.015 (0.009 - 0.0231) |
| $\zeta_{153}$ - Bassetlaw | 0.123 (0.0621 - 0.232) |
| $\zeta_{154}$ - Gedling | 0.175 (0.0783 - 0.364) |
| $\zeta_{155}$ - Ashfield | 0.0931 (0.0484 - 0.167) |
| $\zeta_{156}$ - Newark and Sherwood | 0.0682 (0.0253 - 0.133) |
| $\zeta_{157}$ - Broxtowe | 0.167 (0.0637 - 0.396) |
| $\zeta_{158}$ - Mansfield | 0.184 (0.0915 - 0.329) |
| $\zeta_{159}$ - West Oxfordshire | 0.528 (0.215 - 0.942) |
| $\zeta_{160}$ - Oxford | 0.0797 (0.0174 - 0.16) |
| $\zeta_{161}$ - Mendip | 0.0682 (0.0361 - 0.119) |
| $\zeta_{162}$ - Cannock Chase | 0.144 (0.0726 - 0.265) |
| $\zeta_{163}$ - Mid Suffolk | 0.367 (0.132 - 0.859) |
| $\zeta_{164}$ - Ipswich | 0.214 (0.0897 - 0.387) |
| $\zeta_{165}$ - Waverley | 0.218 (0.104 - 0.434) |
| $\zeta_{166}$ - Woking | 0.0466 (0.00566 - 0.106) |
| $\zeta_{167}$ - Surrey Heath | 0.194 (0.0829 - 0.462) |
| $\zeta_{168}$ - Runnymede | 0.156 (0.0723 - 0.306) |
| $\zeta_{169}$ - Guildford | 0.499 (0.177 - 0.948) |
| $\zeta_{170}$ - Reigate and Banstead | 0.187 (0.0848 - 0.365) |
| $\zeta_{171}$ - Mole Valley | 0.497 (0.167 - 0.953) |
| $\zeta_{172}$ - Elmbridge | 0.354 (0.159 - 0.764) |
| $\zeta_{173}$ - Epsom and Ewell | 0.404 (0.16 - 0.872) |
| $\zeta_{174}$ - Rugby | 0.0477 (0.0191 - 0.0949) |
| $\zeta_{175}$ - Nuneaton and Bedworth | 0.145 (0.0568 - 0.281) |
| $\zeta_{176}$ - Mid Sussex | 0.121 (0.0566 - 0.225) |
| $\zeta_{177}$ - Crawley | 0.164 (0.0887 - 0.273) |
| $\zeta_{178}$ - Malvern Hills | 0.0609 (0.0268 - 0.119) |
| $\zeta_{179}$ - Wyre Forest | 0.13 (0.0649 - 0.236) |
| $\zeta_{180}$ - Wychavon | 0.198 (0.0778 - 0.444) |
| $\zeta_{181}$ - Bromsgrove | 0.00983 (0.00341 - 0.0183) |
| $\zeta_{182}$ - Worcester | 0.355 (0.153 - 0.653) |
| $\zeta_{183}$ - Redditch | 0.223 (0.113 - 0.386) |
| $\zeta_{184}$ - Broadland | 0.296 (0.128 - 0.616) |
| $\zeta_{185}$ - South Lakeland | 0.298 (0.141 - 0.617) |
| $\zeta_{186}$ - Copeland | 0.176 (0.0933 - 0.3) |
| $\zeta_{187}$ - Barrow-in-Furness | 0.214 (0.128 - 0.331) |
| $\zeta_{188}$ - South Derbyshire | 0.165 (0.0649 - 0.388) |
| $\zeta_{189}$ - Bolsover | 0.0678 (0.0307 - 0.135) |
| $\zeta_{190}$ - Derbyshire Dales | 0.0132 (0.00794 - 0.0203) |
| $\zeta_{191}$ - Teignbridge | 0.13 (0.0446 - 0.286) |
| $\zeta_{192}$ - West Devon | 0.0309 (0.0164 - 0.053) |
| $\zeta_{193}$ - Wealden | 0.619 (0.258 - 0.973) |
| $\zeta_{194}$ - Eastbourne | 0.456 (0.236 - 0.762) |
| $\zeta_{195}$ - Hastings | 0.755 (0.437 - 0.985) |
| $\zeta_{196}$ - Rochford | 0.153 (0.0498 - 0.409) |
| $\zeta_{197}$ - Tendring | 0.273 (0.132 - 0.487) |
| $\zeta_{198}$ - Colchester | 0.312 (0.16 - 0.53) |
| $\zeta_{199}$ - Maldon | 0.133 (0.0574 - 0.284) |
| $\zeta_{200}$ - Braintree | 0.0924 (0.0517 - 0.148) |
| $\zeta_{201}$ - Basildon | 0.265 (0.124 - 0.494) |
| $\zeta_{202}$ - Castle Point | 0.204 (0.0977 - 0.424) |
| $\zeta_{203}$ - Exeter | 0.0301 (0.00132 - 0.0861) |
| $\zeta_{204}$ - Lewes | 0.221 (0.0922 - 0.506) |
| $\zeta_{205}$ - Rother | 0.0946 (0.0383 - 0.196) |
| $\zeta_{206}$ - Hertsmere | 0.222 (0.0659 - 0.665) |
| $\zeta_{207}$ - Folkestone and Hythe | 0.246 (0.127 - 0.438) |
| $\zeta_{208}$ - Thanet | 0.201 (0.111 - 0.316) |
| $\zeta_{209}$ - Canterbury | 0.0695 (0.027 - 0.124) |
| $\zeta_{210}$ - Dover | 0.263 (0.138 - 0.463) |

Continued on next page

**Table S2 – continued from previous page**

| Parameter | Posterior distribution mean (95% CrI) |
|---|---|
| $\zeta_{211}$ - Swale | 0.0944 (0.0525 - 0.152) |
| $\zeta_{212}$ - West Lancashire | 0.0712 (0.0298 - 0.143) |
| $\zeta_{213}$ - Lancaster | 0.143 (0.0706 - 0.239) |
| $\zeta_{214}$ - South Ribble | 0.0359 (0.0156 - 0.0644) |
| $\zeta_{215}$ - Fylde | 0.235 (0.0989 - 0.545) |
| $\zeta_{216}$ - Wyre | 0.0609 (0.0282 - 0.107) |
| $\zeta_{217}$ - Ribble Valley | 0.0251 (0.0142 - 0.0398) |
| $\zeta_{218}$ - Hyndburn | 0.355 (0.189 - 0.657) |
| $\zeta_{219}$ - North West Leicestershire | 0.0966 (0.0451 - 0.181) |
| $\zeta_{220}$ - East Lindsey | 0.207 (0.112 - 0.353) |
| $\zeta_{221}$ - King's Lynn and West Norfolk | 0.277 (0.135 - 0.544) |
| $\zeta_{222}$ - Selby | 0.0293 (0.015 - 0.0511) |
| $\zeta_{223}$ - Richmondshire | 0.0269 (0.011 - 0.0526) |
| $\zeta_{224}$ - Harrogate | 0.00962 (0.00412 - 0.0163) |
| $\zeta_{225}$ - West Suffolk | 0.229 (0.103 - 0.459) |
| $\zeta_{226}$ - Tandridge | 0.143 (0.0431 - 0.427) |
| $\zeta_{227}$ - Spelthorne | 0.0573 (0.0252 - 0.114) |
| $\zeta_{228}$ - North Warwickshire | 0.014 (0.0075 - 0.0234) |
| $\zeta_{229}$ - Warwick | 0.0677 (0.0343 - 0.113) |
| $\zeta_{230}$ - Stratford-on-Avon | 0.052 (0.0234 - 0.101) |
| $\zeta_{231}$ - Arun | 0.219 (0.0931 - 0.403) |
| $\zeta_{232}$ - Adur | 0.134 (0.0555 - 0.308) |
| $\zeta_{233}$ - Chichester | 0.453 (0.197 - 0.892) |
| $\zeta_{234}$ - Horsham | 0.08 (0.0308 - 0.161) |
| $\zeta_{235}$ - Worthing | 0.672 (0.38 - 0.969) |
| $\zeta_{236}$ - Kingston upon Thames | 0.0483 (0.00826 - 0.107) |
| $\zeta_{237}$ - Croydon | 0.381 (0.137 - 0.821) |
| $\zeta_{238}$ - Bromley | 0.147 (0.0518 - 0.341) |
| $\zeta_{239}$ - Hounslow | 0.149 (0.0374 - 0.336) |
| $\zeta_{240}$ - Ealing | 0.503 (0.133 - 0.953) |
| $\zeta_{241}$ - Havering | 0.257 (0.105 - 0.556) |
| $\zeta_{242}$ - Hillingdon | 0.0995 (0.0285 - 0.223) |
| $\zeta_{243}$ - Harrow | 0.201 (0.0471 - 0.583) |
| $\zeta_{244}$ - Brent | 0.0854 (0.0157 - 0.202) |
| $\zeta_{245}$ - Barnet | 0.141 (0.0383 - 0.315) |
| $\zeta_{246}$ - Lambeth | 0.227 (0.0858 - 0.518) |
| $\zeta_{247}$ - Southwark | 0.312 (0.103 - 0.787) |
| $\zeta_{248}$ - Lewisham | 0.578 (0.195 - 0.971) |
| $\zeta_{249}$ - Greenwich | 0.187 (0.0639 - 0.452) |
| $\zeta_{250}$ - Bexley | 0.567 (0.208 - 0.967) |
| $\zeta_{251}$ - Enfield | 0.323 (0.124 - 0.739) |
| $\zeta_{252}$ - Waltham Forest | 0.161 (0.0302 - 0.478) |
| $\zeta_{253}$ - Redbridge | 0.614 (0.254 - 0.972) |
| $\zeta_{254}$ - Sutton | 0.568 (0.229 - 0.965) |
| $\zeta_{255}$ - Richmond upon Thames | 0.156 (0.0665 - 0.321) |
| $\zeta_{256}$ - Merton | 0.0571 (0.0142 - 0.126) |
| $\zeta_{257}$ - Wandsworth | 0.342 (0.113 - 0.815) |
| $\zeta_{258}$ - Hammersmith and Fulham | 0.207 (0.0842 - 0.492) |
| $\zeta_{259}$ - Kensington and Chelsea | 0.394 (0.113 - 0.912) |
| $\zeta_{260}$ - Westminster | 0.0494 (0.00577 - 0.125) |
| $\zeta_{261}$ - Camden | 0.321 (0.12 - 0.786) |
| $\zeta_{262}$ - Tower Hamlets | 0.228 (0.0905 - 0.489) |
| $\zeta_{263}$ - Islington | 0.221 (0.0761 - 0.492) |
| $\zeta_{264}$ - Hackney and City of London | 0.112 (0.0511 - 0.21) |
| $\zeta_{265}$ - Haringey | 0.603 (0.217 - 0.974) |
| $\zeta_{266}$ - Newham | 0.129 (0.0268 - 0.312) |
| $\zeta_{267}$ - Barking and Dagenham | 0.512 (0.184 - 0.95) |
| $\zeta_{268}$ - North Norfolk | 0.0985 (0.0522 - 0.173) |
| $\zeta_{269}$ - Rushcliffe | 0.057 (0.0292 - 0.0986) |
| $\zeta_{270}$ - Sedgemoor | 0.187 (0.0978 - 0.32) |
| $\zeta_{271}$ - Staffordshire Moorlands | 0.0674 (0.031 - 0.132) |
| $\zeta_{272}$ - South Staffordshire | 0.064 (0.0308 - 0.126) |
| $\zeta_{273}$ - Lichfield | 0.00987 (0.00451 - 0.0173) |
| $\zeta_{274}$ - Newcastle-under-Lyme | 0.202 (0.0926 - 0.426) |
| $\zeta_{275}$ - Stafford | 0.087 (0.041 - 0.168) |
| $\zeta_{276}$ - East Staffordshire | 0.159 (0.0785 - 0.275) |
| $\zeta_{277}$ - Tamworth | 0.779 (0.482 - 0.99) |
| $\zeta_{278}$ - Allerdale | 0.175 (0.0994 - 0.281) |
| $\zeta_{279}$ - South Hams | 0.055 (0.0275 - 0.0989) |
| $\zeta_{280}$ - Torridge | 0.0349 (0.0209 - 0.054) |
| $\zeta_{281}$ - North Devon | 0.0808 (0.0464 - 0.127) |
| $\zeta_{282}$ - South Somerset | 0.0674 (0.035 - 0.114) |

Continued on next page

| Table S2 – continued from previous page | |
| --- | --- |
| Parameter | Posterior distribution mean (95% CrI) |
| $\zeta_{283}$ - Mid Devon | 0.171 (0.0766 - 0.332) |
| $\zeta_{284}$ - East Devon | 0.124 (0.0639 - 0.222) |
| $\zeta_{285}$ - Ryedale | 0.0264 (0.0133 - 0.046) |
| $\zeta_{286}$ - Hambleton | 0.0852 (0.0446 - 0.157) |
| $\zeta_{287}$ - Scarborough | 0.148 (0.0775 - 0.257) |
| $\zeta_{288}$ - Eastleigh | 0.265 (0.106 - 0.614) |
| $\zeta_{289}$ - Stroud | 0.192 (0.0849 - 0.398) |
| $\zeta_{290}$ - Forest of Dean | 0.0457 (0.0201 - 0.0856) |
| $\zeta_{291}$ - Winchester | 0.262 (0.117 - 0.584) |
| $\zeta_{292}$ - Test Valley | 0.359 (0.115 - 0.867) |
| $\zeta_{293}$ - Havant | 0.132 (0.0637 - 0.241) |
| $\zeta_{294}$ - East Hampshire | 0.34 (0.121 - 0.811) |
| $\zeta_{295}$ - Basingstoke and Deane | 0.111 (0.0509 - 0.21) |
| $\zeta_{296}$ - Fareham | 0.429 (0.175 - 0.882) |
| $\zeta_{297}$ - Vale of White Horse | 0.377 (0.119 - 0.883) |
| $\zeta_{298}$ - Cherwell | 0.107 (0.0525 - 0.206) |
| $\zeta_{299}$ - Carlisle | 0.0601 (0.0312 - 0.0972) |
| $\zeta_{300}$ - Eden | 0.0148 (0.00973 - 0.0216) |
| $\zeta_{301}$ - Great Yarmouth | 0.123 (0.0645 - 0.201) |
| $\zeta_{302}$ - East Suffolk | 0.329 (0.142 - 0.654) |
| $\zeta_{303}$ - South Norfolk | 0.077 (0.0289 - 0.155) |
| $\zeta_{304}$ - Babergh | 0.152 (0.0673 - 0.347) |
| $\zeta_{305}$ - Somerset West and Taunton | 0.189 (0.113 - 0.3) |
| $\zeta_{306}$ - South Oxfordshire | 0.109 (0.0586 - 0.194) |
| $X_1$ | 0.125 (-0.122 - 0.485) |
| $X_2$ | -0.211 (-0.266 - -0.158) |
| $X_3$ | 0.0456 (-0.0125 - 0.103) |
| $X_4$ | -0.0714 (-0.134 - -0.00966) |
| $X_5$ | -0.00626 (-0.0729 - 0.0611) |
| $X_6$ | -0.18 (-0.252 - -0.108) |
| $X_7$ | 0.00684 (-0.0695 - 0.0844) |
| $X_8$ | 0.297 (0.22 - 0.373) |
| $X_9$ | -0.108 (-0.182 - -0.0343) |
| $X_{10}$ | -0.0188 (-0.0873 - 0.0516) |
| $X_{11}$ | -0.0763 (-0.144 - -0.0097) |
| $X_{12}$ | -0.0453 (-0.11 - 0.018) |
| $X_{13}$ | 0.0408 (-0.0213 - 0.104) |
| $X_{14}$ | -0.139 (-0.198 - -0.0792) |
| $X_{15}$ | 0.0788 (0.0198 - 0.137) |
| $X_{16}$ | 0.468 (0.414 - 0.521) |
| $X_{17}$ | -0.523 (-0.571 - -0.475) |
| $X_{18}$ | -0.272 (-0.319 - -0.225) |
| $X_{19}$ | 0.496 (0.45 - 0.542) |
| $X_{20}$ | 0.183 (0.14 - 0.226) |
| $X_{21}$ | -0.203 (-0.244 - -0.163) |
| $X_{22}$ | -0.273 (-0.313 - -0.233) |
| $X_{23}$ | 0.22 (0.18 - 0.259) |
| $X_{24}$ | -0.199 (-0.238 - -0.161) |
| $X_{25}$ | 0.122 (0.0828 - 0.161) |
| $X_{26}$ | 0.112 (0.0736 - 0.15) |
| $X_{27}$ | -0.336 (-0.374 - -0.297) |
| $X_{28}$ | 0.00712 (-0.0317 - 0.0455) |
| $X_{29}$ | 0.225 (0.185 - 0.265) |
| $X_{30}$ | 0.216 (0.177 - 0.255) |
| $X_{31}$ | 0.0933 (0.0557 - 0.132) |
| $X_{32}$ | -0.1 (-0.139 - -0.0607) |
| $X_{33}$ | 0.538 (0.496 - 0.581) |
| $X_{34}$ | -0.436 (-0.474 - -0.398) |
| $X_{35}$ | -0.305 (-0.343 - -0.268) |
| $X_{36}$ | -0.0298 (-0.0668 - 0.0074) |
| $X_{37}$ | -0.119 (-0.156 - -0.0817) |
| $X_{38}$ | 0.0248 (-0.0129 - 0.0629) |
| $X_{39}$ | -0.0812 (-0.12 - -0.0428) |
| $X_{40}$ | 0.208 (0.169 - 0.247) |
| $X_{41}$ | -0.205 (-0.245 - -0.165) |
| $X_{42}$ | -0.174 (-0.215 - -0.132) |
| $X_{43}$ | 0.247 (0.204 - 0.29) |
| $X_{44}$ | -0.0913 (-0.135 - -0.0473) |
| $X_{45}$ | -0.024 (-0.0696 - 0.0209) |
| $X_{46}$ | -0.259 (-0.306 - -0.211) |
| $X_{47}$ | 0.0658 (0.0144 - 0.118) |

Continued on next page

| Table S2 – continued from previous page | |
|---|---|
| Parameter | Posterior distribution mean (95% CrI) |
| $X_{48}$ | -0.111 (-0.163 - -0.0573) |
| $X_{49}$ | 0.0537 (-0.0195 - 0.128) |
| $X_{50}$ | -0.0801 (-0.137 - -0.022) |
| $X_{51}$ | 0.132 (0.0752 - 0.189) |
| $X_{52}$ | -0.0494 (-0.105 - 0.00678) |
| $X_{53}$ | -0.00115 (-0.0579 - 0.0542) |
| $X_{54}$ | 0.243 (0.189 - 0.297) |
| $X_{55}$ | 0.0839 (0.0323 - 0.135) |
| $X_{56}$ | -0.0993 (-0.146 - -0.0528) |
| $X_{57}$ | -0.192 (-0.236 - -0.148) |
| $X_{58}$ | 0.195 (0.153 - 0.236) |
| $X_{59}$ | 0.0444 (0.00568 - 0.0833) |
| $X_{60}$ | -0.21 (-0.248 - -0.172) |
| $X_{61}$ | 0.151 (0.114 - 0.189) |
| $X_{62}$ | -0.834 (-0.872 - -0.796) |
| $X_{63}$ | 0.0852 (0.048 - 0.124) |
| $X_{64}$ | 0.362 (0.324 - 0.399) |
| $X_{65}$ | -0.0213 (-0.058 - 0.0158) |
| $X_{66}$ | 0.0371 (-0.000431 - 0.0746) |
| $X_{67}$ | -0.181 (-0.218 - -0.144) |
| $X_{68}$ | 0.149 (0.111 - 0.186) |
| $X_{69}$ | -0.218 (-0.254 - -0.181) |
| $X_{70}$ | -0.0497 (-0.0871 - -0.0125) |
| $X_{71}$ | 0.338 (0.3 - 0.376) |
| $X_{72}$ | -0.224 (-0.261 - -0.187) |
| $X_{73}$ | 0.153 (0.116 - 0.19) |
| $X_{74}$ | 0.122 (0.0854 - 0.159) |
| $X_{75}$ | -0.192 (-0.229 - -0.154) |
| $X_{76}$ | -0.141 (-0.178 - -0.104) |
| $X_{77}$ | -0.00607 (-0.0426 - 0.031) |
| $X_{78}$ | 0.266 (0.229 - 0.303) |
| $X_{79}$ | -0.0126 (-0.05 - 0.0248) |
| $X_{80}$ | -0.0904 (-0.128 - -0.053) |
| $X_{81}$ | 0.135 (0.0984 - 0.172) |
| $X_{82}$ | -0.0582 (-0.0952 - -0.0211) |
| $X_{83}$ | 0.403 (0.365 - 0.44) |
| $X_{84}$ | -0.0041 (-0.0509 - 0.0428) |
| $X_{85}$ | 0.119 (0.0713 - 0.165) |
| $X_{86}$ | -0.176 (-0.215 - -0.138) |
| $X_{87}$ | -1.04 (-1.08 - -0.996) |
| $X_{88}$ | 0.571 (0.531 - 0.61) |
| $X_{89}$ | 0.0944 (0.0565 - 0.132) |
| $X_{90}$ | -0.228 (-0.266 - -0.191) |
| $X_{91}$ | -0.172 (-0.21 - -0.135) |
| $X_{92}$ | -0.000335 (-0.0382 - 0.0378) |
| $X_{93}$ | 0.101 (0.0621 - 0.139) |
| $X_{94}$ | 0.444 (0.405 - 0.483) |
| $X_{95}$ | 0.276 (0.237 - 0.314) |
| $\theta_1$ - West Northamptonshire | 0.225 (-0.0965 - 0.527) |
| $\theta_2$ - Barnsley | 0.125 (-0.204 - 0.431) |
| $\theta_3$ - Birmingham | 0.24 (-0.232 - 0.701) |
| $\theta_4$ - Bolton | 0.339 (0.0667 - 0.604) |
| $\theta_5$ - Bradford | 0.0727 (-0.334 - 0.488) |
| $\theta_6$ - Bury | 0.327 (0.077 - 0.564) |
| $\theta_7$ - Calderdale | 0.148 (-0.108 - 0.391) |
| $\theta_8$ - Wolverhampton | -0.265 (-0.9 - 0.259) |
| $\theta_9$ - Coventry | -0.217 (-0.62 - 0.147) |
| $\theta_{10}$ - Doncaster | 0.222 (-0.0856 - 0.5) |
| $\theta_{11}$ - Dudley | 0.376 (0.167 - 0.588) |
| $\theta_{12}$ - Kirklees | -0.0513 (-0.451 - 0.317) |
| $\theta_{13}$ - Knowsley | -0.69 (-1.51 - 0.00959) |
| $\theta_{14}$ - Leeds | 0.346 (0.0493 - 0.634) |
| $\theta_{15}$ - Liverpool | -0.0365 (-0.524 - 0.426) |
| $\theta_{16}$ - Manchester | 0.196 (-0.249 - 0.629) |
| $\theta_{17}$ - Salford | -0.00668 (-0.456 - 0.4) |
| $\theta_{18}$ - Sandwell | -0.0221 (-0.483 - 0.4) |
| $\theta_{19}$ - Sefton | 0.498 (0.0711 - 0.908) |
| $\theta_{20}$ - Sheffield | 0.218 (-0.185 - 0.605) |
| $\theta_{21}$ - Solihull | 0.187 (-0.205 - 0.522) |
| $\theta_{22}$ - South Tyneside | 0.149 (-0.264 - 0.521) |
| $\theta_{23}$ - Stockport | 0.254 (-0.112 - 0.583) |

Continued on next page

| Table S2 – continued from previous page<br>Parameter | Posterior distribution mean (95% CrI) |
|---|---|
| $\theta_{24}$ - St. Helens | -0.083 (-0.583 - 0.357) |
| $\theta_{25}$ - Sunderland | -0.193 (-0.695 - 0.234) |
| $\theta_{26}$ - Tameside | 0.211 (-0.106 - 0.507) |
| $\theta_{27}$ - Trafford | 0.269 (-0.0934 - 0.598) |
| $\theta_{28}$ - Wakefield | -0.205 (-0.68 - 0.171) |
| $\theta_{29}$ - Walsall | -0.014 (-0.387 - 0.329) |
| $\theta_{30}$ - Wigan | 0.169 (-0.161 - 0.497) |
| $\theta_{31}$ - Wirral | 0.502 (0.111 - 0.885) |
| $\theta_{32}$ - Bath and North East Somerset | 0.339 (0.0247 - 0.631) |
| $\theta_{33}$ - Bedford | 0.462 (0.106 - 0.811) |
| $\theta_{34}$ - Blackburn with Darwen | -0.00456 (-0.54 - 0.545) |
| $\theta_{35}$ - Blackpool | -0.132 (-0.691 - 0.421) |
| $\theta_{36}$ - Bournemouth, Christchurch and Poole | 0.546 (0.32 - 0.778) |
| $\theta_{37}$ - Bracknell Forest | -0.372 (-0.916 - 0.0978) |
| $\theta_{38}$ - Central Bedfordshire | -0.312 (-1.04 - 0.219) |
| $\theta_{39}$ - Cheshire East | 0.409 (0.18 - 0.629) |
| $\theta_{40}$ - Cheshire West and Chester | 0.164 (-0.14 - 0.427) |
| $\theta_{41}$ - Bristol, City of | 0.475 (0.159 - 0.784) |
| $\theta_{42}$ - Derby | 0.194 (-0.0893 - 0.461) |
| $\theta_{43}$ - Kingston upon Hull, City of | -0.4 (-0.82 - 0.0253) |
| $\theta_{44}$ - Leicester | 0.325 (-0.183 - 0.835) |
| $\theta_{45}$ - Nottingham | 0.394 (0.0735 - 0.709) |
| $\theta_{46}$ - Peterborough | -0.142 (-0.541 - 0.233) |
| $\theta_{47}$ - Stoke-on-Trent | -0.145 (-0.468 - 0.173) |
| $\theta_{48}$ - Oldham | -0.118 (-0.535 - 0.297) |
| $\theta_{49}$ - Rochdale | 0.0504 (-0.361 - 0.436) |
| $\theta_{50}$ - Darlington | -0.53 (-1.2 - -0.0524) |
| $\theta_{51}$ - Luton | 0.336 (-0.0968 - 0.767) |
| $\theta_{52}$ - Medway | -0.172 (-0.585 - 0.189) |
| $\theta_{53}$ - Middlesbrough | 0.0616 (-0.402 - 0.496) |
| $\theta_{54}$ - Milton Keynes | 0.385 (0.0468 - 0.721) |
| $\theta_{55}$ - North East Lincolnshire | 0.19 (-0.083 - 0.458) |
| $\theta_{56}$ - North Lincolnshire | 0.132 (-0.139 - 0.381) |
| $\theta_{57}$ - Halton | -0.425 (-0.967 - 0.0462) |
| $\theta_{58}$ - Shropshire | 0.261 (-0.0194 - 0.521) |
| $\theta_{59}$ - Slough | 0.101 (-0.55 - 0.764) |
| $\theta_{60}$ - Southend-on-Sea | 0.433 (0.0978 - 0.768) |
| $\theta_{61}$ - South Gloucestershire | -0.786 (-1.2 - -0.282) |
| $\theta_{62}$ - Stockton-on-Tees | -0.0825 (-0.48 - 0.261) |
| $\theta_{63}$ - Swindon | 0.117 (-0.139 - 0.367) |
| $\theta_{64}$ - Telford and Wrekin | -0.302 (-0.664 - 0.023) |
| $\theta_{65}$ - Brighton and Hove | 0.193 (-0.149 - 0.529) |
| $\theta_{66}$ - Thurrock | -0.156 (-0.701 - 0.347) |
| $\theta_{67}$ - Torbay | 0.401 (0.0351 - 0.777) |
| $\theta_{68}$ - Warrington | 0.163 (-0.183 - 0.482) |
| $\theta_{69}$ - Redcar and Cleveland | -0.129 (-0.513 - 0.207) |
| $\theta_{70}$ - Rutland | -0.384 (-1.09 - 0.143) |
| $\theta_{71}$ - Wiltshire | -0.0243 (-0.471 - 0.342) |
| $\theta_{72}$ - Windsor and Maidenhead | 0.324 (-0.097 - 0.69) |
| $\theta_{73}$ - North Somerset | -0.0023 (-0.438 - 0.358) |
| $\theta_{74}$ - Plymouth | 0.128 (-0.158 - 0.417) |
| $\theta_{75}$ - Cornwall and Isles of Scilly | 0.142 (-0.153 - 0.433) |
| $\theta_{76}$ - North Northamptonshire | 0.285 (0.0511 - 0.498) |
| $\theta_{77}$ - Dorset | 0.372 (-0.0247 - 0.739) |
| $\theta_{78}$ - York | 0.436 (0.192 - 0.683) |
| $\theta_{79}$ - East Riding of Yorkshire | -0.1 (-0.729 - 0.398) |
| $\theta_{80}$ - Southampton | 0.0384 (-0.295 - 0.37) |
| $\theta_{81}$ - North Tyneside | 0.376 (0.0495 - 0.691) |
| $\theta_{82}$ - Newcastle upon Tyne | 0.271 (-0.055 - 0.59) |
| $\theta_{83}$ - Gateshead | 0.374 (0.0093 - 0.709) |
| $\theta_{84}$ - County Durham | -0.215 (-0.671 - 0.243) |
| $\theta_{85}$ - Northumberland | 0.186 (-0.15 - 0.5) |
| $\theta_{86}$ - Herefordshire, County of | -0.0538 (-0.388 - 0.248) |
| $\theta_{87}$ - Portsmouth | 0.22 (-0.157 - 0.595) |
| $\theta_{88}$ - Buckinghamshire | 0.0657 (-0.504 - 0.505) |
| $\theta_{89}$ - Wokingham | -0.55 (-1.34 - 0.13) |
| $\theta_{90}$ - West Berkshire | 0.0346 (-0.366 - 0.396) |
| $\theta_{91}$ - Reading | 0.244 (-0.105 - 0.568) |
| $\theta_{92}$ - Hartlepool | -0.225 (-0.678 - 0.191) |
| $\theta_{93}$ - Rotherham | -0.0383 (-0.585 - 0.39) |
| $\theta_{94}$ - Fenland | -0.494 (-1.01 - -0.0623) |
| $\theta_{95}$ - South Cambridgeshire | 0.0718 (-0.412 - 0.464) |

Continued on next page

| Table S2 – continued from previous page<br>Parameter | Posterior distribution mean (95% CrI) |
|---|---|
| $\theta_{96}$ - East Cambridgeshire | -0.041 (-0.516 - 0.349) |
| $\theta_{97}$ - Huntingdonshire | 0.238 (-0.00441 - 0.466) |
| $\theta_{98}$ - Cambridge | 0.386 (0.0417 - 0.724) |
| $\theta_{99}$ - High Peak | 0.412 (0.194 - 0.623) |
| $\theta_{100}$ - Erewash | 0.0199 (-0.414 - 0.383) |
| $\theta_{101}$ - North East Derbyshire | 0.422 (0.139 - 0.689) |
| $\theta_{102}$ - Amber Valley | 0.0586 (-0.292 - 0.354) |
| $\theta_{103}$ - Chesterfield | -0.154 (-0.559 - 0.254) |
| $\theta_{104}$ - Brentwood | 0.0809 (-0.483 - 0.527) |
| $\theta_{105}$ - Epping Forest | 0.0784 (-0.497 - 0.522) |
| $\theta_{106}$ - Uttlesford | -0.696 (-1.52 - -0.0276) |
| $\theta_{107}$ - Chelmsford | 0.149 (-0.344 - 0.542) |
| $\theta_{108}$ - Harlow | -0.11 (-0.501 - 0.243) |
| $\theta_{109}$ - Cotswold | -0.0878 (-0.685 - 0.358) |
| $\theta_{110}$ - Tewkesbury | -0.768 (-1.43 - -0.24) |
| $\theta_{111}$ - Gloucester | 0.00308 (-0.352 - 0.345) |
| $\theta_{112}$ - Cheltenham | -0.0557 (-0.331 - 0.265) |
| $\theta_{113}$ - New Forest | -0.0207 (-0.653 - 0.49) |
| $\theta_{114}$ - Hart | -0.122 (-0.754 - 0.409) |
| $\theta_{115}$ - Gosport | 0.102 (-0.271 - 0.478) |
| $\theta_{116}$ - Rushmoor | 0.128 (-0.232 - 0.469) |
| $\theta_{117}$ - Three Rivers | -0.774 (-1.88 - 0.0723) |
| $\theta_{118}$ - Broxbourne | -0.926 (-1.45 - -0.289) |
| $\theta_{119}$ - Dacorum | -0.38 (-1.08 - 0.118) |
| $\theta_{120}$ - East Hertfordshire | 0.126 (-0.288 - 0.495) |
| $\theta_{121}$ - St Albans | 0.457 (0.0961 - 0.795) |
| $\theta_{122}$ - Welwyn Hatfield | -0.196 (-0.688 - 0.227) |
| $\theta_{123}$ - North Hertfordshire | 0.369 (0.0256 - 0.678) |
| $\theta_{124}$ - Watford | 0.157 (-0.215 - 0.525) |
| $\theta_{125}$ - Stevenage | 0.167 (-0.132 - 0.465) |
| $\theta_{126}$ - Tunbridge Wells | -0.129 (-0.577 - 0.255) |
| $\theta_{127}$ - Sevenoaks | 0.142 (-0.301 - 0.52) |
| $\theta_{128}$ - Tonbridge and Malling | -0.0563 (-0.557 - 0.349) |
| $\theta_{129}$ - Ashford | -0.025 (-0.384 - 0.301) |
| $\theta_{130}$ - Maidstone | -0.401 (-0.943 - 0.0338) |
| $\theta_{131}$ - Dartford | -0.316 (-0.985 - 0.243) |
| $\theta_{132}$ - Gravesham | 0.0785 (-0.488 - 0.596) |
| $\theta_{133}$ - Chorley | 0.672 (0.336 - 1.02) |
| $\theta_{134}$ - Rossendale | 0.222 (-0.128 - 0.553) |
| $\theta_{135}$ - Preston | 0.275 (-0.04 - 0.588) |
| $\theta_{136}$ - Pendle | -0.151 (-0.526 - 0.225) |
| $\theta_{137}$ - Burnley | -0.0129 (-0.428 - 0.376) |
| $\theta_{138}$ - Hinckley and Bosworth | -0.157 (-0.624 - 0.226) |
| $\theta_{139}$ - Melton | -0.325 (-0.755 - 0.0373) |
| $\theta_{140}$ - Harborough | 0.34 (0.0407 - 0.63) |
| $\theta_{141}$ - Blaby | 0.248 (-0.0461 - 0.529) |
| $\theta_{142}$ - Charnwood | 0.329 (0.0771 - 0.573) |
| $\theta_{143}$ - Oadby and Wigston | 0.304 (-0.319 - 0.918) |
| $\theta_{144}$ - West Lindsey | -0.195 (-0.589 - 0.148) |
| $\theta_{145}$ - South Kesteven | 0.321 (0.0123 - 0.598) |
| $\theta_{146}$ - South Holland | 0.118 (-0.159 - 0.375) |
| $\theta_{147}$ - Boston | -0.316 (-0.658 - -0.00845) |
| $\theta_{148}$ - North Kesteven | 0.37 (0.0611 - 0.664) |
| $\theta_{149}$ - Lincoln | 0.134 (-0.241 - 0.505) |
| $\theta_{150}$ - Breckland | -0.148 (-0.578 - 0.215) |
| $\theta_{151}$ - Norwich | -0.14 (-0.593 - 0.305) |
| $\theta_{152}$ - Craven | 0.473 (0.185 - 0.757) |
| $\theta_{153}$ - Bassetlaw | -0.303 (-0.735 - 0.0565) |
| $\theta_{154}$ - Gedling | 0.152 (-0.341 - 0.533) |
| $\theta_{155}$ - Ashfield | -0.128 (-0.506 - 0.215) |
| $\theta_{156}$ - Newark and Sherwood | 0.157 (-0.177 - 0.453) |
| $\theta_{157}$ - Broxtowe | -0.000549 (-0.629 - 0.455) |
| $\theta_{158}$ - Mansfield | -0.192 (-0.573 - 0.153) |
| $\theta_{159}$ - West Oxfordshire | -0.572 (-1.1 - -0.017) |
| $\theta_{160}$ - Oxford | 0.376 (0.00298 - 0.738) |
| $\theta_{161}$ - Mendip | -0.0173 (-0.358 - 0.29) |
| $\theta_{162}$ - Cannock Chase | -0.235 (-0.658 - 0.128) |
| $\theta_{163}$ - Mid Suffolk | -0.823 (-1.61 - -0.171) |
| $\theta_{164}$ - Ipswich | 0.0855 (-0.3 - 0.466) |
| $\theta_{165}$ - Waverley | 0.0666 (-0.444 - 0.488) |
| $\theta_{166}$ - Woking | 0.752 (0.453 - 1.05) |
| $\theta_{167}$ - Surrey Heath | -0.0446 (-0.752 - 0.485) |

Continued on next page

| Table S2 – continued from previous page | |
|---|---|
| Parameter | Posterior distribution mean (95% CrI) |
| $\theta_{168}$ - Runnymede | 0.123 (-0.354 - 0.522) |
| $\theta_{169}$ - Guildford | -0.343 (-0.95 - 0.284) |
| $\theta_{170}$ - Reigate and Banstead | 0.0881 (-0.398 - 0.5) |
| $\theta_{171}$ - Mole Valley | -0.944 (-1.66 - -0.119) |
| $\theta_{172}$ - Elmbridge | -0.167 (-0.823 - 0.364) |
| $\theta_{173}$ - Epsom and Ewell | -0.302 (-1.02 - 0.319) |
| $\theta_{174}$ - Rugby | 0.138 (-0.248 - 0.47) |
| $\theta_{175}$ - Nuneaton and Bedworth | -0.119 (-0.509 - 0.234) |
| $\theta_{176}$ - Mid Sussex | 0.287 (-0.121 - 0.642) |
| $\theta_{177}$ - Crawley | 0.0144 (-0.358 - 0.373) |
| $\theta_{178}$ - Malvern Hills | 0.0309 (-0.441 - 0.438) |
| $\theta_{179}$ - Wyre Forest | -0.136 (-0.544 - 0.224) |
| $\theta_{180}$ - Wychavon | -0.185 (-0.777 - 0.252) |
| $\theta_{181}$ - Bromsgrove | 0.587 (0.332 - 0.835) |
| $\theta_{182}$ - Worcester | 0.0398 (-0.344 - 0.413) |
| $\theta_{183}$ - Redditch | -0.0785 (-0.434 - 0.25) |
| $\theta_{184}$ - Broadland | -0.0329 (-0.569 - 0.412) |
| $\theta_{185}$ - South Lakeland | -0.429 (-1.05 - 0.0731) |
| $\theta_{186}$ - Copeland | -0.0708 (-0.429 - 0.263) |
| $\theta_{187}$ - Barrow-in-Furness | 0.0571 (-0.277 - 0.385) |
| $\theta_{188}$ - South Derbyshire | -0.256 (-0.922 - 0.211) |
| $\theta_{189}$ - Bolsover | -0.244 (-0.703 - 0.135) |
| $\theta_{190}$ - Derbyshire Dales | 0.514 (0.226 - 0.807) |
| $\theta_{191}$ - Teignbridge | 0.058 (-0.41 - 0.441) |
| $\theta_{192}$ - West Devon | 0.0655 (-0.358 - 0.471) |
| $\theta_{193}$ - Wealden | -0.497 (-0.985 - 0.114) |
| $\theta_{194}$ - Eastbourne | 0.204 (-0.152 - 0.547) |
| $\theta_{195}$ - Hastings | 0.0683 (-0.374 - 0.515) |
| $\theta_{196}$ - Rochford | 0.00367 (-0.747 - 0.513) |
| $\theta_{197}$ - Tendring | -0.0204 (-0.523 - 0.473) |
| $\theta_{198}$ - Colchester | 0.0536 (-0.282 - 0.373) |
| $\theta_{199}$ - Maldon | -0.154 (-0.733 - 0.286) |
| $\theta_{200}$ - Braintree | 0.142 (-0.125 - 0.397) |
| $\theta_{201}$ - Basildon | -0.0652 (-0.463 - 0.294) |
| $\theta_{202}$ - Castle Point | -0.343 (-0.956 - 0.157) |
| $\theta_{203}$ - Exeter | 0.502 (0.214 - 0.791) |
| $\theta_{204}$ - Lewes | -0.0526 (-0.677 - 0.409) |
| $\theta_{205}$ - Rother | 0.331 (-0.228 - 0.838) |
| $\theta_{206}$ - Hertsmere | -0.542 (-1.54 - 0.143) |
| $\theta_{207}$ - Folkestone and Hythe | -0.248 (-0.673 - 0.122) |
| $\theta_{208}$ - Thanet | 0.053 (-0.381 - 0.48) |
| $\theta_{209}$ - Canterbury | 0.299 (0.014 - 0.573) |
| $\theta_{210}$ - Dover | -0.221 (-0.594 - 0.0944) |
| $\theta_{211}$ - Swale | 0.000679 (-0.318 - 0.303) |
| $\theta_{212}$ - West Lancashire | 0.154 (-0.328 - 0.592) |
| $\theta_{213}$ - Lancaster | 0.286 (-0.0101 - 0.578) |
| $\theta_{214}$ - South Ribble | 0.619 (0.25 - 0.987) |
| $\theta_{215}$ - Fylde | -0.0973 (-0.854 - 0.528) |
| $\theta_{216}$ - Wyre | 0.502 (0.0946 - 0.892) |
| $\theta_{217}$ - Ribble Valley | 0.648 (0.192 - 1.11) |
| $\theta_{218}$ - Hyndburn | -0.651 (-1.16 - -0.199) |
| $\theta_{219}$ - North West Leicestershire | -0.014 (-0.386 - 0.305) |
| $\theta_{220}$ - East Lindsey | -0.179 (-0.67 - 0.294) |
| $\theta_{221}$ - King's Lynn and West Norfolk | -0.272 (-0.786 - 0.165) |
| $\theta_{222}$ - Selby | 0.117 (-0.235 - 0.442) |
| $\theta_{223}$ - Richmondshire | 0.0802 (-0.31 - 0.42) |
| $\theta_{224}$ - Harrogate | 0.614 (0.404 - 0.821) |
| $\theta_{225}$ - West Suffolk | -0.317 (-0.83 - 0.095) |
| $\theta_{226}$ - Tandridge | -0.241 (-1.2 - 0.385) |
| $\theta_{227}$ - Spelthorne | 0.166 (-0.266 - 0.538) |
| $\theta_{228}$ - North Warwickshire | 0.203 (-0.0592 - 0.442) |
| $\theta_{229}$ - Warwick | 0.449 (0.162 - 0.721) |
| $\theta_{230}$ - Stratford-on-Avon | 0.216 (-0.206 - 0.584) |
| $\theta_{231}$ - Arun | 0.254 (-0.154 - 0.634) |
| $\theta_{232}$ - Adur | -0.282 (-0.929 - 0.187) |
| $\theta_{233}$ - Chichester | -0.487 (-1.11 - 0.0706) |
| $\theta_{234}$ - Horsham | 0.299 (-0.116 - 0.655) |
| $\theta_{235}$ - Worthing | -0.199 (-0.574 - 0.194) |
| $\theta_{236}$ - Kingston upon Thames | 0.434 (-0.0425 - 0.889) |
| $\theta_{237}$ - Croydon | 0.32 (-0.417 - 1.04) |
| $\theta_{238}$ - Bromley | 0.216 (-0.359 - 0.642) |
| $\theta_{239}$ - Hounslow | -0.148 (-0.752 - 0.428) |

Continued on next page

| Table S2 – continued from previous page | |
|---|---|
| Parameter | Posterior distribution mean (95% CrI) |
| $\theta_{240}$ - Ealing | -0.403 (-1.13 - 0.365) |
| $\theta_{241}$ - Havering | -0.027 (-0.55 - 0.369) |
| $\theta_{242}$ - Hillingdon | 0.0898 (-0.508 - 0.642) |
| $\theta_{243}$ - Harrow | -0.255 (-1.15 - 0.572) |
| $\theta_{244}$ - Brent | 0.295 (-0.389 - 0.966) |
| $\theta_{245}$ - Barnet | 0.182 (-0.405 - 0.754) |
| $\theta_{246}$ - Lambeth | 0.473 (-0.434 - 1.35) |
| $\theta_{247}$ - Southwark | 0.244 (-0.708 - 1.13) |
| $\theta_{248}$ - Lewisham | -0.0027 (-0.866 - 0.872) |
| $\theta_{249}$ - Greenwich | 0.125 (-0.616 - 0.773) |
| $\theta_{250}$ - Bexley | -0.535 (-1.18 - 0.189) |
| $\theta_{251}$ - Enfield | -0.0621 (-0.829 - 0.653) |
| $\theta_{252}$ - Waltham Forest | 0.0818 (-0.722 - 0.706) |
| $\theta_{253}$ - Redbridge | -0.619 (-1.37 - 0.153) |
| $\theta_{254}$ - Sutton | -0.563 (-1.11 - 0.0367) |
| $\theta_{255}$ - Richmond upon Thames | -0.0195 (-0.565 - 0.441) |
| $\theta_{256}$ - Merton | 0.55 (0.096 - 0.968) |
| $\theta_{257}$ - Wandsworth | -0.02 (-0.751 - 0.63) |
| $\theta_{258}$ - Hammersmith and Fulham | -0.114 (-0.907 - 0.564) |
| $\theta_{259}$ - Kensington and Chelsea | -0.573 (-1.58 - 0.402) |
| $\theta_{260}$ - Westminster | 0.116 (-0.862 - 1.1) |
| $\theta_{261}$ - Camden | -0.57 (-1.46 - 0.234) |
| $\theta_{262}$ - Tower Hamlets | -0.16 (-1.25 - 0.923) |
| $\theta_{263}$ - Islington | 0.247 (-0.609 - 1.1) |
| $\theta_{264}$ - Hackney and City of London | 0.213 (-0.546 - 1.01) |
| $\theta_{265}$ - Haringey | -0.919 (-1.62 - -0.109) |
| $\theta_{266}$ - Newham | -0.0978 (-0.8 - 0.571) |
| $\theta_{267}$ - Barking and Dagenham | -0.642 (-1.55 - 0.274) |
| $\theta_{268}$ - North Norfolk | 0.0687 (-0.452 - 0.56) |
| $\theta_{269}$ - Rushcliffe | 0.498 (0.109 - 0.886) |
| $\theta_{270}$ - Sedgemoor | -0.104 (-0.464 - 0.226) |
| $\theta_{271}$ - Staffordshire Moorlands | -0.023 (-0.445 - 0.329) |
| $\theta_{272}$ - South Staffordshire | -0.0339 (-0.511 - 0.357) |
| $\theta_{273}$ - Lichfield | 0.494 (0.251 - 0.726) |
| $\theta_{274}$ - Newcastle-under-Lyme | -0.513 (-1.1 - -0.0787) |
| $\theta_{275}$ - Stafford | -0.0434 (-0.488 - 0.318) |
| $\theta_{276}$ - East Staffordshire | -0.04 (-0.354 - 0.243) |
| $\theta_{277}$ - Tamworth | -0.478 (-0.789 - -0.128) |
| $\theta_{278}$ - Allerdale | -0.03 (-0.336 - 0.266) |
| $\theta_{279}$ - South Hams | 0.00801 (-0.41 - 0.385) |
| $\theta_{280}$ - Torridge | -0.0201 (-0.378 - 0.334) |
| $\theta_{281}$ - North Devon | 0.215 (-0.0501 - 0.477) |
| $\theta_{282}$ - South Somerset | 0.14 (-0.176 - 0.435) |
| $\theta_{283}$ - Mid Devon | -0.307 (-0.792 - 0.0905) |
| $\theta_{284}$ - East Devon | 0.122 (-0.336 - 0.553) |
| $\theta_{285}$ - Ryedale | 0.283 (-0.0416 - 0.588) |
| $\theta_{286}$ - Hambleton | -0.133 (-0.592 - 0.262) |
| $\theta_{287}$ - Scarborough | -0.244 (-0.715 - 0.198) |
| $\theta_{288}$ - Eastleigh | -0.171 (-0.819 - 0.314) |
| $\theta_{289}$ - Stroud | -0.126 (-0.656 - 0.288) |
| $\theta_{290}$ - Forest of Dean | 0.179 (-0.145 - 0.471) |
| $\theta_{291}$ - Winchester | -0.425 (-1.12 - 0.104) |
| $\theta_{292}$ - Test Valley | -0.845 (-1.71 - -0.109) |
| $\theta_{293}$ - Havant | 0.0613 (-0.315 - 0.422) |
| $\theta_{294}$ - East Hampshire | -0.39 (-1.14 - 0.205) |
| $\theta_{295}$ - Basingstoke and Deane | 0.227 (-0.138 - 0.553) |
| $\theta_{296}$ - Fareham | -0.355 (-1 - 0.224) |
| $\theta_{297}$ - Vale of White Horse | -0.812 (-1.67 - -0.0243) |
| $\theta_{298}$ - Cherwell | -0.161 (-0.628 - 0.234) |
| $\theta_{299}$ - Carlisle | 0.148 (-0.108 - 0.4) |
| $\theta_{300}$ - Eden | 0.214 (-0.0501 - 0.473) |
| $\theta_{301}$ - Great Yarmouth | -0.201 (-0.673 - 0.268) |
| $\theta_{302}$ - East Suffolk | -0.0611 (-0.531 - 0.352) |
| $\theta_{303}$ - South Norfolk | 0.287 (-0.0572 - 0.594) |
| $\theta_{304}$ - Babergh | -0.398 (-1.09 - 0.0956) |
| $\theta_{305}$ - Somerset West and Taunton | -0.00298 (-0.322 - 0.292) |
| $\theta_{306}$ - South Oxfordshire | -0.0844 (-0.512 - 0.287) |

We demonstrate the variation across mean $\zeta_i$ and $\theta_i$ values in Figure S34. The median value of $\zeta_i$ is 0.134 (interquartile range 0.069 - 0.247). The median value of $\theta_i$ is 0.0282 (interquartile range -0.159 - 0.222).

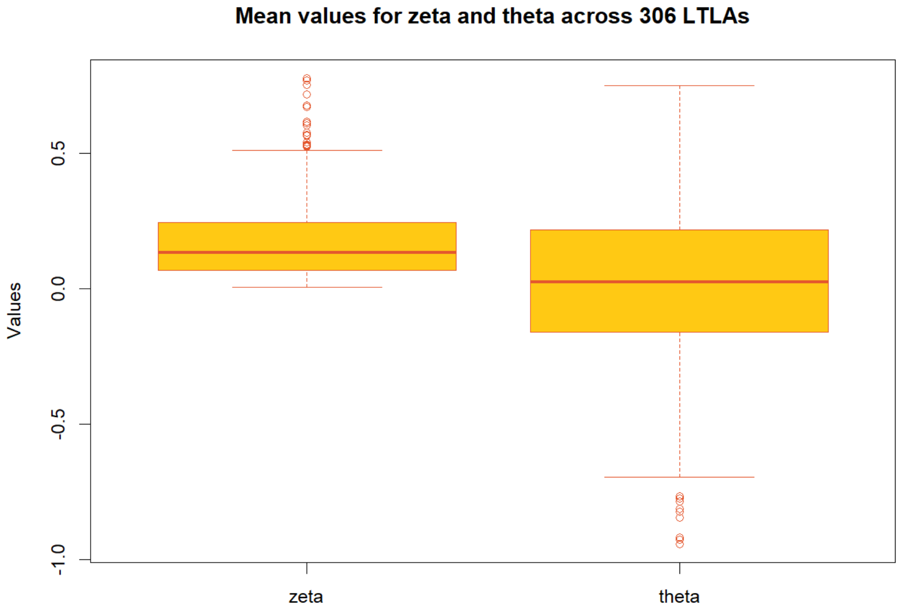

**Figure S34:** Boxplots of the mean posterior value across all 306 LTLAs for parameters $\zeta$ and $\theta$, as listed above in Table S2.

### 3.4 Assessing convergence

Model convergence was assessed via the Gelman-Rubin diagnostic (ensuring all parameters had potential scale reduction factor $< 1.1$) and sampling sufficiency was assessed by visual inspection of the posterior distributions and trace plots. Traceplots for all model variables are provided below in Figures S35 to S59.

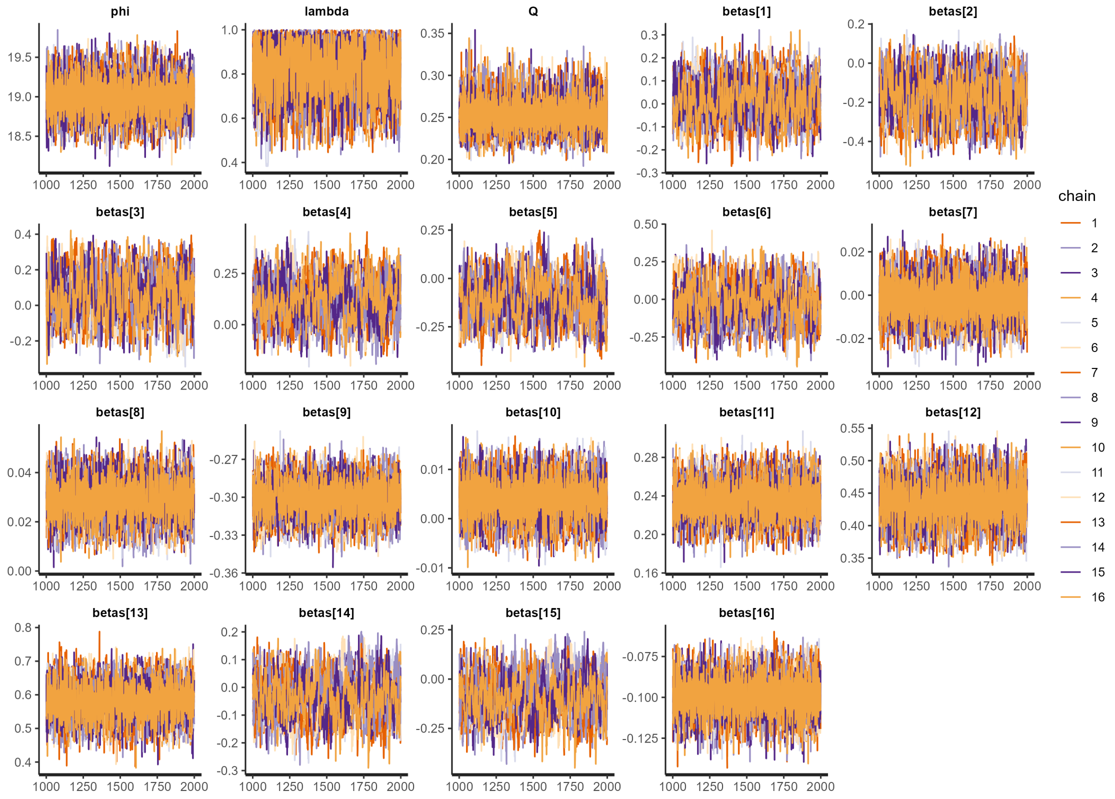

**Figure S35:** Traceplots for the post-burn-in iterations sampling the posterior distributions of $\phi$, $\lambda$, $Q$, and $\beta$.

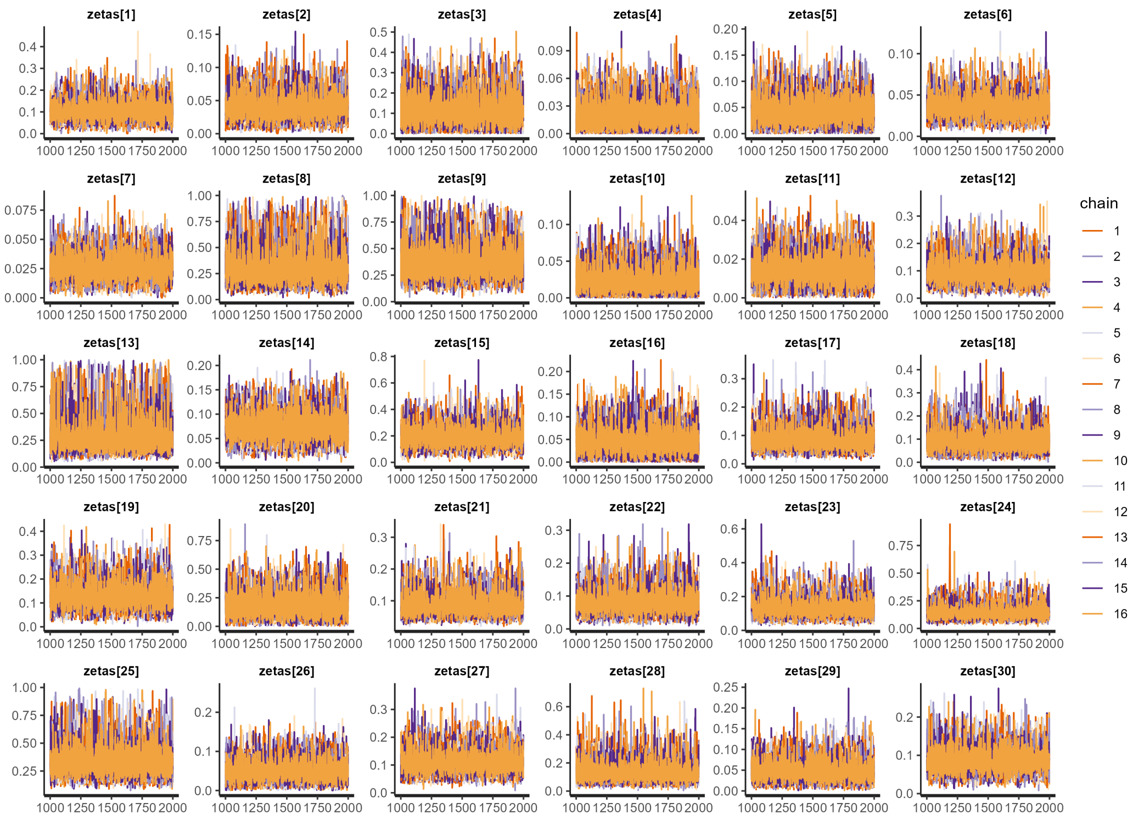

**Figure S36:** Traceplots for the post-burn-in iterations sampling the posterior distributions of $\zeta_1$ to $\zeta_{30}$.

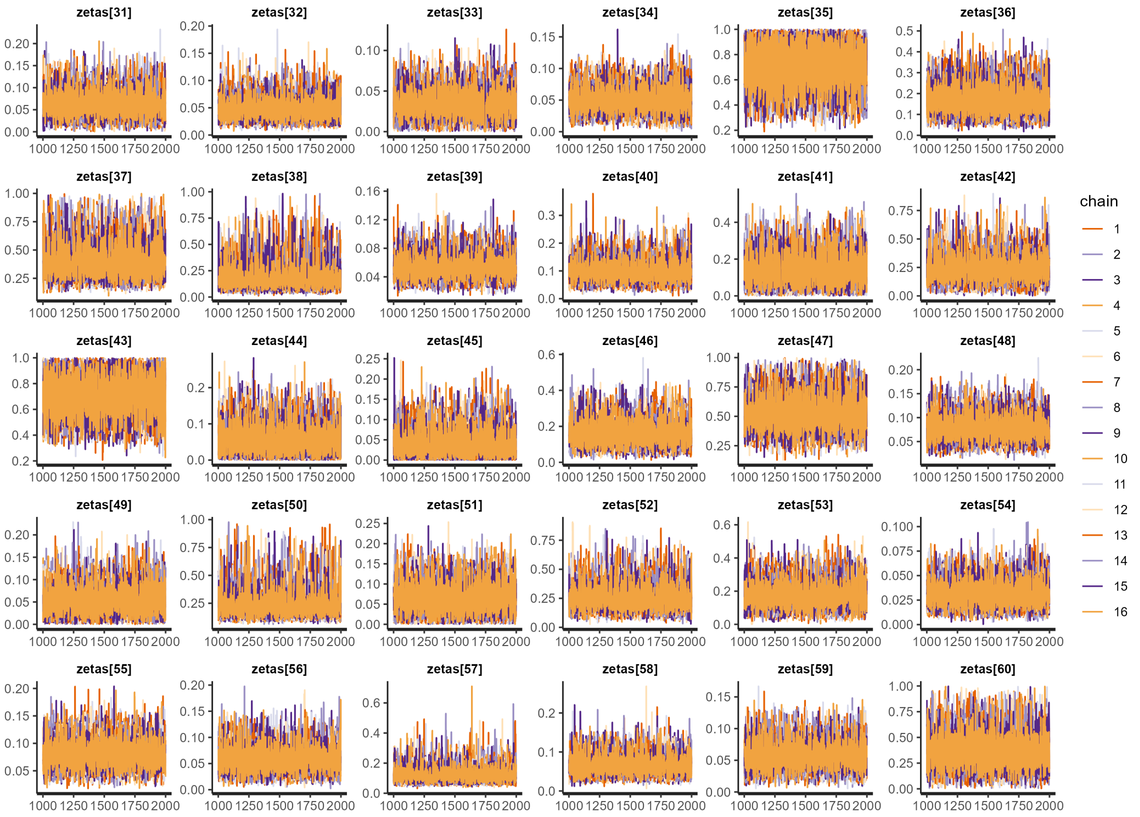

**Figure S37:** Traceplots for the post-burn-in iterations sampling the posterior distributions of $\zeta_{31}$ to $\zeta_{60}$.

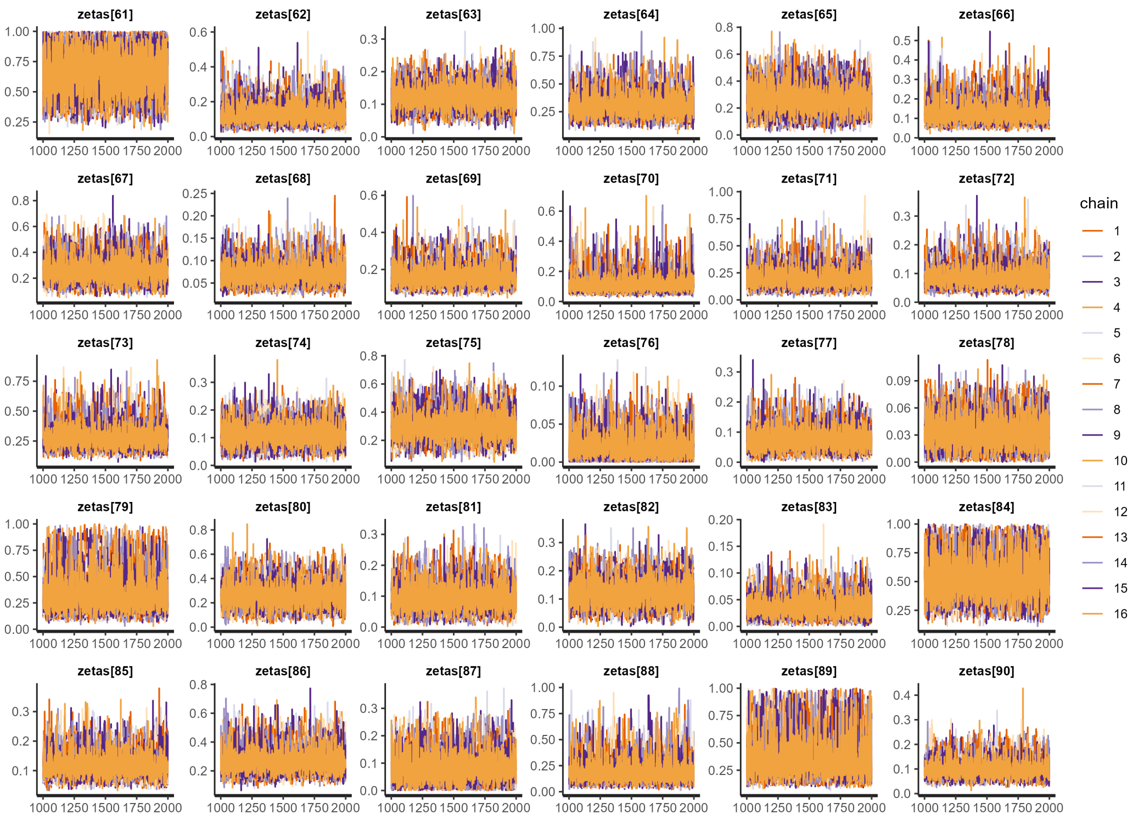

**Figure S38:** Traceplots for the post-burn-in iterations sampling the posterior distributions of $\zeta_{61}$ to $\zeta_{90}$.

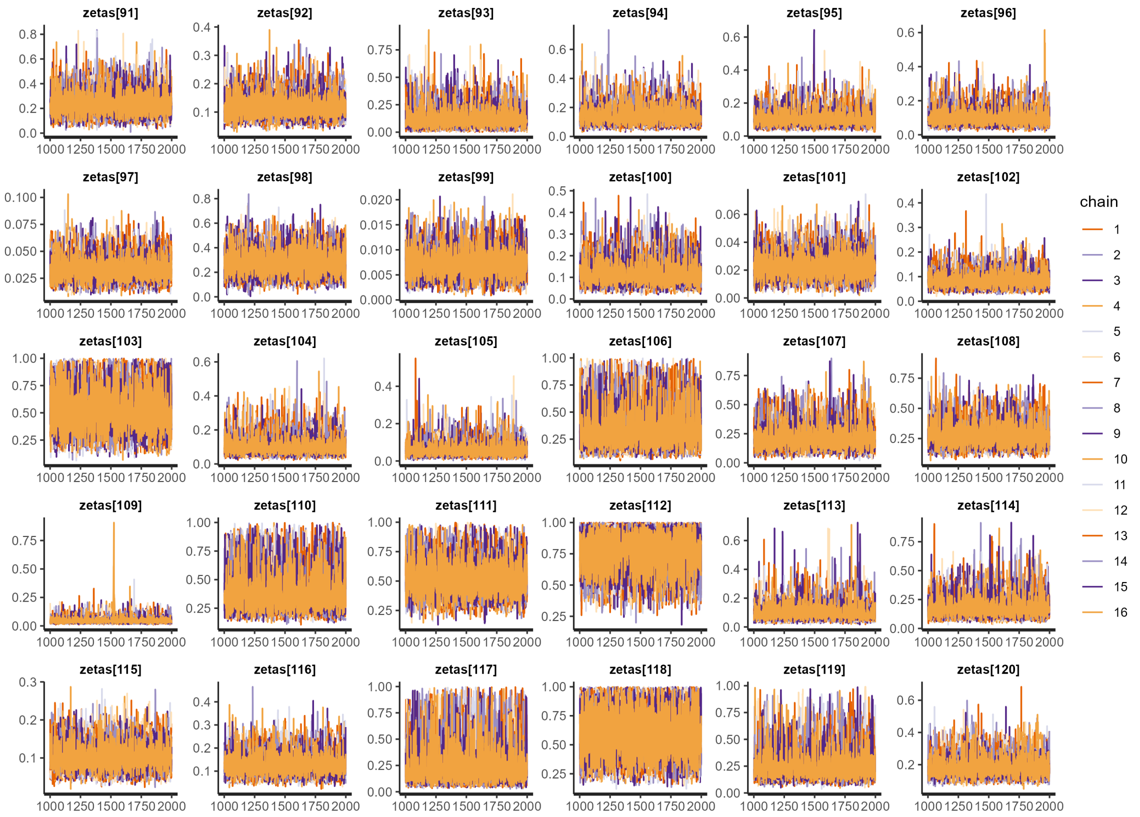

**Figure S39:** Traceplots for the post-burn-in iterations sampling the posterior distributions of $\zeta_{91}$ to $\zeta_{120}$.

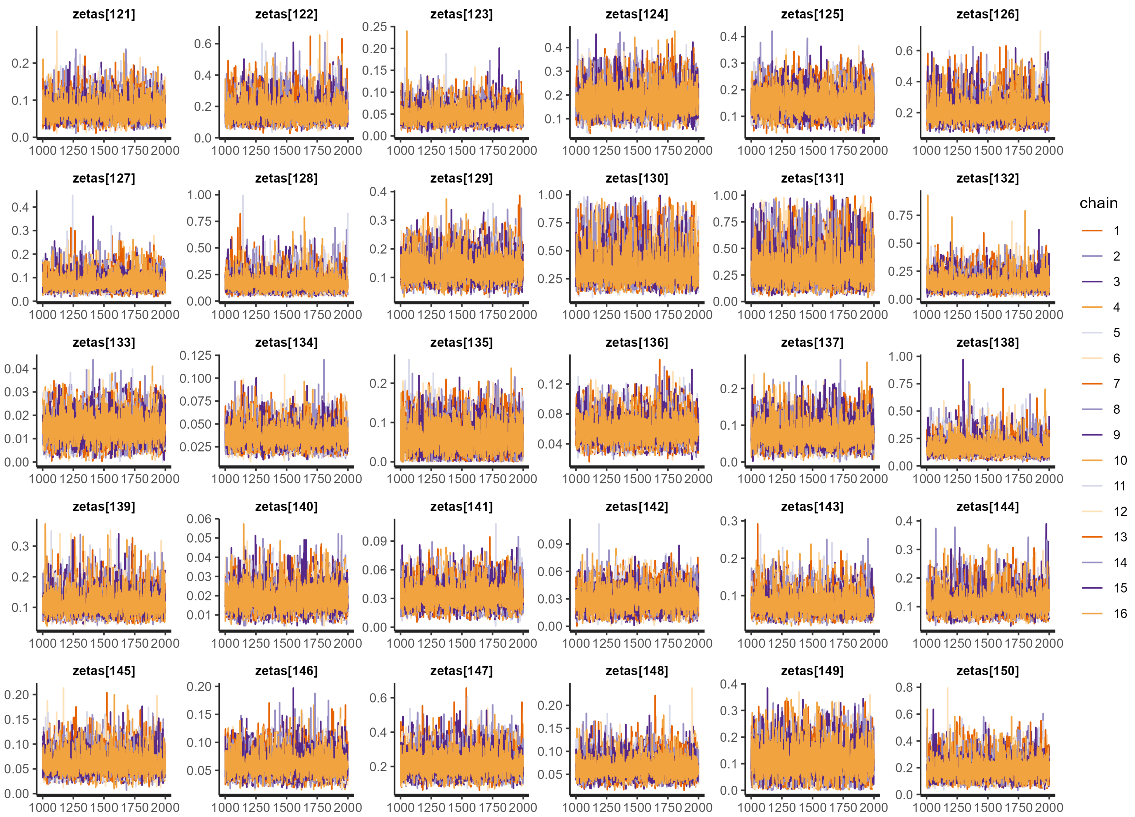

**Figure S40:** Traceplots for the post-burn-in iterations sampling the posterior distributions of $\zeta_{121}$ to $\zeta_{150}$.

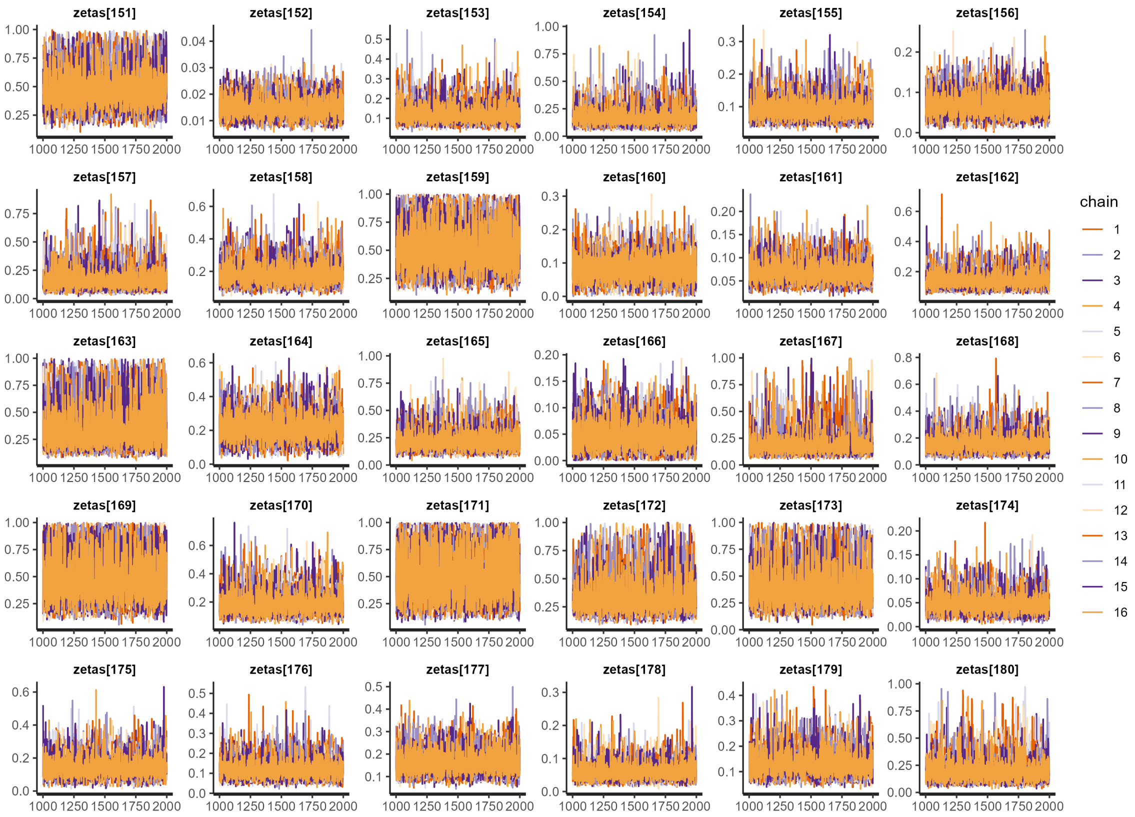

**Figure S41:** Traceplots for the post-burn-in iterations sampling the posterior distributions of $\zeta_{151}$ to $\zeta_{180}$.

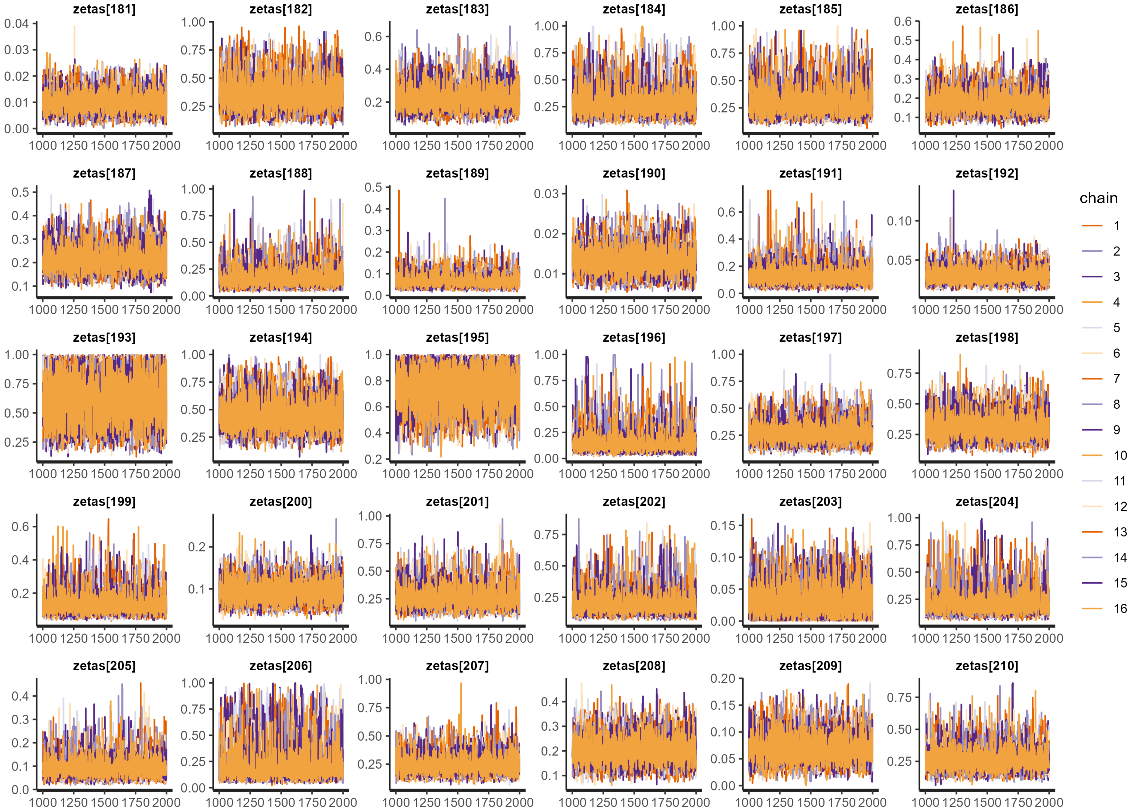

**Figure S42:** Traceplots for the post-burn-in iterations sampling the posterior distributions of $\zeta_{181}$ to $\zeta_{210}$.

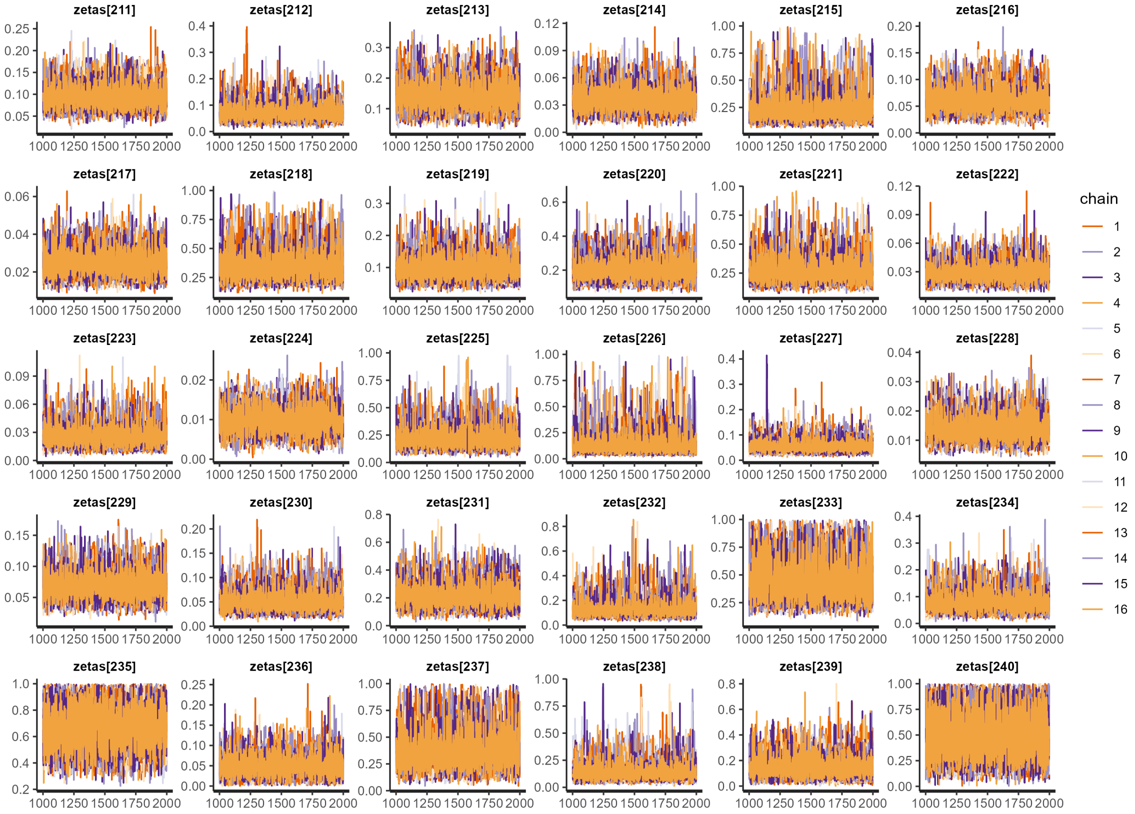

**Figure S43:** Traceplots for the post-burn-in iterations sampling the posterior distributions of $\zeta_{211}$ to $\zeta_{240}$.

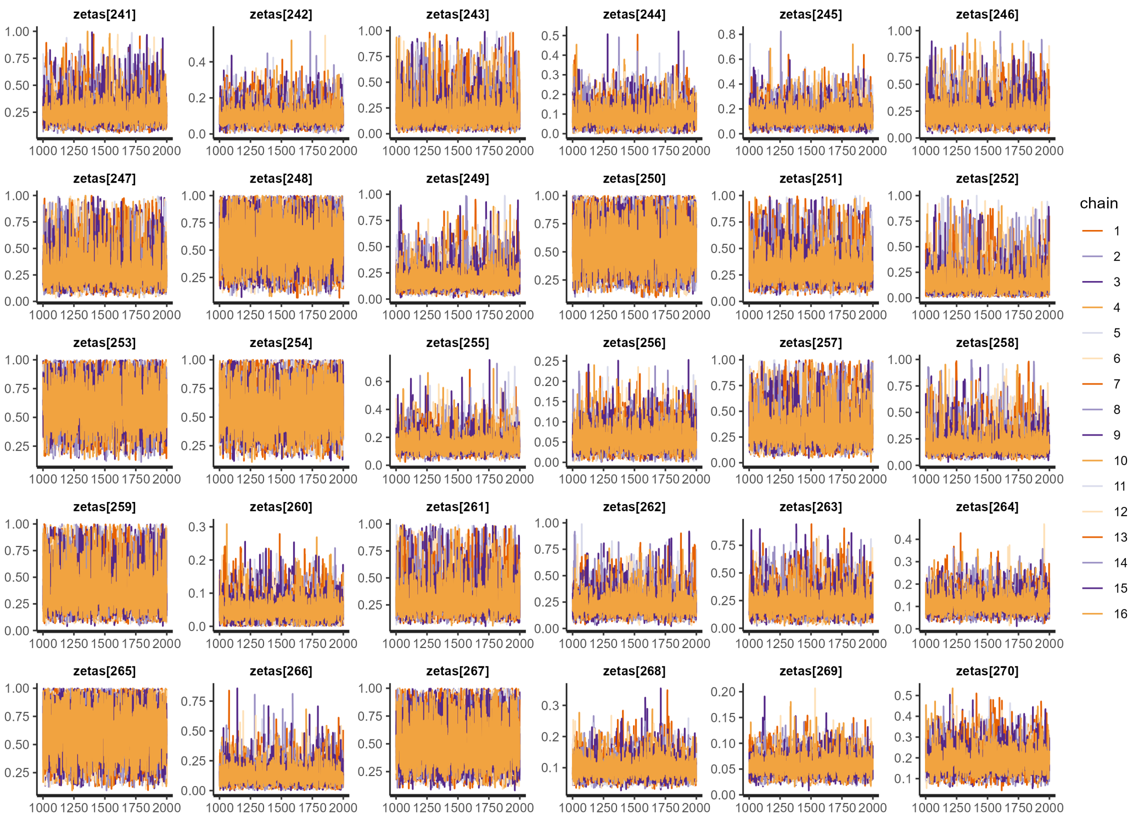

**Figure S44:** Traceplots for the post-burn-in iterations sampling the posterior distributions of $\zeta_{241}$ to $\zeta_{270}$.

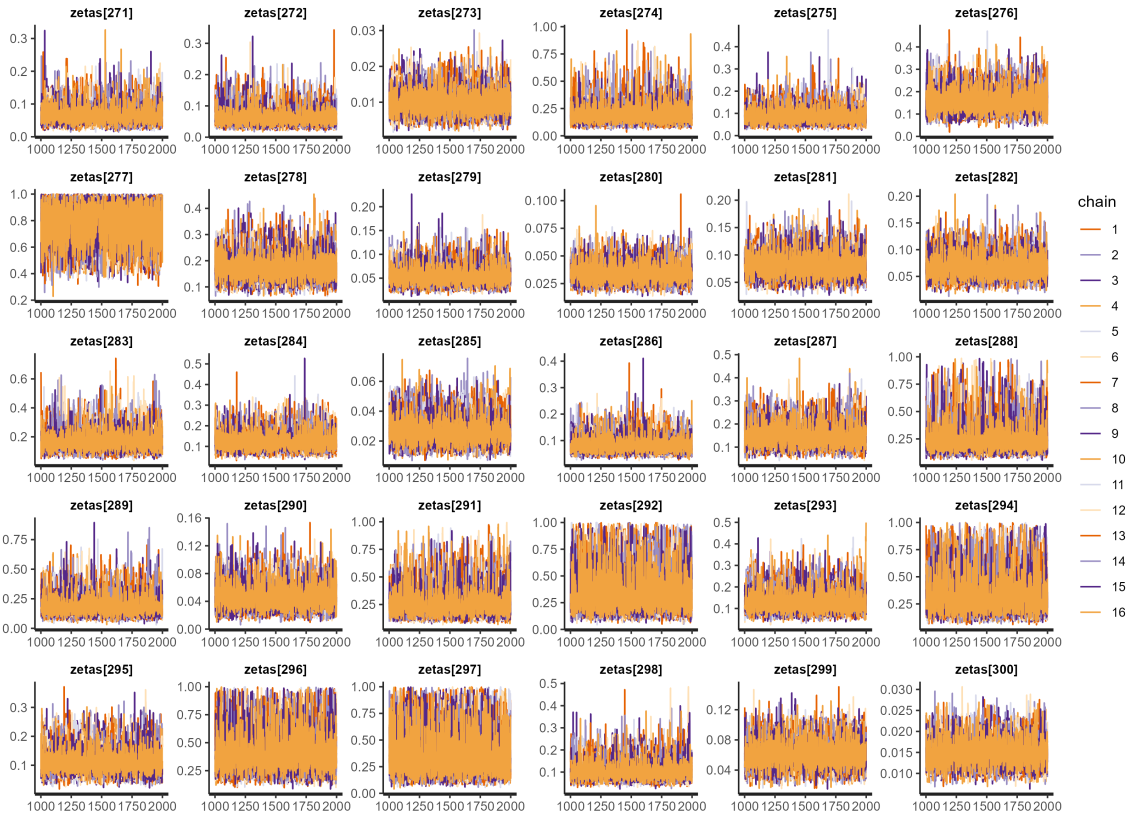

**Figure S45:** Traceplots for the post-burn-in iterations sampling the posterior distributions of $\zeta_{271}$ to $\zeta_{300}$.

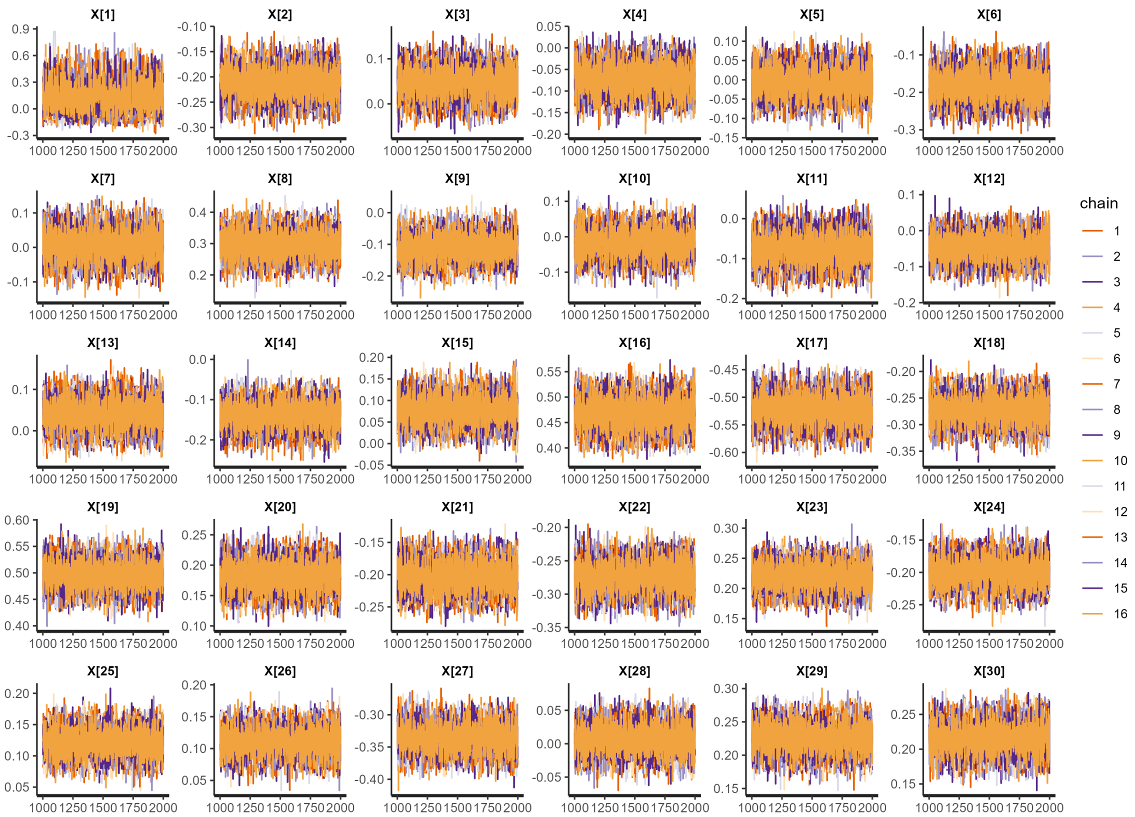

**Figure S46:** Traceplots for the post-burn-in iterations sampling the posterior distributions of $X_1$ to $X_{30}$.

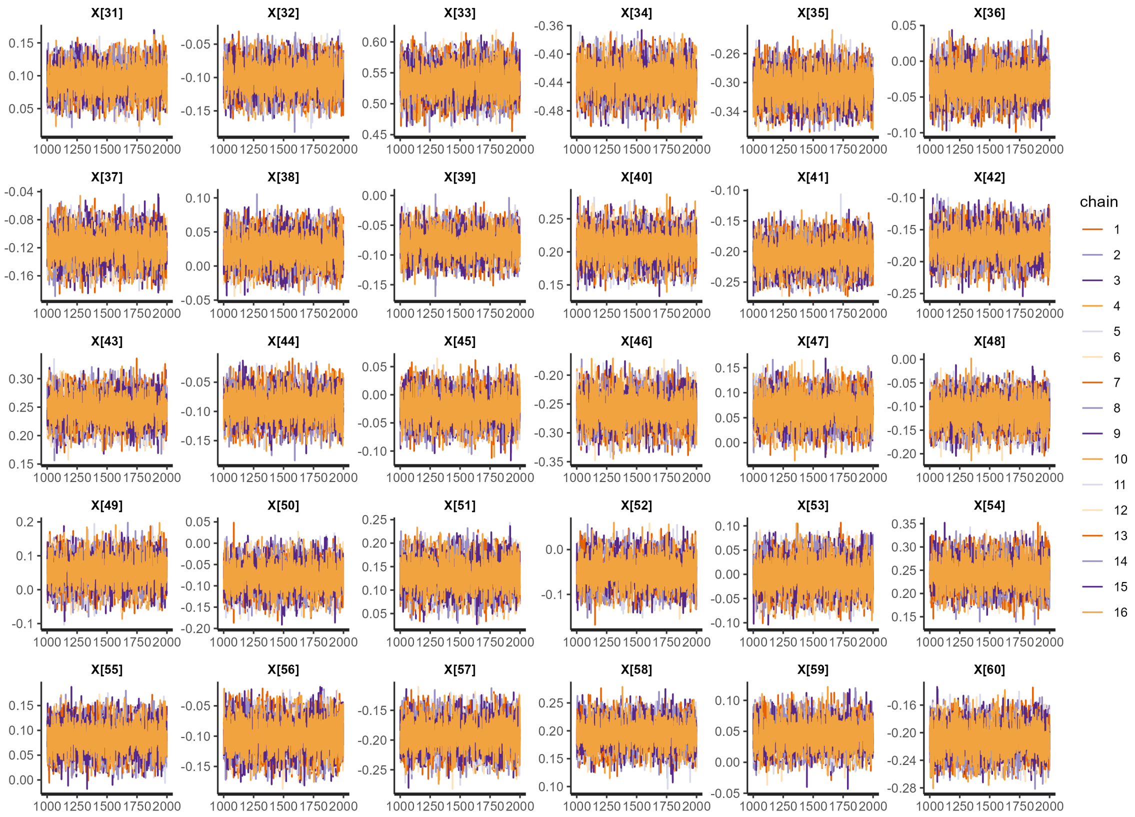

**Figure S47:** Traceplots for the post-burn-in iterations sampling the posterior distributions of $X_{31}$ to $X_{60}$.

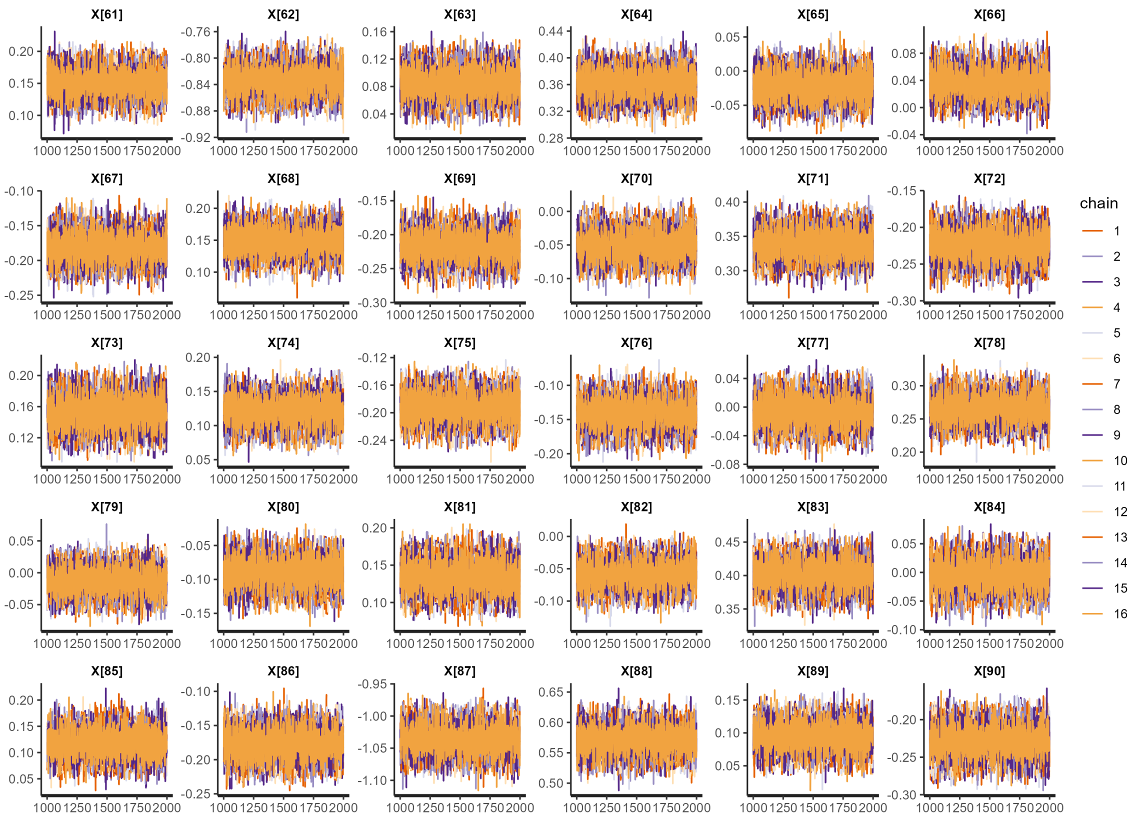

**Figure S48:** Traceplots for the post-burn-in iterations sampling the posterior distributions of $X_{61}$ to $X_{90}$.

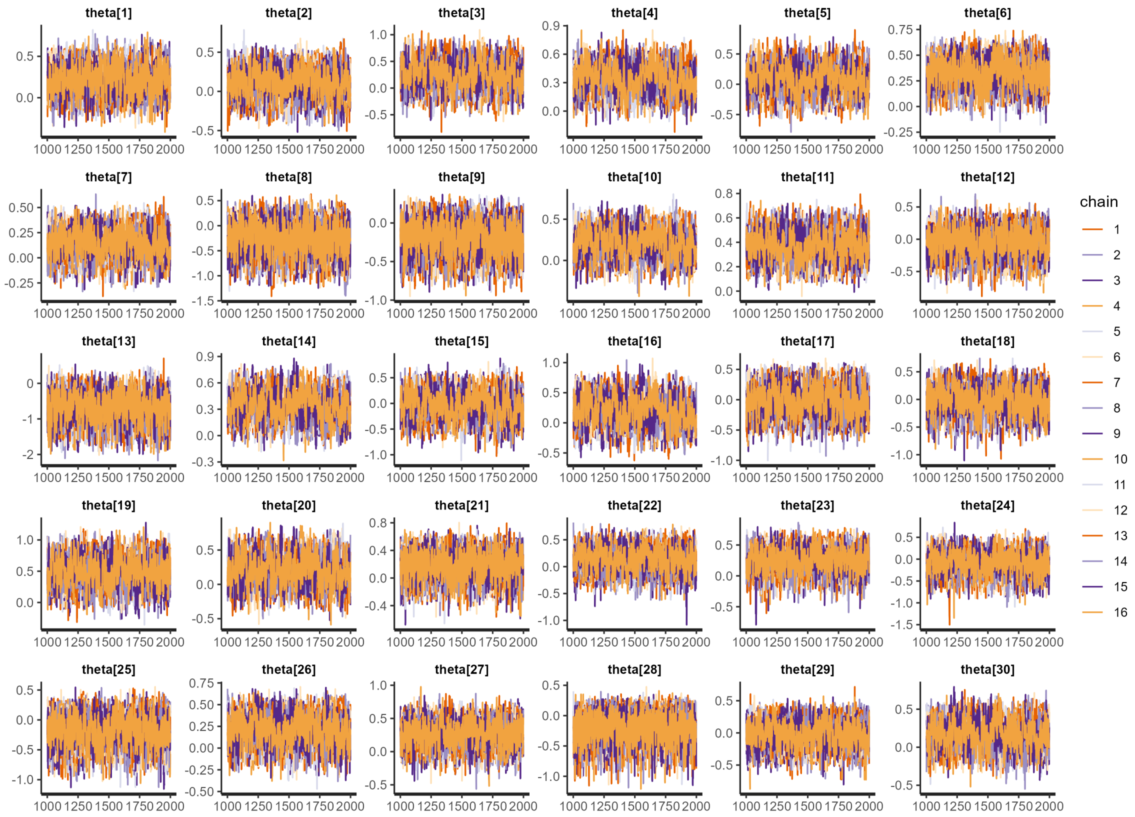

**Figure S49:** Traceplots for the post-burn-in iterations sampling the posterior distributions of $\theta_1$ to $\theta_{30}$.

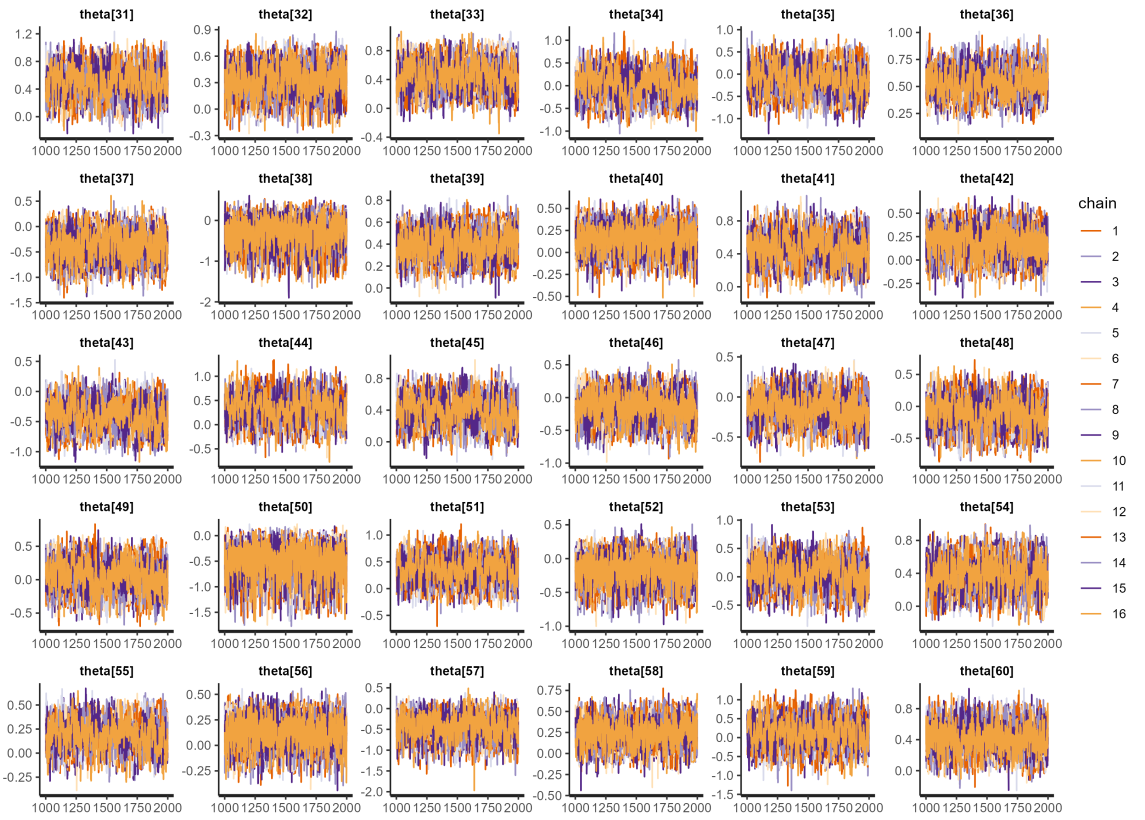

**Figure S50:** Traceplots for the post-burn-in iterations sampling the posterior distributions of $\theta_{31}$ to $\theta_{60}$.

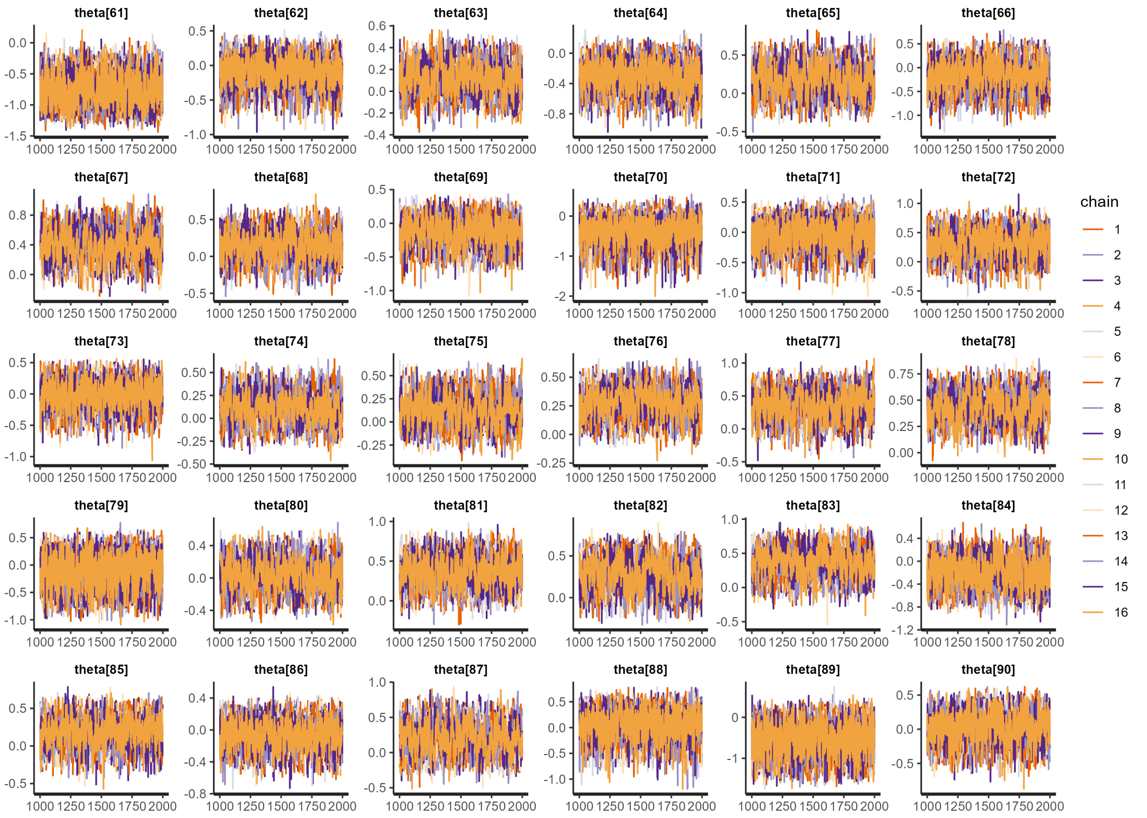

**Figure S51:** Traceplots for the post-burn-in iterations sampling the posterior distributions of $\theta_{61}$ to $\theta_{90}$.

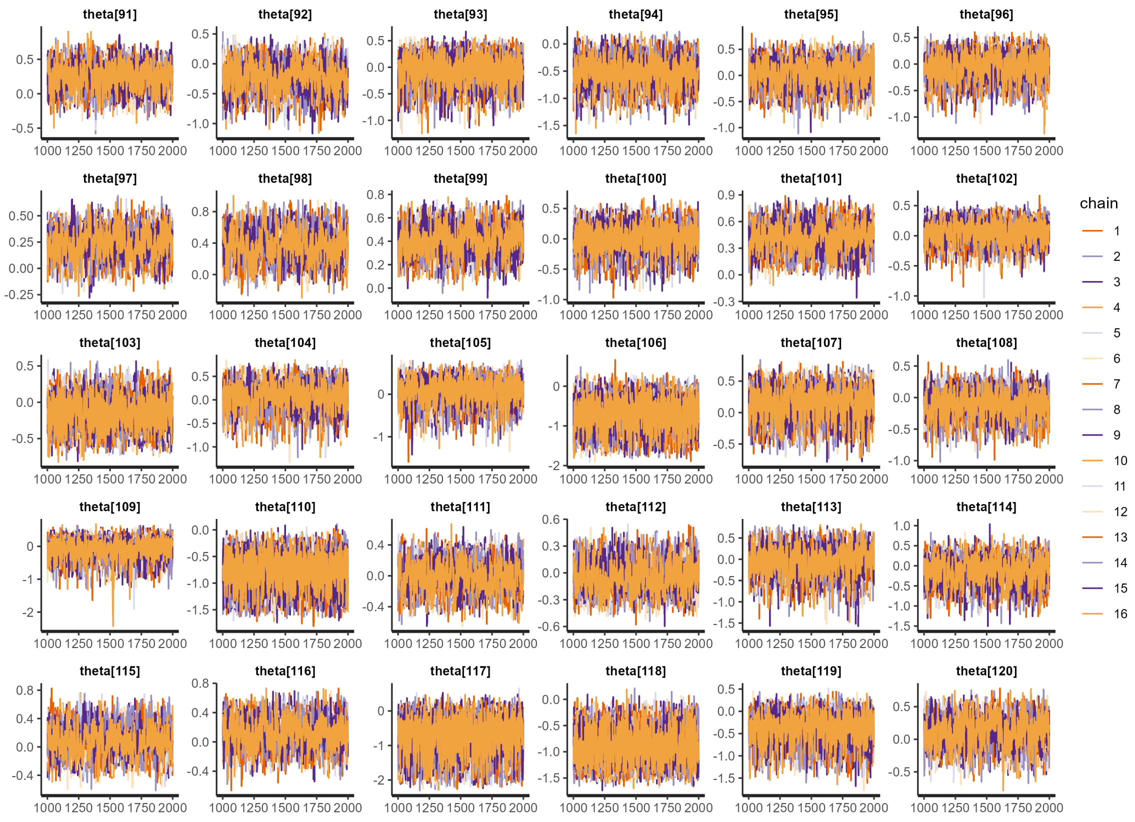

**Figure S52:** Traceplots for the post-burn-in iterations sampling the posterior distributions of $\theta_{91}$ to $\theta_{120}$.

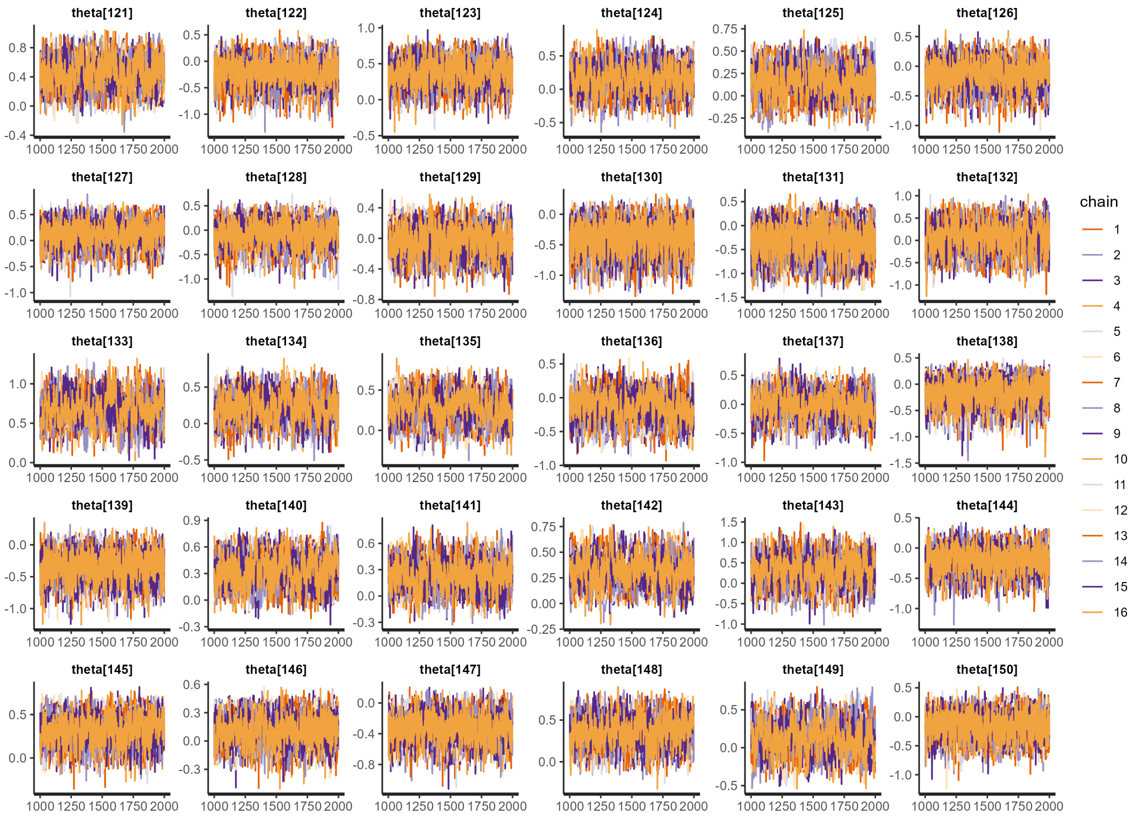

**Figure S53:** Traceplots for the post-burn-in iterations sampling the posterior distributions of $\theta_{121}$ to $\theta_{150}$.

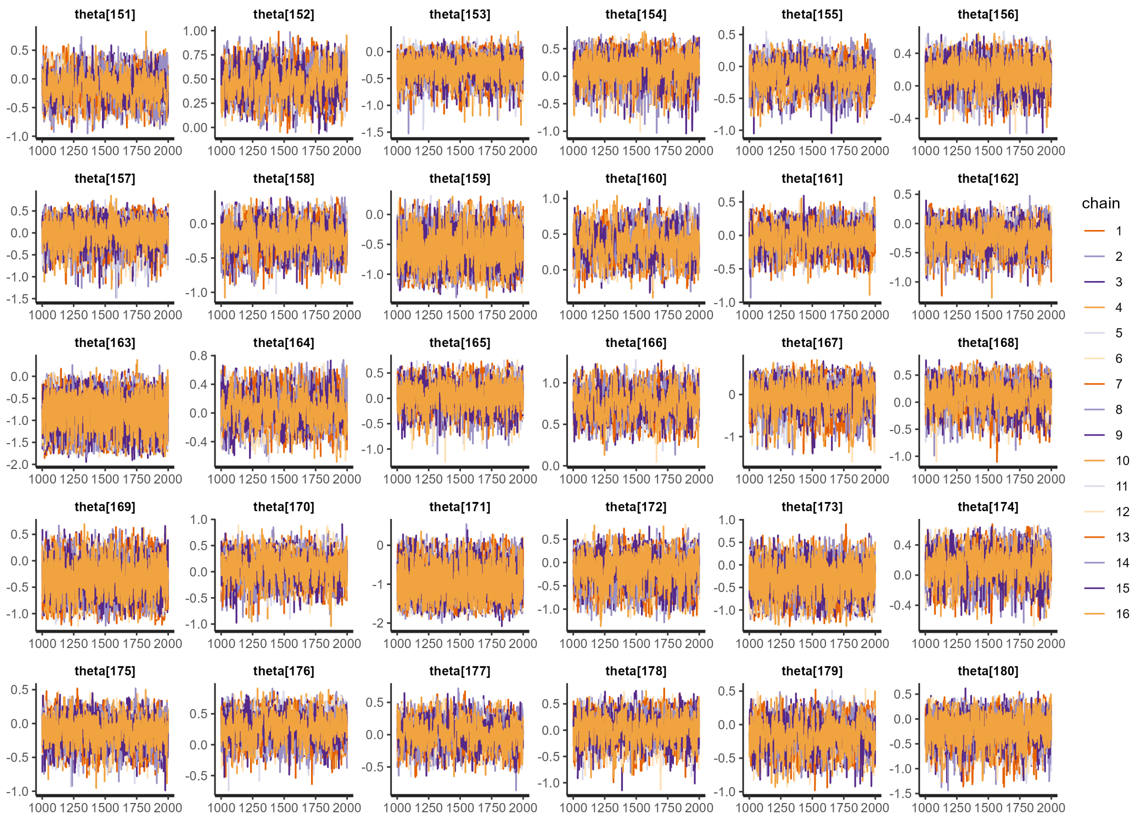

**Figure S54:** Traceplots for the post-burn-in iterations sampling the posterior distributions of $\theta_{151}$ to $\theta_{180}$.

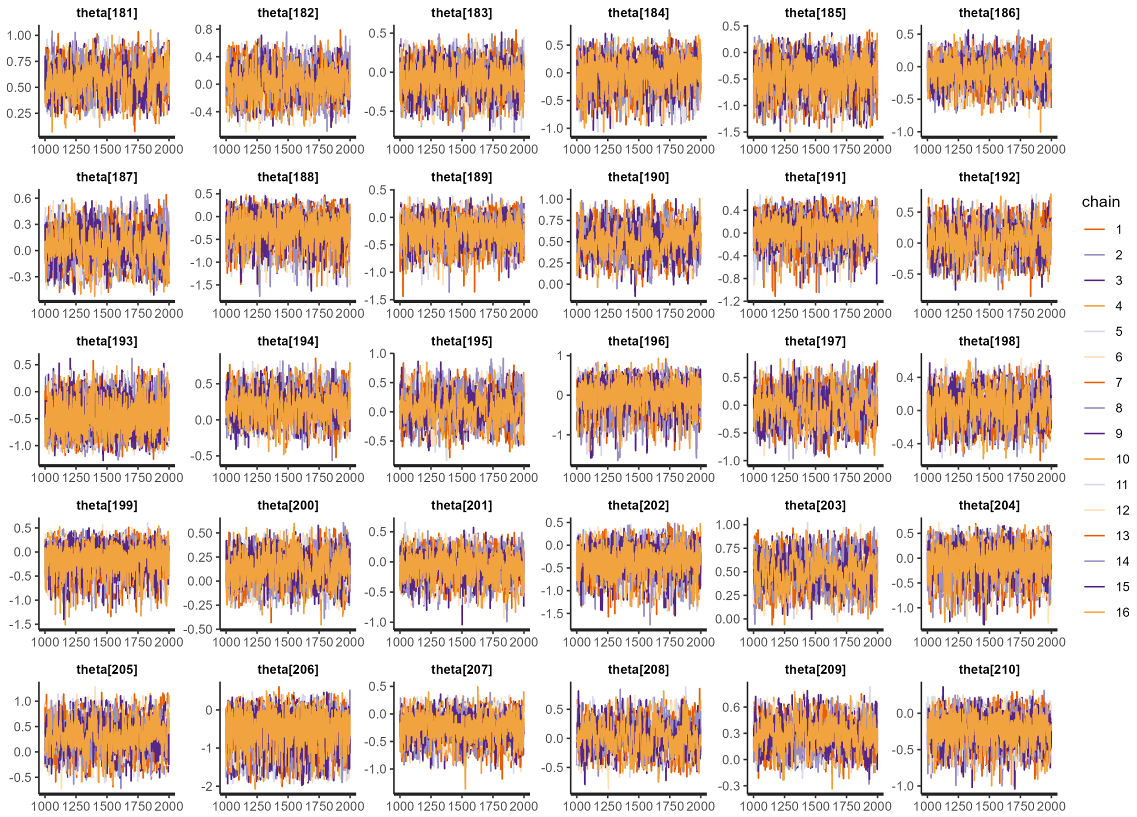

**Figure S55:** Traceplots for the post-burn-in iterations sampling the posterior distributions of $\theta_{181}$ to $\theta_{210}$.

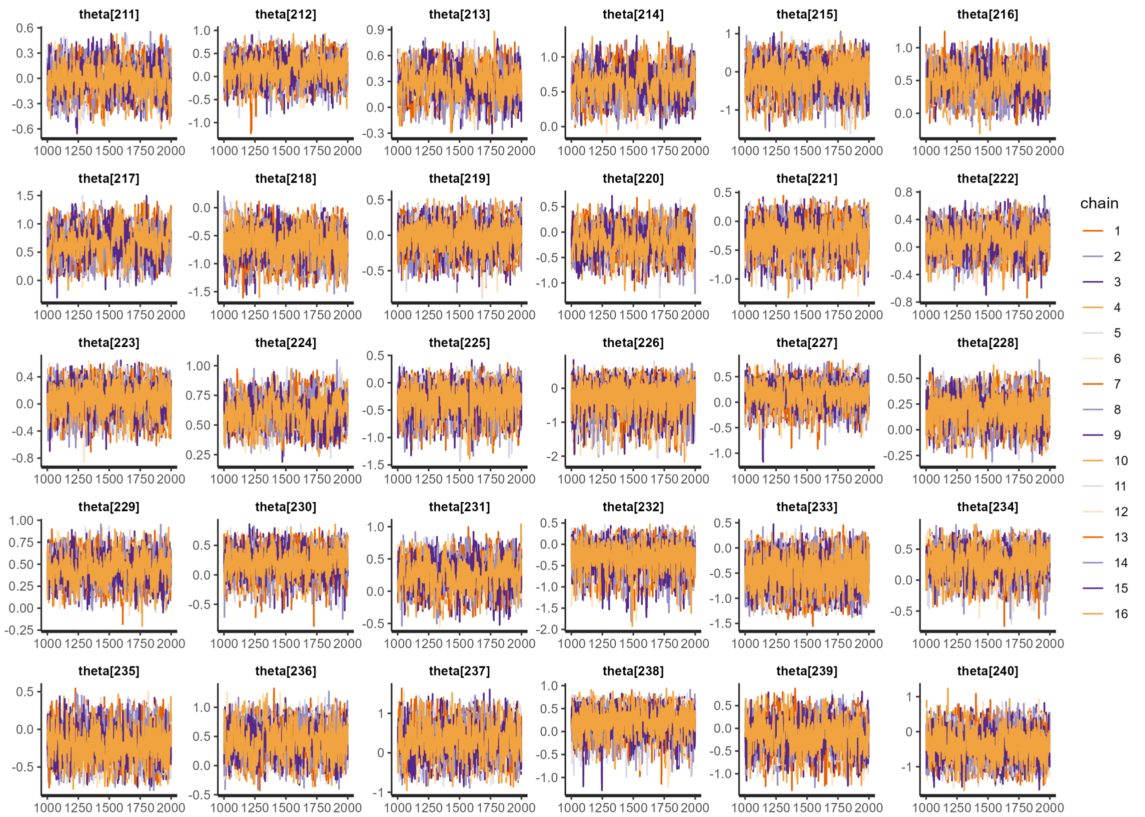

**Figure S56:** Traceplots for the post-burn-in iterations sampling the posterior distributions of $\theta_{211}$ to $\theta_{240}$.

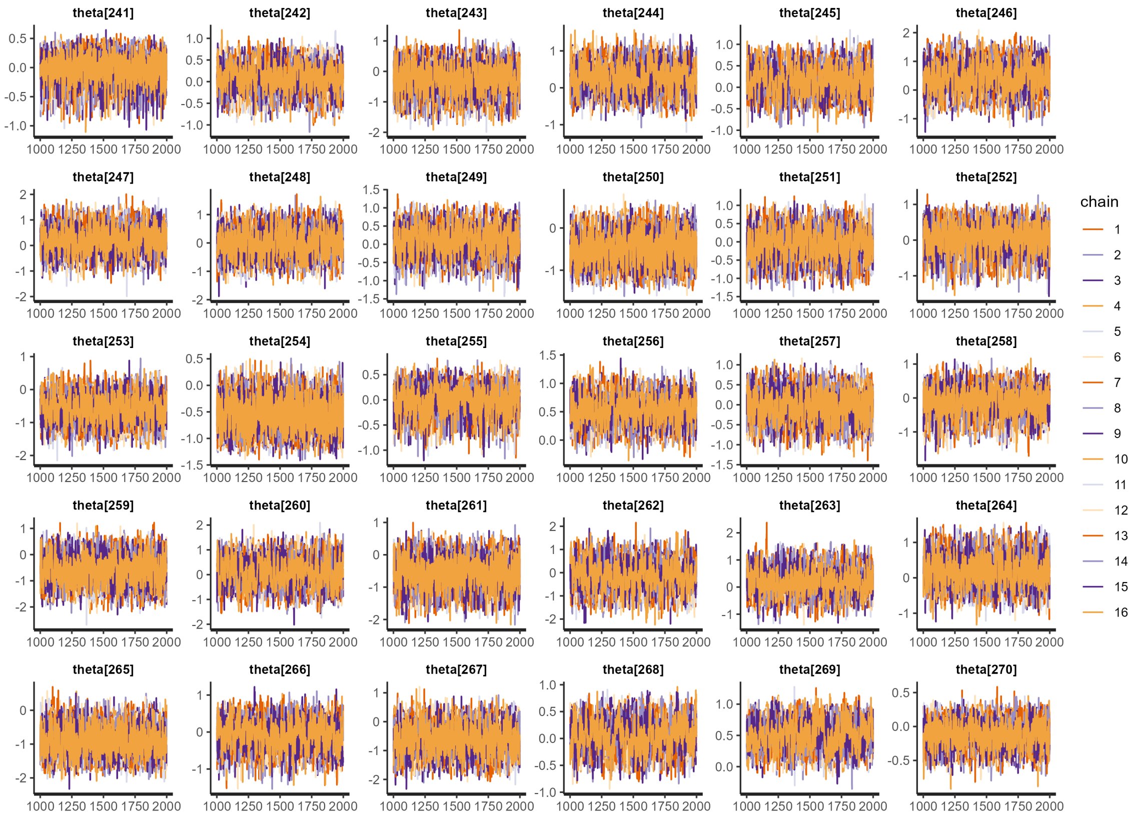

**Figure S57:** Traceplots for the post-burn-in iterations sampling the posterior distributions of $\theta_{241}$ to $\theta_{270}$.

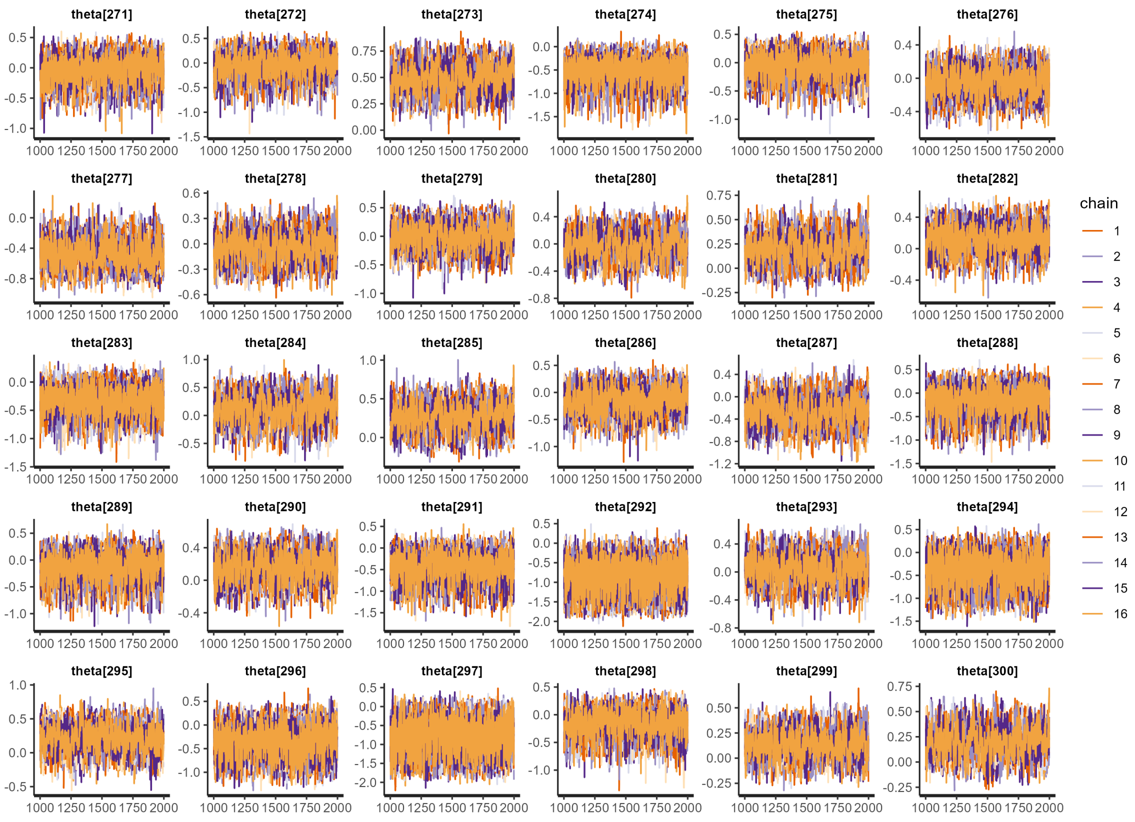

**Figure S58:** Traceplots for the post-burn-in iterations sampling the posterior distributions of $\theta_{271}$ to $\theta_{300}$.

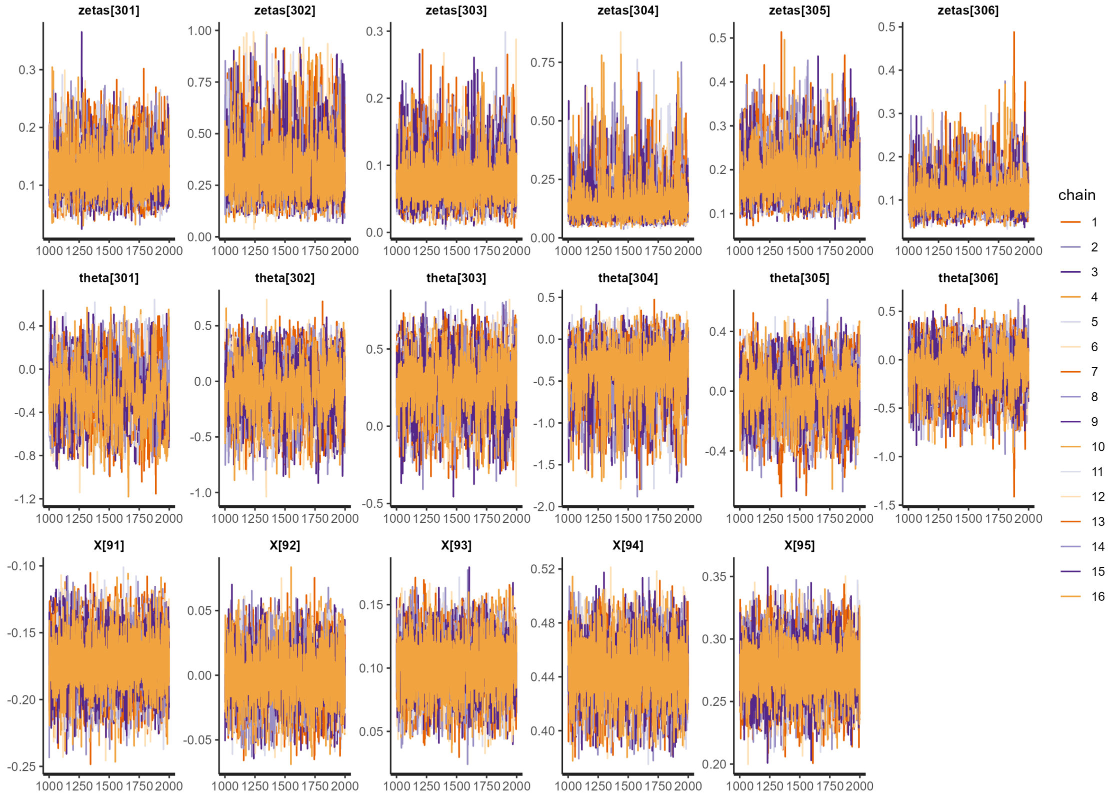

**Figure S59:** Traceplots for the post-burn-in iterations sampling the posterior distributions of $\zeta_{301}$ to $\zeta_{306}$, $\theta_{301}$ to $\theta_{306}$, and $X_{91}$ to $X_{95}$.

### 4 Sensitivity Analyses

To assess the impact that modelling terms have on improving model fit, we consider a total of 12 model formulations, including the main analysis in Equation (3). The 12 models are variations in how we define the number of cases causing onward infection in an LTLA, and how we define the reproduction number.

#### 4.1 Alternate models

We consider models where the sixteen model population / variant / funding variables, $x_{i,t}$ are included, and we consider models where they are removed. We consider models where the spatial error term, $\theta_i$, is included as a model variable, and we consider models where $\theta_i$ is not included. We consider models where the spatial kernel variable, $\zeta_i$ is included for every LTLA. We also consider models where $\zeta$ does not vary by LTLA, and is instead a single fixed model parameter. We also consider models where inter-LTLA infection is not possible, i.e. $\zeta$ is fixed to 0.

These 2×2×3 modelling permutations make up the 12 models considered, which we formally define in Table S3 below.

**Table S3:** Each model considered is a negative binomial model as written in Equation (1). The models differ in the expression of the mean $\mu_{i,t}$, as formally defined in this table.

| Model | Negative Binomial mean $\mu_{i,t}$ |
|---|---|
| **(A)** Main analysis | $\lambda S_{i,t-1}\left(Y_{i,t-1}+\zeta_i\sum_{j\in\Omega_i}Y_{j,t-1}\right)\exp\left(x_{i,t-1}\beta+z_{t-1}+\theta_i\right)$ |
| **(B)** LTLA-varying $\zeta_i$, model variables $x_{i,t}$ removed, $\theta_i$ included. | $\lambda S_{i,t-1}\left(Y_{i,t-1}+\zeta_i\sum_{j\in\Omega_i}Y_{j,t-1}\right)\exp\left(z_{t-1}+\theta_i\right)$ |
| **(C)** LTLA-varying $\zeta_i$, model variables $x_{i,t}$ included, $\theta_i$ removed. | $\lambda S_{i,t-1}\left(Y_{i,t-1}+\zeta_i\sum_{j\in\Omega_i}Y_{j,t-1}\right)\exp\left(x_{i,t-1}\beta+z_{t-1}\right)$ |
| **(D)** LTLA-varying $\zeta_i$, model variables $x_{i,t}$ removed, $\theta_i$ removed. | $\lambda S_{i,t-1}\left(Y_{i,t-1}+\zeta_i\sum_{j\in\Omega_i}Y_{j,t-1}\right)\exp\left(z_{t-1}\right)$ |
| **(E)** constant $\zeta$, model variables $x_{i,t}$ included, $\theta_i$ included. | $\lambda S_{i,t-1}\left(Y_{i,t-1}+\zeta\sum_{j\in\Omega_i}Y_{j,t-1}\right)\exp\left(x_{i,t-1}\beta+z_{t-1}+\theta_i\right)$ |
| **(F)** constant $\zeta$, model variables $x_{i,t}$ removed, $\theta_i$ included. | $\lambda S_{i,t-1}\left(Y_{i,t-1}+\zeta\sum_{j\in\Omega_i}Y_{j,t-1}\right)\exp\left(z_{t-1}+\theta_i\right)$ |
| **(G)** constant $\zeta$, model variables $x_{i,t}$ included, $\theta_i$ removed. | $\lambda S_{i,t-1}\left(Y_{i,t-1}+\zeta\sum_{j\in\Omega_i}Y_{j,t-1}\right)\exp\left(x_{i,t-1}\beta+z_{t-1}\right)$ |
| **(H)** constant $\zeta$, model variables $x_{i,t}$ removed, $\theta_i$ removed. | $\lambda S_{i,t-1}\left(Y_{i,t-1}+\zeta\sum_{j\in\Omega_i}Y_{j,t-1}\right)\exp\left(z_{t-1}\right)$ |
| **(I)** $\zeta_i$ removed, model variables $x_{i,t}$ included, $\theta_i$ included. | $\lambda S_{i,t-1}\left(Y_{i,t-1}\right)\exp\left(x_{i,t-1}\beta+z_{t-1}+\theta_i\right)$ |
| **(J)** $\zeta_i$ removed, model variables $x_{i,t}$ removed, $\theta_i$ included. | $\lambda S_{i,t-1}\left(Y_{i,t-1}\right)\exp\left(z_{t-1}+\theta_i\right)$ |
| **(K)** $\zeta_i$ removed, model variables $x_{i,t}$ included, $\theta_i$ removed. | $\lambda S_{i,t-1}\left(Y_{i,t-1}\right)\exp\left(x_{i,t-1}\beta+z_{t-1}\right)$ |
| **(L)** $\zeta_i$ removed, model variables $x_{i,t}$ removed, $\theta_i$ removed. | $\lambda S_{i,t-1}\left(Y_{i,t-1}\right)\exp\left(z_{t-1}\right)$ |

For simplicity, only three aditional models were presented in the main manuscript, alongside the primary analysis (model

(A)). The manuscript presents model comparison results for models (A), (B), (I) and (J).

#### 4.2 Model comparison

To compare models, we need a metric by which to assess how well the model explains the given data. The estimated log pointwise predictive density (elpd) [22], is one such measure. However, the metric is susceptible to being skewed by highly significant data. Ideally, we want a model that remains an effective simulation of the data when data points are removed. Leave-one-out cross validation (LOO-CV) is a way to correct for this. If there are $N$ data being fit to, $\text{elpd}_{\text{loo}}$ is defined as the sum of $N$ elpd calculations, each time with one data point left out of the data set being fit to. However, such a procedure is unsuitable for spatial and time-series data, due to the inherent lack of independence between data points. As such, we construct a model comparison metric more representative of the predictive task being assessed. For each of the twelve models (A to L), we fit the model 306 times (once for each LTLA), each time leaving out the second-half of the time series data (weeks 47 to 95) for one LTLA in particular. In each instance, we then only calculate the log-likelihood (elpd) for the data left out during the fitting process. Thus, we are assessing how well the model can predict the future time series for that LTLA based on our knowledge of covariates of interest and observed spatial importation effects (our intended result). We then sum these 306 elpd scores together to arrive at a cumulative final elpd (LFO) score for each of the twelve models.

A greater elpd (LFO) value indicates a better model fit.

We calculate the elpd (LFO) score for each model, and present the point estimate and the standard error for each in Table S4 and figure S60 below.

We note that greater mean estimate scores are sometimes calculated for, seemingly, more restrictive models, such as model C compared to model A. We highlight that in due to the leave-future-out scheme undertaken, this is reflective of the more flexible model "over-fitting" its spatial random effect terms ($\theta_i$) to the first half of the time-series included. As such, the more restrictive model performs better, due to not being overly tuned to the earlier time-series.

**Table S4:** Estimated elpd (LFO) estimates and standard error for models.

| Model | elpd (LFO) point estimate (standard error) |
|---|---|
| **(A)** Main analysis | -81,006 (505) |
| **(B)** LTLA-varying $\zeta_i$, model variables $x_{i,t}$ removed, $\theta_i$ included. | -81,124 (502) |
| **(C)** LTLA-varying $\zeta_i$, model variables $x_{i,t}$ included, $\theta_i$ removed. | -80,487 (506) |
| **(D)** LTLA-varying $\zeta_i$, model variables $x_{i,t}$ removed, $\theta_i$ removed. | -80,659 (513) |
| **(E)** constant $\zeta$, model variables $x_{i,t}$ included, $\theta_i$ included. | -80,589 (492) |
| **(F)** constant $\zeta$, model variables $x_{i,t}$ removed, $\theta_i$ included. | -80,762 (491) |
| **(G)** constant $\zeta$, model variables $x_{i,t}$ included, $\theta_i$ removed. | -81,027 (482) |
| **(H)** constant $\zeta$, model variables $x_{i,t}$ removed, $\theta_i$ removed. | -81,247 (472) |
| **(I)** $\zeta_i$ removed, model variables $x_{i,t}$ included, $\theta_i$ included. | -83,945 (667) |
| **(J)** $\zeta_i$ removed, model variables $x_{i,t}$ removed, $\theta_i$ included. | -84,018 (666) |
| **(K)** $\zeta_i$ removed, model variables $x_{i,t}$ included, $\theta_i$ removed. | -83,641 (665) |
| **(L)** $\zeta_i$ removed, model variables $x_{i,t}$ removed, $\theta_i$ removed. | -83,749 (666) |

We note that, if the data were independently sampled, the "leave-one-out-information criterion" (LOOIC) is a more traditional measure of model comparison, which we also present for completeness in Section 4.9.

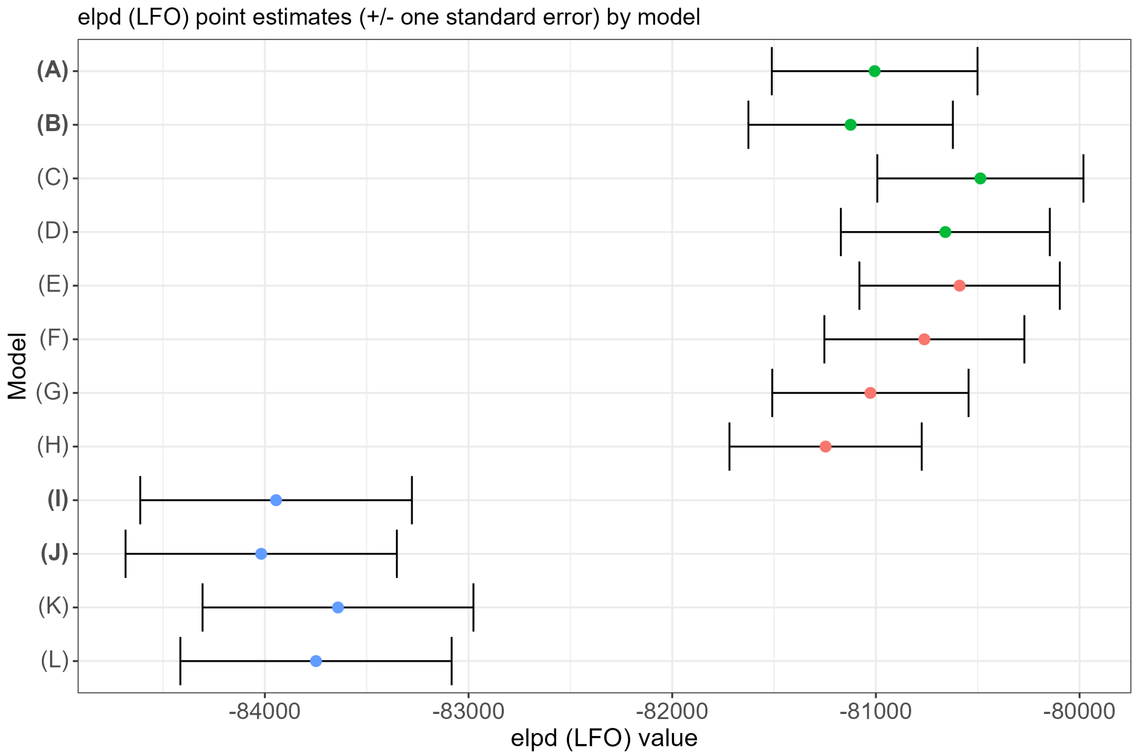

**Figure S60:** The estimated log pointwise predictive density (elpd) leave-future-out (LFO) score point estimates and standard error range for the 12 models defined in Table S3. Models (A), (B), (I) and (J) in bold are the four models presented in the main manuscript.

#### 4.3 Univariate model comparison

Due to the inherent correlation between model variables as shown in Section 1.7, it is useful to fit model (A) (as defined in Table S3) with just one of the 16 variables in $x_{i,t}$, and to compare the posterior estimate for $\beta$ between the univariate and multivariate models.

Figure S61 shows that there is very little difference between the two approaches, and the credible intervals overlap in almost all instances. This indicates that the correlation between variables is having minimal impact on the interpretation of the relative importance of each variable.

One difference to note though is that the credible interval for variable 4, the IMD average score, no longer straddles zero for the univariate model, suggesting that the variable is significantly impactful when one corrects for the correlation with other variables. For example, Figure S31 suggests strong correlation between IMD and the COMF funding, as more deprived areas will have been assigned greater funding resources.

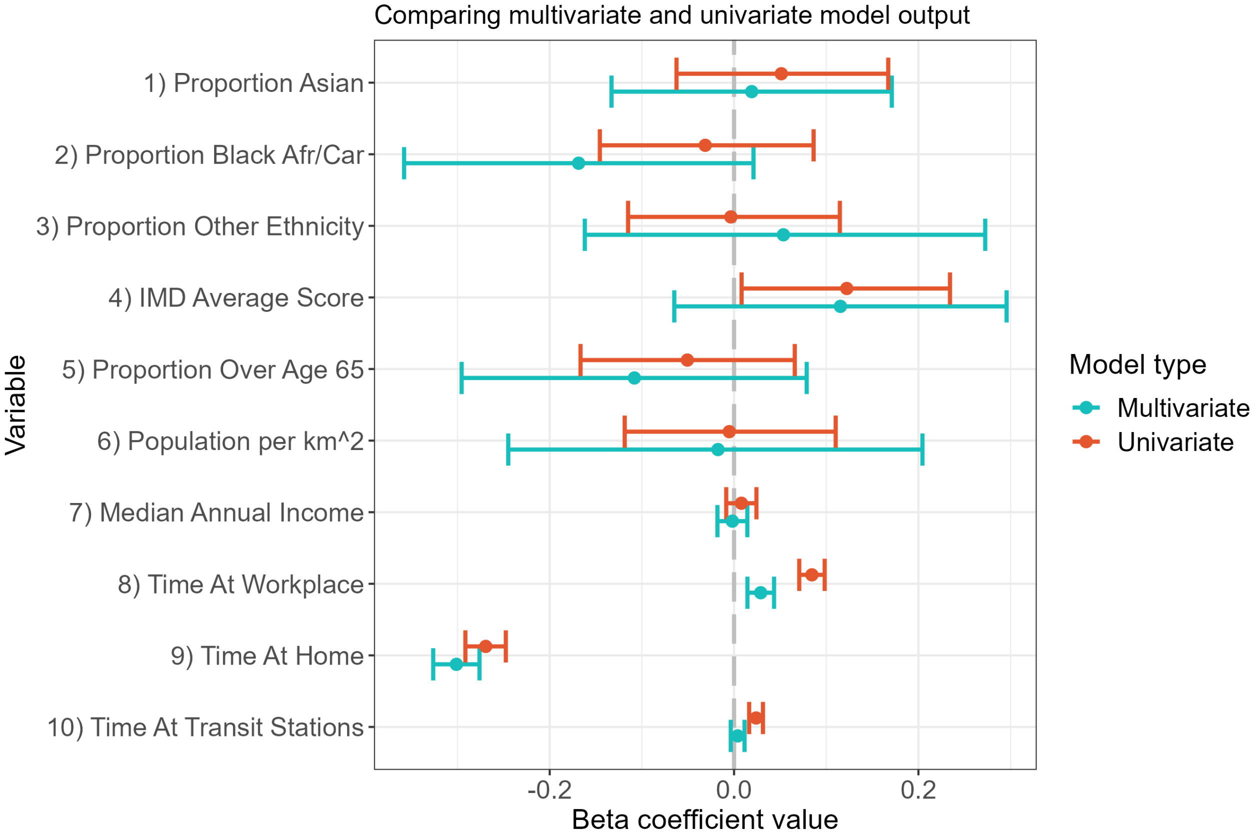

**Figure S61:** The posterior estimates of $\beta$ are shown for the multivariate model including all variables, and the univariate model featuring only one of the variables. The multivariate model (in blue) is identical to the distributions shown in Figure 3 of the main manuscript.

We also note that the impact of Black population proportion is greatly reduced in the univariate model.

We do not plot COVID-19 variant univariate results, as these variables rely on being fit alongside all variants for proper interpretation. Nor do we plot funding univariate results, as the significant drop in funding between financial years means that their impact can only feasibly be assessed with a model that includes COVID-19 variant proportion as well.

We also seek to understand how these correlation effects change in response to the absence of spatial random effects ($\theta_i$), and so recreate Figure S61 for models C, G, and K as introduced above in Table S3 - models with our covariates of interest, but with different spatial importation assumptions, and spatial random effects removed. These are presented in Figure S62 and **????**. In general, we see that this causes the width of the confidence intervals to shrink, also demonstrating a greater impact of correlation between variables, though these values remain small in magnitude.

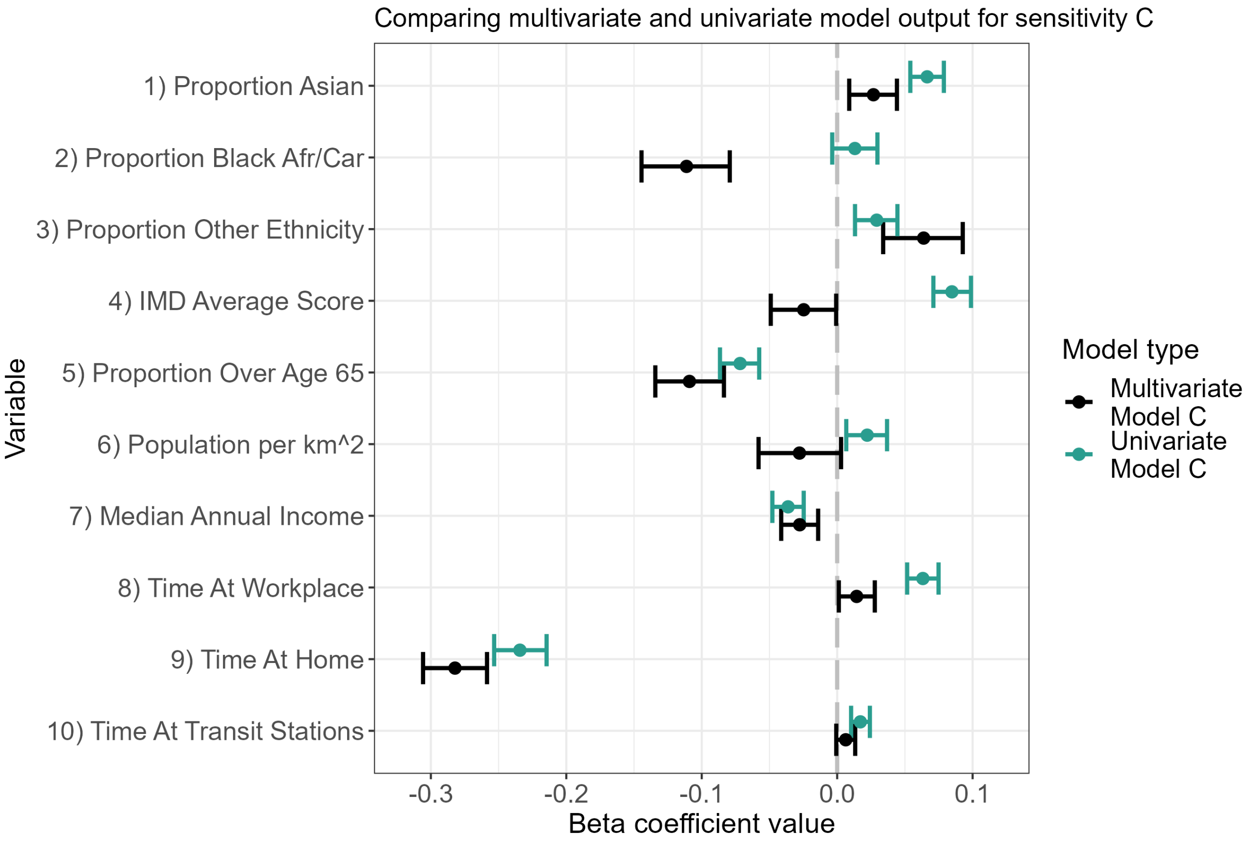

**Figure S62:** The posterior estimates of $\beta$ are shown for the multivariate version of model C including all variables, and the univariate version of model C featuring only one of the variables. Points indicate the mean posterior estimate, and bars indicate the 95% CIs.

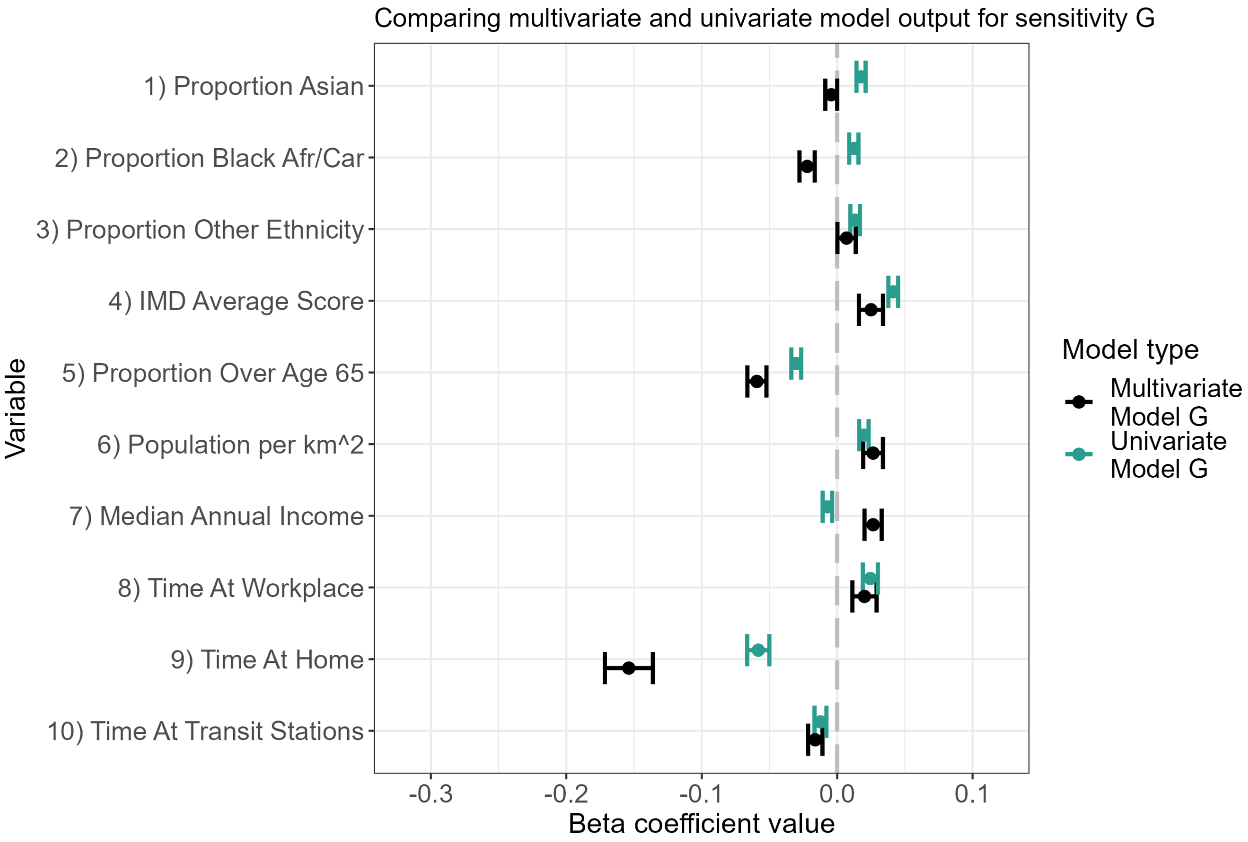

**Figure S63:** The posterior estimates of $\beta$ are shown for the multivariate version of model G including all variables, and the univariate version of model G featuring only one of the variables. Points indicate the mean posterior estimate, and bars indicate the 95% CIs.

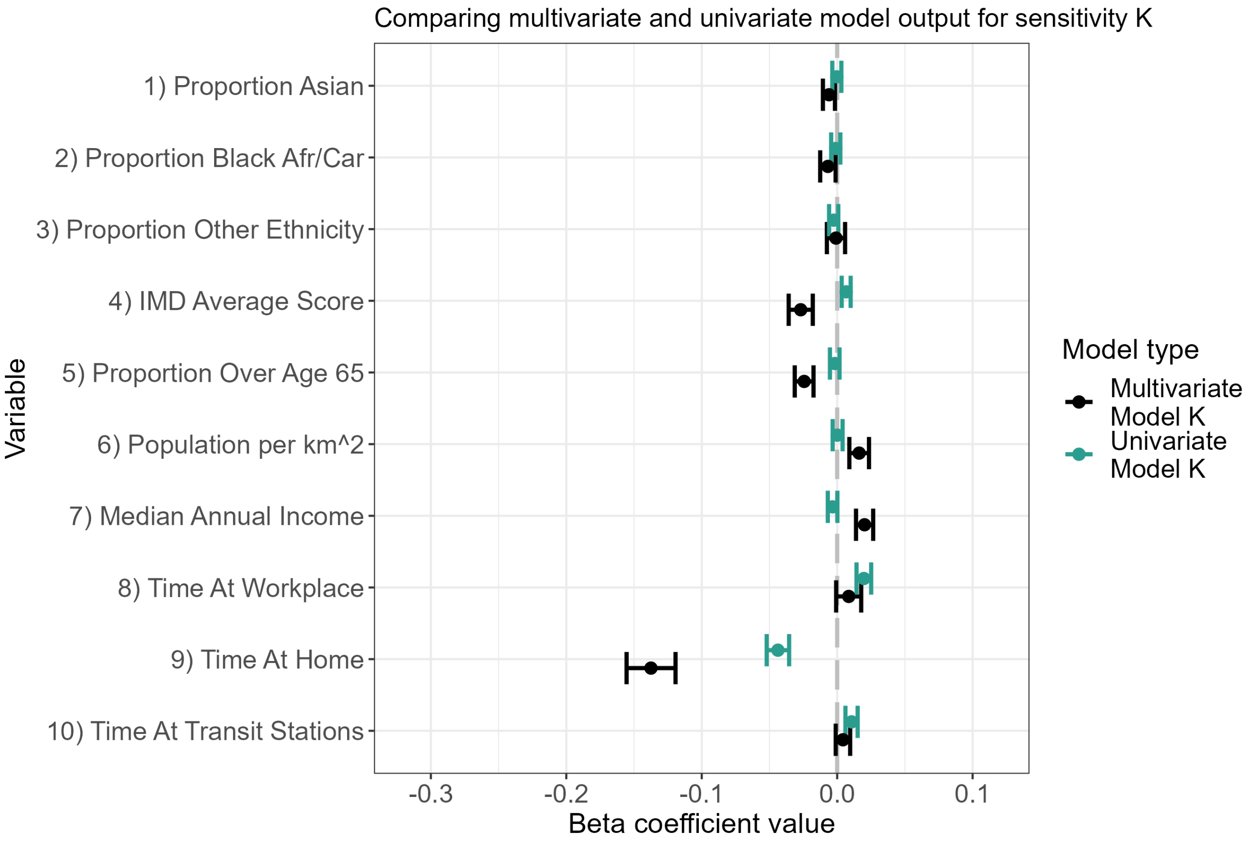

**Figure S64:** The posterior estimates of $\beta$ are shown for the multivariate version of model K including all variables, and the univariate version of model K featuring only one of the variables. Points indicate the mean posterior estimate, and bars indicate the 95% CIs.

#### 4.4 Alternate data streams

We further test the reliability of our model by repeating the analysis for alternate data streams.

Firstly, as explained in Section 1.2, we consider only pillar 2 cases confirmed by PCR testing. As an additional sensitivity analysis, we also fit the model to the broader definition featured on the UK COVID-19 data dashboard [2], which encompasses both pillar 1 and pillar 2 cases, and also includes cases that were confirmed only by LFDs or loop-mediated isothermal amplification (LAMP) test. If a case tested positive via LFD but then tested negative by PCR, they are not counted as a case. Figure S65 shows the national totals for case numbers via these two definitions. We see that more cases are reported from the dashboard as time goes on - in line with the increasing use of LFDs.

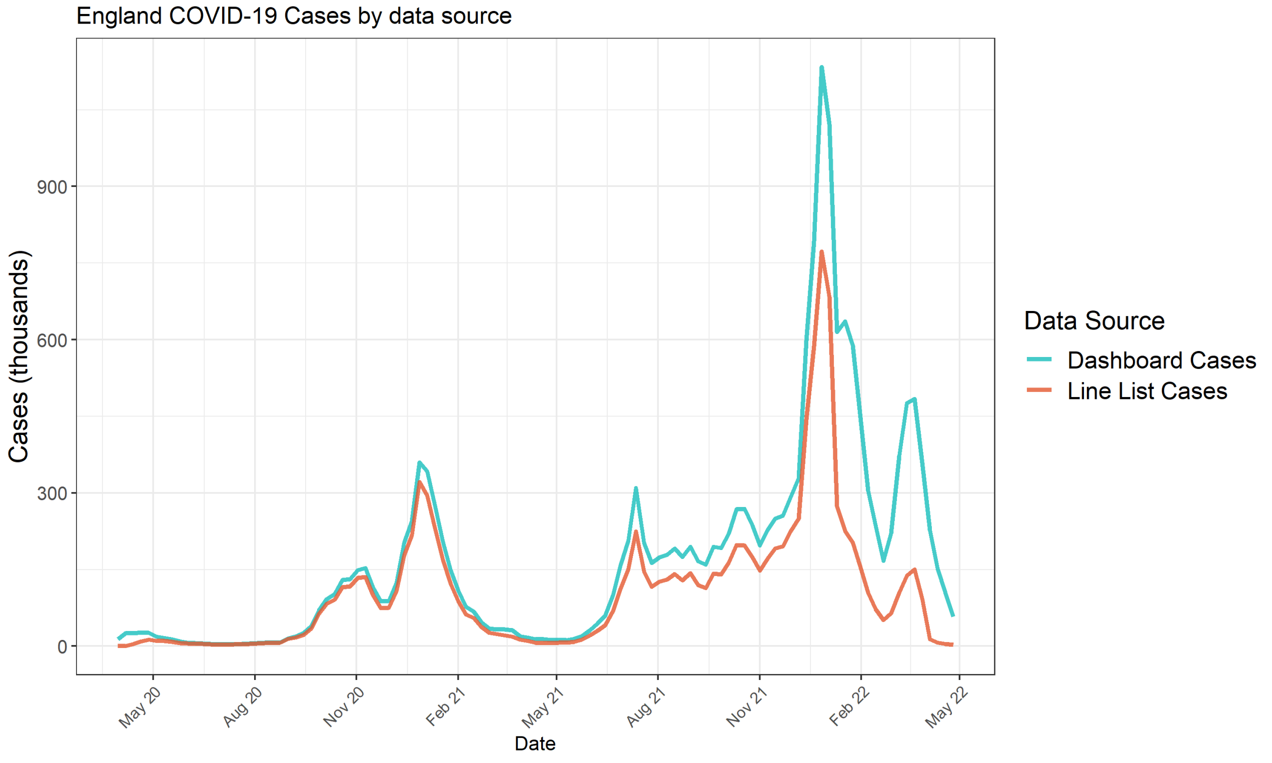

**Figure S65:** The number of new registered cases each week by case definition. Red depicts the definition used in the main manuscript and previous analyses - only pillar 2, PCR-confirmed cases from the national linelist. Blue depicts the number of cases as reported on the UK dashboard [2] - pillar 1 and pillar 2, confirmed via PCR, LFD, or LAMP.

Additionally, we considered an alternate definition of variant-of-concern (VOC) proportions. As defined in Section 1.3, we use SGTF data from the national line list to report the proportion of cases that belong to specific VOCs each week. We consider a sensitivity analysis where we use variant data from the UK dashboard. This VOC identity dashboard data is confirmed via whole genome sequencing (WGS), however the data is only available as fine as the NHS region level: East of England, London, Midlands, North East and Yorkshire, North West, South East, South West. We assume that every LTLA in an NHS region has the same VOC proportions over time.

This data stream reports sub-lineages, which we aggregate. "V-21APR-02 (Delta B.1.617.2)" and "V-21OCT-01 (Delta AY 4.2)" are combined to just "Delta". "VOC-21NOV-01 (Omicron BA.1)", "VOC-22JAN-01 (Omicron BA.2)", "VOC-22APR-03 (Omicron BA.4)", "VOC-22APR-04 (Omicron BA.5)", "V-22OCT-01 (Omicron BQ.1)" are combined to just "Omicron".

Figure S66 displays how the variable coefficients, $\beta$, varies for these different data streams. There is no noticeable difference in the relative importance of any of the variables except for variant proportion. When using the WGS dashboard variant data, variants are considered to be more impactful in driving further transmission.

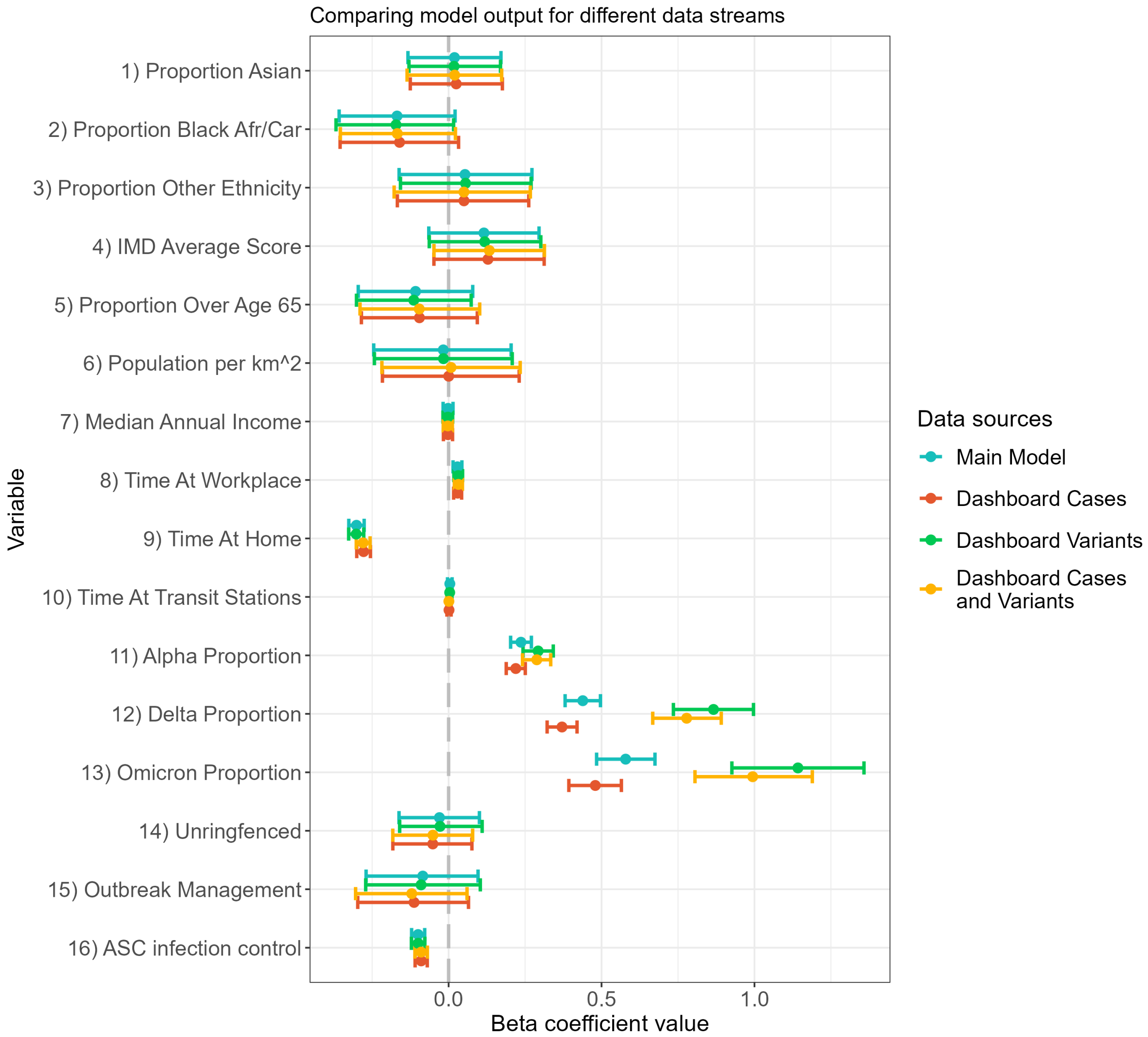

**Figure S66:** We plot the $\beta$ coefficient values for four sensitivity analyses. Blue shows the main analysis as presented in the main manuscript. Red shows the output when fit to case data from the UK dashboard [2]. Green shows when variant proportion is instead defined by whole genome sequencing across NHS regions, from the UK dashboard. Yellow shows when both case definition and variant definition is taken from the UK dashboard.

#### 4.5 Fitting to subsections of the time series

The vector, $\beta$, representing the contribution coefficients of our covariates is static in time. To explore whether these covariates differ in time, we re-fit the baseline analysis (model A) to three distinct subsections of the whole time series;

1. May 10th 2020 - January 3rd 2021,
2. January 10th 2021 - August 1st 2021,
3. August 8th 2021 - February 27th 2022.

These dates broadly correspond to, "the pre-vaccination period", "the final lockdown" period, and the "post-lockdown" period, respectively. Figure S67 below shows the resulting beta coefficients for these three sensitivites. We see that the results are broadly unchanged across all sensitivities. We note that the random walk term follows the same general trend, although will be re-scaled in each instance for the random walk to begin from it's initial prior step of a standard normal distribution. Since the third period (green) relates to the period when no stay-at-home orders were in place, we can see that this scaling difference in the random walk is directly attributed to a reduction in the importance of the residential mobility term.

The one difference of note is that we can see a time-sensitive impact of the importance of the ASC infection control fund. The respective covariate's importance is principly seen during the second time period, after vaccination was introduced, and capturing the remainder of England's stay-at-home order. This aligns with what was observed at the time, whereby increased transmission in care homes in England was prominent at the start of the COVID-19 pandemic [23], necessitating greater focus on adult social care provision leading into the second period considered. This effect then diminishes as non-pharmaceutical interventions were broadly ended in the third period considered.

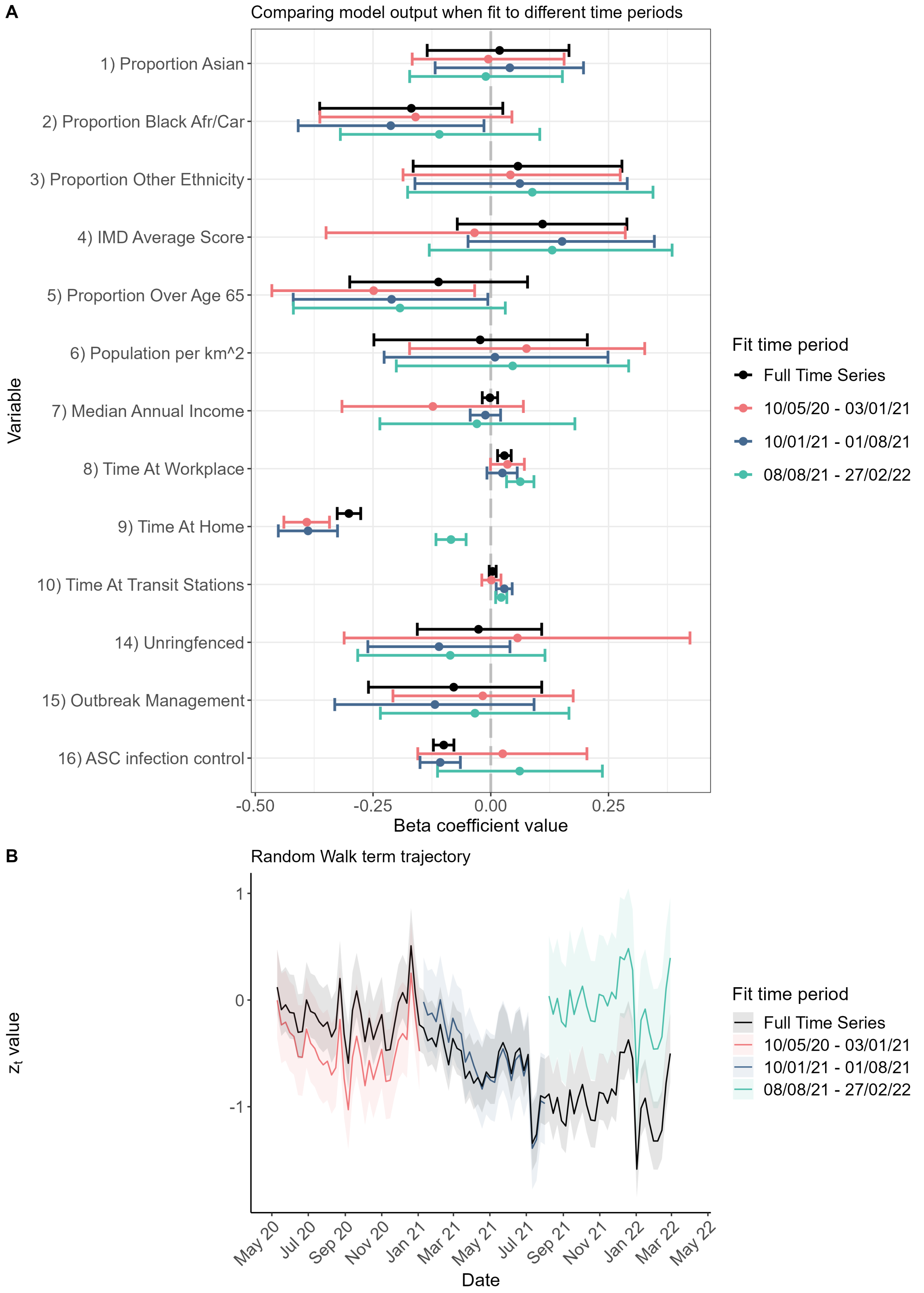

**Figure S67:** We plot the $\beta$ coefficient values **(A)** and $z_t$ random walk values **(B)** when the baseline analysis is repeated for three distinct subsets of the time series. Black shows the main analysis as presented in the main manuscript. Red shows when the model is fit to just May 10th 2020 - January 3rd 2021. Blue shows when the model is fit to just January 10th 2021 - August 1st 2021. Green shows when the model is fit to just August 8th 2021 - February 27th 2022. The line in plot (B) depicts the mean estimate, and the shaded region the 95% CI.

#### 4.6 Infection-acquired immunity

To adjust for the impact of infection-acquired immunity, our model incorporates the reports of "first-episode" infections through the $S_{i,t}$ term introduced in Equation (3). This formulation assumes permanent protection in its current formulation, an assumption we test with two additional sensitivity analyses in this section.

The UK SIREN study assessed ongoing protection against reinfection from primary infection via routine testing of a cohort of health service personnel in England throughout the pandemic. Protection against reinfection was found to be very high for the wild-type and Alpha variant [24], remaining strong for the Delta variant, though beginning to wane for the Omicron variant [25]. Similar results are found in the global systematic review conducted by Kojima et al. (2021) [26], which reports an average risk reduction against reinfection of 90.4% during this period, and where protection against reinfection holds for, on average, up to 10 months.

We conduct two sensitivity analyses to account for the potential impact of reinfection. Firstly, a sensitivity analysis where we only fit the model up to September 12th 2021 – the last week before any confirmed Omicron cases are reported. Secondly, we adjust the model such that reported first episodes are only factored into the term $S_{i,t}$ for the first 10 months after they are reported, effectively assuming infected individuals are immune to reinfection for 10 months only following infection.

The results of these sensitivities are presented below in Figure S68. Our results remain unchanged in both of these analyses.

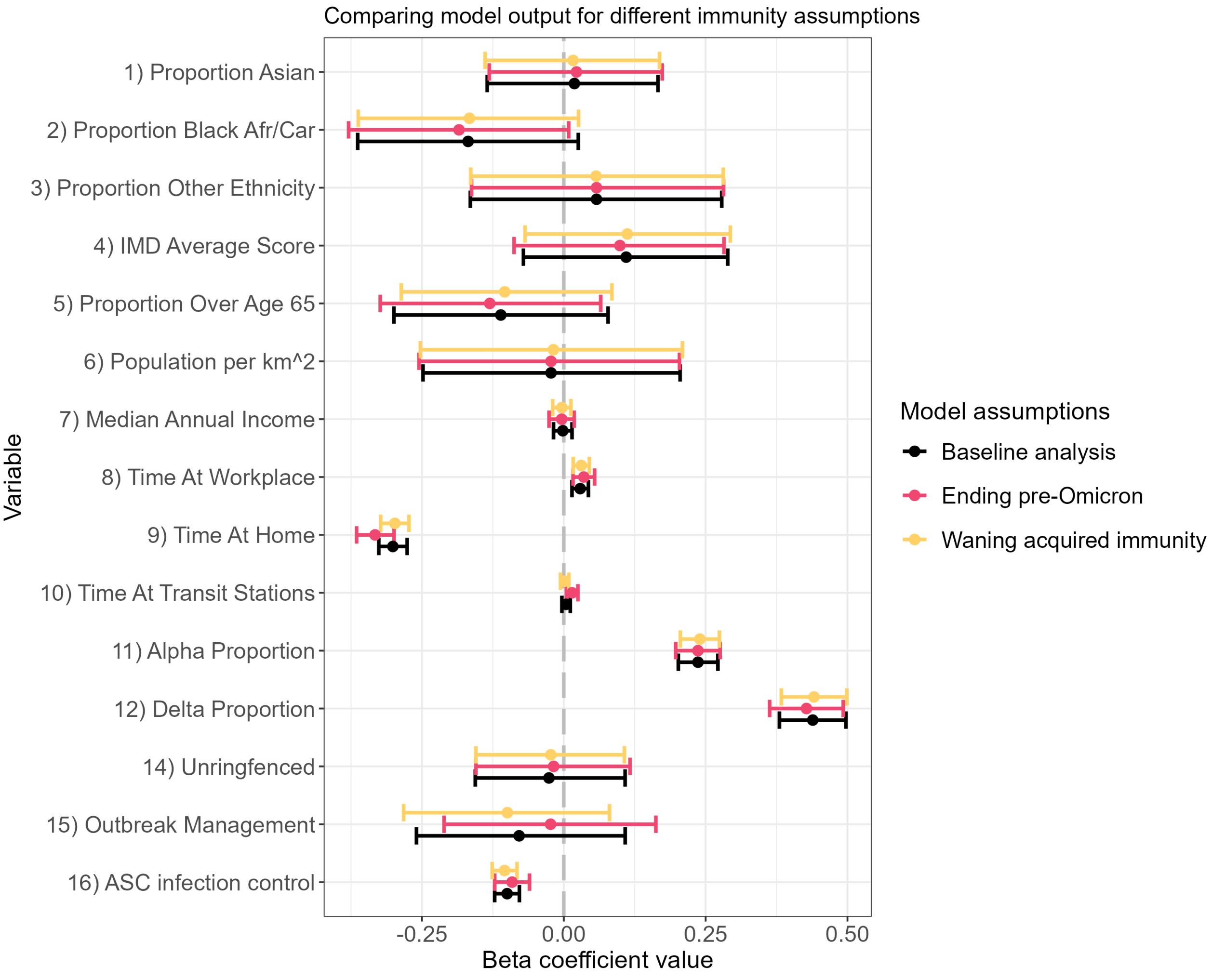

**Figure S68:** We plot the $\beta$ coefficient values for sensitivity analyses considering, (i) when the model is fit up to the emergence of the Omicron variant (September 12th 2021), and (ii) when a recorded first-episode is only assumed to offer protection for 10 months. Black shows the main analysis as presented in the main manuscript. Pink shows sensitivity (i). Yellow shows sensitivity (ii).

#### 4.7 Variation in reporting by covariate

There has been some indication that reporting of cases may have varied by ethnicity during the pandemic. Mathur et al. (2021) [27] showed that while ethnicity did not impact reported testing during the first wave of the pandemic, a slight trend of reduced reporting for Black and Asian populations could be identified for the second wave.

Our model assumes equal reporting by covariate, and to explore this assumption we conduct three additional sensitivity analyses testing this assumption. The UK linelist case data that we fit to provides detail on ethnicity and IMD decile of each case. We conduct three sensitivity analyses where we double the number of cases reported as (i) Black Afr/Car, (ii) Asian, (iii) top two IMD deciles (the two most deprived), respectively.

Figure S69 shows the $\beta$ coefficient values for these three sensitivities compared to our baseline study, demonstrating that our results are unaffected by these changes.

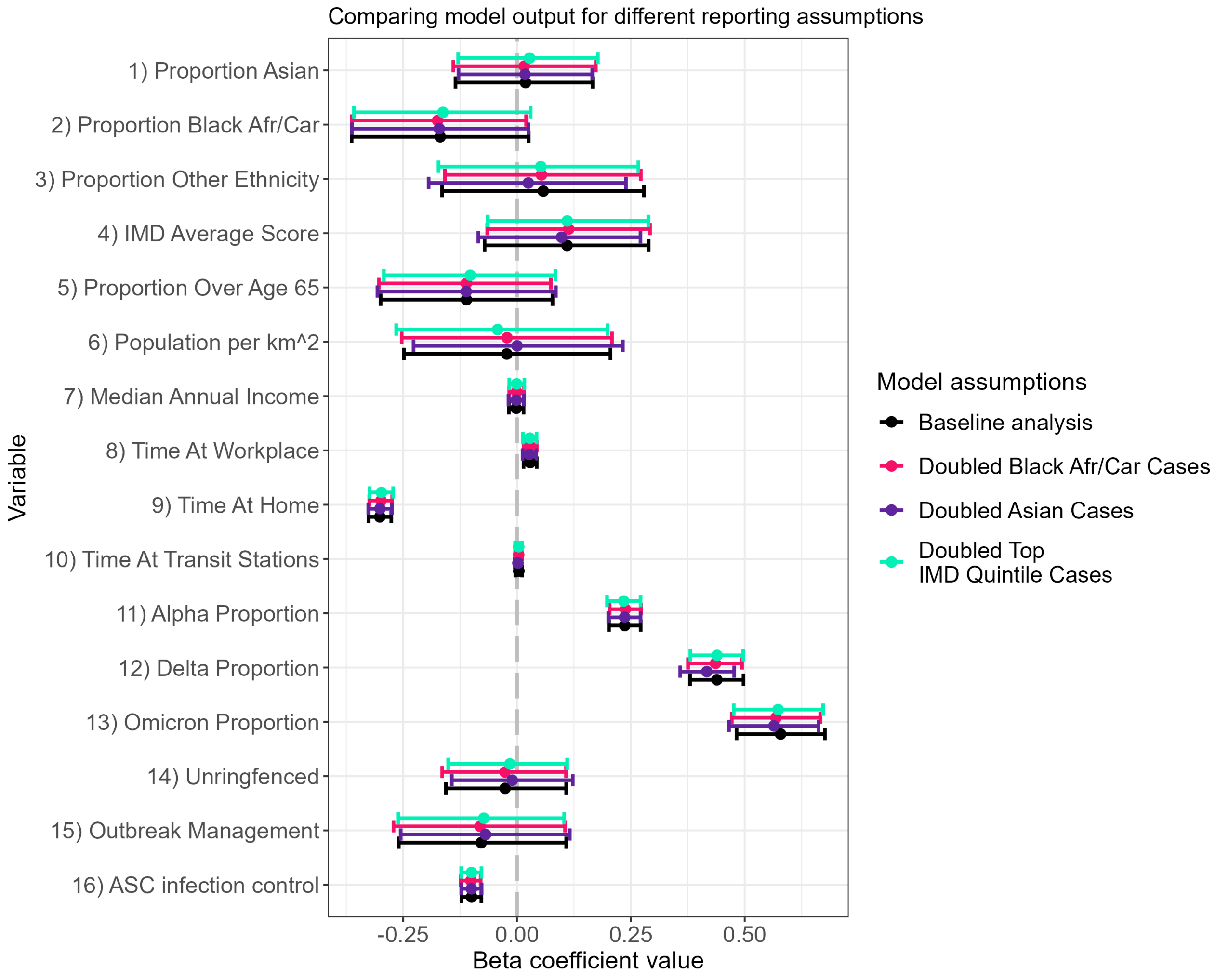

**Figure S69:** We plot the $\beta$ coefficient values for sensitivity analyses exploring differences in reporting by ethnicity and IMD. We fit three different sensitivity analyses: (i) where the number of Black Afr/Car reported cases is doubled (ii) where the number of Asian reported cases is doubled (iii) where the the number of top two (IMD) decile reported cases is doubled. Black shows the main analysis as presented in the main manuscript. Pink shows sensitivity (i). Purple shows sensitivity (ii). Green shows sensitivity (iii).

#### 4.8 Gravity model spatial kernel

Our model considers the impact of spatial exportation of cases through an adjacency assumption, whereby cases in an LTLA can cause secondary cases in either their original LTLA, or LTLAs they share a direct border with. As an extension of this, we also consider an alternate model of spatial exportation based on the gravity model assumptions of Truscott & Ferguson (2012) [28] as a sensitivity analysis.

Our initial model considers attacking cases of the form $\left(Y_{i,t-1} + \zeta_i \sum_{j \in \Omega_i} Y_{j,t-1}\right)$, where $\zeta_i$ is a model parameter controlling what proportion of cases in neighbouring LTLAs trigger secondary cases within LTLA $i$. Instead, we consider the following format, where $\left(Y_{i,t-1} + \zeta_i \sum_{j \in \Omega_i} Y_{j,t-1}\right)$ is replaced by

$$(GY_{t-1}) \tag{4}$$

where $Y_{t-1}$ is the column vector of all cases in each LTLA at week $t-1$, and $G$ is a matrix where element $\{i, j\}$ details the probability of a case in LTLA $j$ causing a secondary infection in LTLA $i$ subject to gravity equation assumptions detailed as follows. Consider the symmetric matrix $g$, where element $g(i, j)$ is defined as

$$g(i,j) = \frac{\text{Population}(i)\text{Population}(j)}{\left(\frac{\text{Distance}(i,j)}{\alpha}\right)^{\gamma}} \tag{5}$$

where Population$(i)$ is the proportion of the national population residing in LTLA $i$, Distance$(i, j)$ is the euclidean distance between the midpoints of LTLAs $i$ and $j$, and $\alpha$ and $\gamma$ are model parameters controlling the relative importance of distance between locations. Matrix $G$ is then acquired by scaling each column of matrix $g$ to sum to 1.

Figure S70 below depicts the mean and 95% CI of the posterior distributions for the gravity parameters $\gamma$ and $\alpha$, 1.41 (1.35 - 1.47) and 0.0019 (0.0016 - 0.0023) respectively. Figure S71 shows the $\beta$ coefficients for the baseline analysis and the gravity model sensitivity analysis. Figure S71 shows that our study results are unchanged by this change of spatial kernel, and Figure S70 shows, by the low value of $\alpha$, and $\gamma$ being greater than 1, that this formulation strongly highlights the importance of distance - i.e. more geographically close cases are the greatest contribution of onward infection, further supporting our use of an adjacency model.

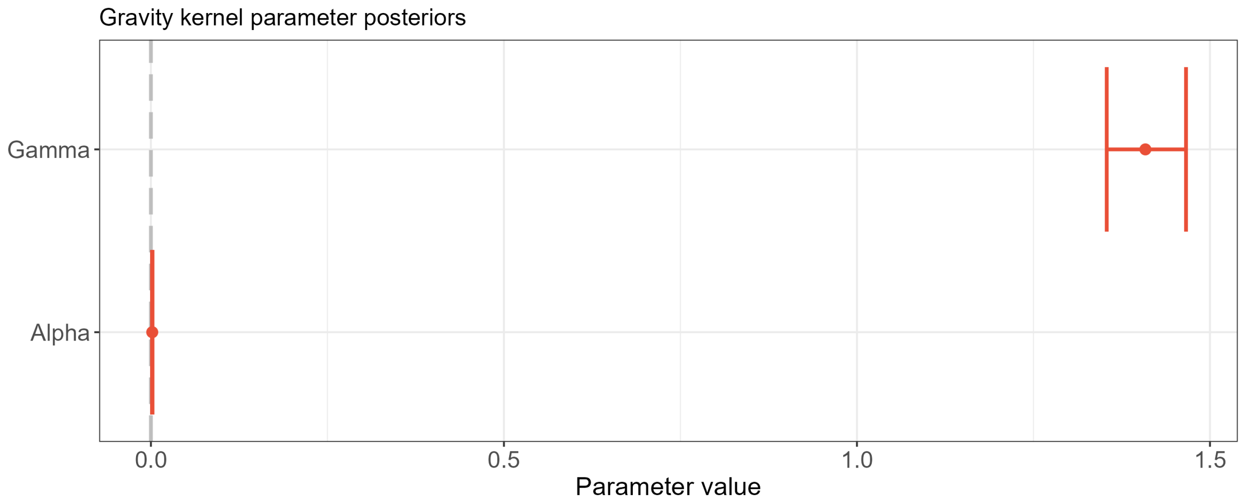

**Figure S70:** We plot the mean and 95% CI of the posterior distribution for gravity model parameters $\alpha$ and $\gamma$.

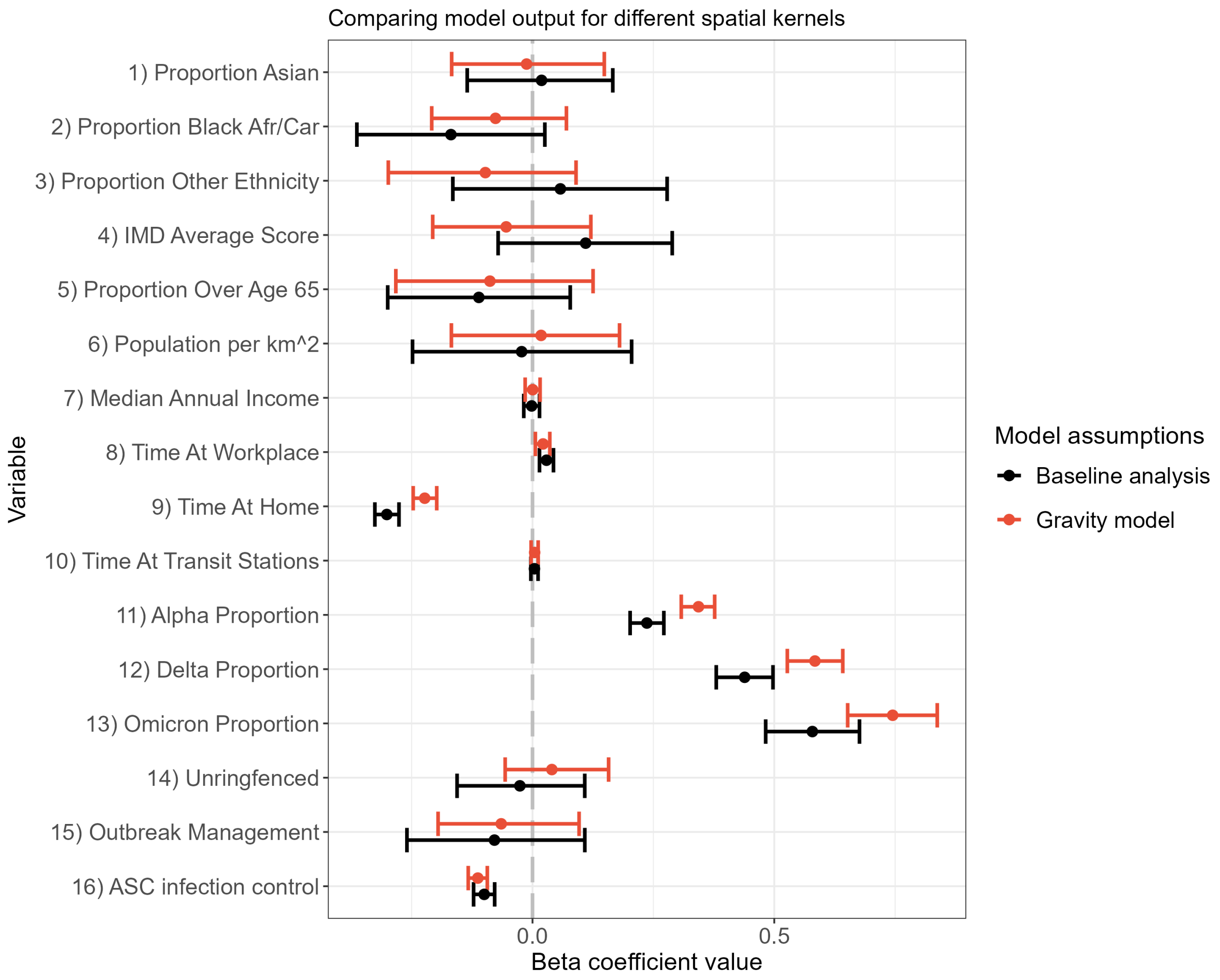

**Figure S71:** We plot the $\beta$ coefficient values for our baseline analysis, and a sensitivity analysis where the spatial importation of cases is described by a gravity model. Black depicts the baseline anaylsis and red the gravity model sensitivity.

#### 4.9 Model comparison via LOOIC

In Section 4.2 we introduce our measure of model comparison. Usually, Leave-one-out cross validation (LOO-CV) is a standard way to calculate elpd estimates. We did not take this approach due to the lack of independence in time series data. For completeness, in this section, we nonetheless present model comparison scores via LOO-CV, though we caution interpretation, as discussed in greater detail in Vehtari et al. (2018) [29].

Usually, this would require re-fitting the model $N$ times, an unfeasibly long time for models (and data) of our size. However, $\text{ELPD}_{\text{loo}}$ can instead be approximated using Pareto smoothed importance sampling (PSIS), via importance weighted moment matching, as outlined in Vehtari et al. (2015) [30]. Finally, one transforms this value into the LOO information criterion (LOOIC) via the transform $\text{LOOIC} = -2\ \text{ELPD}_{\text{loo}}$, to provide the output on the more-commonly presented scale of "deviance".

A smaller LOOIC value indicates a better model fit.

We calculate the LOOIC for each model, using the loo() function in `rstan` and present the point estimate, and one standard deviation confidence interval for each in Table S5 and figure S72 below.

**Table S5:** Estimated LOOIC estimates and one standard deviation range for models, calculated via Pareto smoothed importance sampling (PSIS).

| Model | LOOIC point estimate ($\pm$ one standard deviation) |
|---|---|
| **(A)** Main analysis | 289,620 (288,896 - 290,344) |
| **(B)** LTLA-varying $\zeta_i$, model variables $x_{i,t}$ removed, $\theta_i$ included. | 290,586 (289,869 - 291,303) |
| **(C)** LTLA-varying $\zeta_i$, model variables $x_{i,t}$ included, $\theta_i$ removed. | 289,912 (289,190 - 290,634) |
| **(D)** LTLA-varying $\zeta_i$, model variables $x_{i,t}$ removed, $\theta_i$ removed. | 291,119 (290,403 - 291,835) |
| **(E)** constant $\zeta$, model variables $x_{i,t}$ included, $\theta_i$ included. | 290,756 (290,033 - 291,479) |
| **(F)** constant $\zeta$, model variables $x_{i,t}$ removed, $\theta_i$ included. | 291,739 (291,022 - 292,456) |
| **(G)** constant $\zeta$, model variables $x_{i,t}$ included, $\theta_i$ removed. | 295,371 (294,638 - 296,104) |
| **(H)** constant $\zeta$, model variables $x_{i,t}$ removed, $\theta_i$ removed. | 296,823 (296,089 - 297,557) |
| **(I)** $\zeta_i$ removed, model variables $x_{i,t}$ included, $\theta_i$ included. | 314,906 (313,035 - 316,777) |
| **(J)** $\zeta_i$ removed, model variables $x_{i,t}$ removed, $\theta_i$ included. | 315,832 (313,960 - 317,704) |
| **(K)** $\zeta_i$ removed, model variables $x_{i,t}$ included, $\theta_i$ removed. | 314,766 (312,895 - 316,637) |
| **(L)** $\zeta_i$ removed, model variables $x_{i,t}$ removed, $\theta_i$ removed. | 315,258 (313,385 - 317,131) |

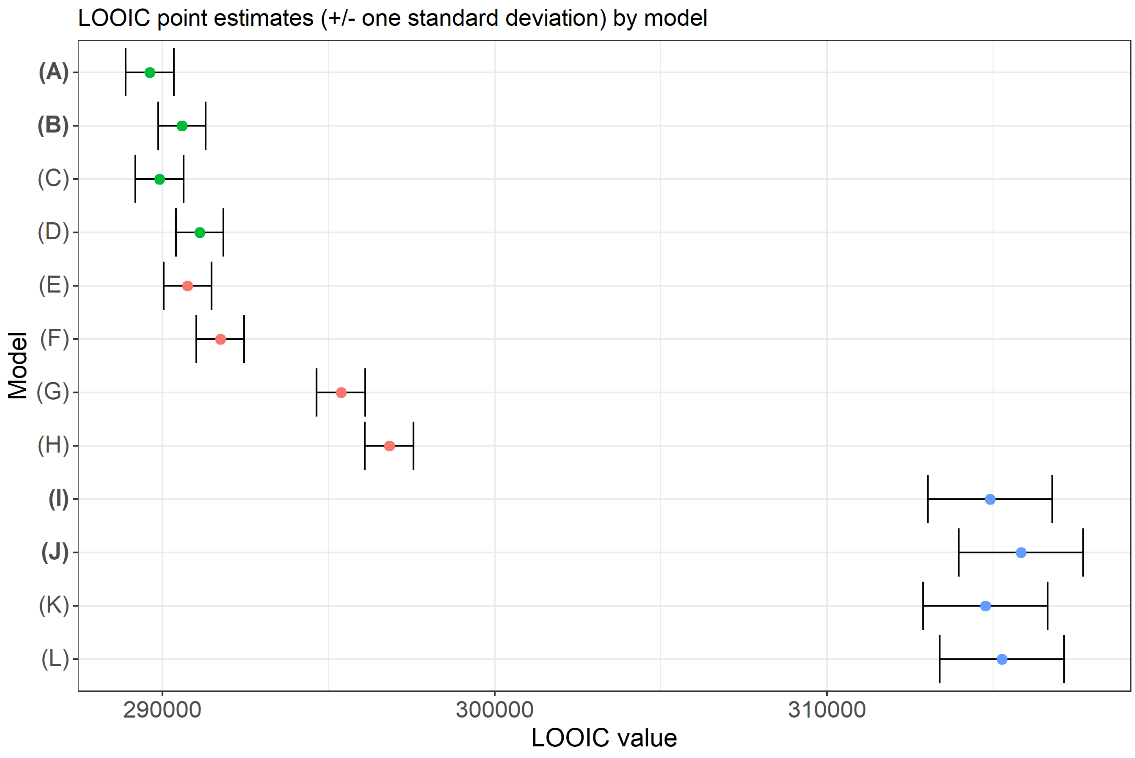

**Figure S72:** The leave-one-out information criterion (LOOIC) point estimate, and one standard deviation range for the 12 models defined in Table S3. Models (A), (B), (I) and (J) in bold are the four models presented in the main manuscript.

### 5 Software and implementation

The primary interface to the model is coded in R [31]. The model is written in Stan, and run with `rstan` v2.21.2. Additional packages used for this paper were `sf` v1.0-3, `dplyr` v1.0.7, `plyr` v1.8.7, `tidyverse` v1.3.1, `janitor` v2.1.0, `spdep` v1.1-11, and `gridExtra` v2.3. The code and scripts used to create the results in this paper are available at https://github.com/thomrawson/Rawson-spatial-covid, and are provided in the `orderly` v1.4.3 reproducible reporting structure.
